# Increased risk of major ischaemic events among autistic people

**DOI:** 10.64898/2026.07.01.26357011

**Authors:** Omar A. Al-Rubaie, Elizabeth Weir, Alex Tsompanidis, Carrie Allison, Matthew C. Fysh, Emanuele Di Angelantonio, Rupert A. Payne, Fiona E. Matthews, Simon Baron-Cohen

## Abstract

**Importance:** Autistic people have increased risks of cardiometabolic conditions and premature mortality; however, no studies specifically assess risks of major ischaemic events in the autistic population.

**Objective:** To determine whether autistic people are at increased risk of major ischaemic events after accounting for known risk factors.

**Design:** A retrospective matched cohort study from 1/1/1990 to 31/12/2019. Cox regression models accounting for matching factors, sociodemographic characteristics, intellectual disability, and cardiovascular risk factors were employed.

**Setting:** This population-based study leveraged lifetime primary and secondary care electronic health records from the Clinical Practice Research Datalink and Hospital Episode Statistics, as well as sociodemographic, ethnicity, and death registration data from the Office of National Statistics.

**Participants:** 23,612 autistic people were matched 1:5 on birth year (±2 years), general practitioner practice ID, and gender to 118,060 non-autistic people. Autistic people were defined as those with a clinical autism diagnosis recorded during the study period. Patients missing Indices of Multiple Deprivation and ethnicity data, and an end date prior to their CPRD start date were excluded along with their matched set.

**Exposure:** Clinical diagnosis of autism. Secondary exposures included health conditions associated with cardiovascular disease with some additional conditions relevant to autism.

**Main Outcome and Measures:** Time to first major ischaemic event (any of myocardial infarction, angina, other ischaemic heart disease, ischaemic stroke, and transient ischaemic attacks).

**Results:** Autistic people had a greater risk of a major ischaemic event in the study period in the minimally adjusted Model 1 (HR 1.19; 95% CI: 1.00, 1.42) as well as for autistic females even after accounting for risk factors (adjusted HR 1.71; 95% CI: 1.10, 2.67). There was also evidence that several cardiometabolic risk factors had a higher prevalence among the autistic group such as severe mental illness, dyslipidaemia, and obesity (all P<0.001).

**Conclusions and Relevance:** Autistic people have an increased risk of major ischaemic events and need improved cardiometabolic risk management. This risk remained in autistic women after adjusting for cardiometabolic and sociodemographic factors. As cardiovascular disease is a primary cause of death globally, research is needed to better understand the mechanism that drives this association.

**Key Points:** *Question:* Do autistic people have higher risks of major ischaemic events than non-autistic people after accounting for cardiometabolic risk factors?

*Findings:* In this retrospective matched cohort study of 141,672 individuals, autistic people had a 19% increased risk of a major ischaemic event. This was 71% for autistic women after accounting for sociodemographic and cardiometabolic factors.

*Meaning:* Elevated risks of these events among autistic women cannot be explained by known cardiometabolic risk factors. Future research should explore factors that may contribute to these risks. Clinicians and health policy makers and providers should implement appropriate screening and cardiometabolic interventions for autistic people.

## Introduction

Autism is a neurodevelopmental condition associated with repetitive behaviour, restricted interests, and differences in socialising and communication.^1^ Approximately 1 in 100 children are diagnosed with autism globally.^2^ The incidence of autism diagnosis is also significantly increasing, with the United Kingdom (UK) seeing a 787% increase between 1998 and 2018.^3^ An estimated 59-72% of autistic people were undiagnosed as of 2018 with underdiagnosis being substantially higher in older and adult populations suggesting age-based inequalities.^4^ Increasing rates of autism diagnosis have also been reported in the United States and globally.^5–7^

When accessing healthcare, autistic patients face numerous challenges and barriers. Up to 80% of autistic adults reported difficulties in visiting a general practitioner, with not feeling understood, having to make appointments via telephone, difficulty deciding if their symptoms justified seeing a doctor, and communication barriers being experienced by most.^8^ This is seen across ages with adults and the families of autistic children both facing barriers to accessing healthcare due to communication and sensory issues.^9,10^

Autistic individuals are understood to have a higher risk of premature mortality, as well as reduced life expectancy both in the presence and absence of co-occurring intellectual disability.^11,12^ Autistic individuals in the UK without intellectual disability had a 1.7-fold higher mortality rate compared to non-autistic individuals.^12^ While suicide is a major factor, physical health conditions also play a fundamental role in increasing the rate of premature mortality.^13^ In a national Swedish cohort, a 2-fold increase in mortality related to the circulatory system was reported for men, although no significant difference was found in women.^11^

As well as an increased risk of premature mortality, survey data has shown an increased prevalence of cardiovascular conditions in both autistic men and women such as arrhythmias.^14^ Additionally, although data on major ischaemic events is limited, a meta-analysis of observational and interventional studies found that autistic people are at an increased risk of heart disease.^15^ It is also recognised that autistic people have higher rates of health conditions associated with cardiovascular disease such as obesity, diabetes, and migraine,^14–18^ but it is unclear to what degree different risk factors increase the risk of cardiovascular disease in the autistic population.

Several risk factors for major ischaemic events are well established in the general population and are already used to inform clinical practice. Such factors can be combined in risk prediction tools, such as the QRISK3 tool used in UK primary care, used to estimate the 10-year risk of a myocardial infarction (MI), angina, stroke, or transient ischaemic attack (TIA).^19,20^ The autistic population has a higher prevalence of a number of the factors that contribute to QRISK3 such as diabetes, dyslipidaemia,^15^ obesity,^17,18^ arrythmias,^14^ although evidence for other risk factors such as migraines is more limited.^16^

Despite this evidence, there are no studies that account for a QRISK3 framing when assessing risks of cardiovascular disease among autistic people. In addition, no studies look at the risk of major ischaemic events specifically in the autistic population. This study seeks to fill the gap in the literature by calculating the risk of major ischaemic events among autistic people compared to matched non-autistic people, account for cardiometabolic factors.

## Methodology

### Study Population

This study leveraged anonymised National Health Service (NHS) Electronic Health Records (EHRs) of n=144,636 patients from Clinical Practice Research Datalink (CPRD) GOLD, which is composed of general practice medical records from over 22 million patients across the UK and is representative of the population. These data were linked to Hospital Episode Statistics (HES) Admitted Patient Care inpatient admission administrative records,^21^ Office of National Statistics (ONS)^22^ mortality data, area-based socioeconomic deprivation data (2015 Index of Multiple Deprivation, IMD),^23^ and ethnicity^24^ data. Clinically diagnosed autistic individuals were matched 1:5 by birth year (±2 years), gender, and GP practice ID to non-autistic individuals who had no autism diagnosis in their clinical record at any time, nor an autistic sibling or mother. Matched sets were excluded where (please see Supplement 1 for exclusion flow diagram): (1) the autistic individual only had a referral code for autism (rather than evidence of a recorded clinical diagnosis) along with their matches; (2) any individual had missing data for (IMD); (3) or any individual’s end date (see below) was prior to their start date (see below). The final sample size was n=141,672, including n=23,612 autistic individuals.

For the purposes of this study, autistic patients were defined by having at least one medical code for autism diagnosis (see Supplement 2) in their clinical GP practice record. The study period was 1^st^ of January 1990 until the 31^st^ of December 2019. The end date for each patient was the earliest of the study end date, ONS death date,^22^ CPRD death date, or outcome event date. The start date for each patient was the latest of the study start date or the patient’s first registration.

### Statistical Analysis

Demographic data along with covariate and outcome prevalence was collected with demographics, covariate, and outcome prevalence differences were assessed using Chi-square tests via the ‘chisq.test’ function in R. Outcome (see Supplement 3 for relevant code lists) and covariate (please see Supplement 4) prevalence was defined as the first recorded medcode and/or ICD-10 code. In addition, the age at which each patient of both genders had their first major ischaemic event was recorded and plotted in a density graph, the mean age was calculated, and a Mann-Whitney U test was carried out to assess significance.

Cox proportional hazards regression models were utilised on four groups (all patients, male and female separately, and excluding patients with intellectual disability via the ‘survival’ and ‘survminer’ packages in R version 4.3.2. In the model time was measured by using the patients age in days at their start date and their age in days at their outcome date, or end date in those with no outcomes. The primary outcome of interest was the first instance of a major ischaemic event during the study period, defined as a composite outcome comprised of five components: MI, angina, other coronary artery diseases such as atherosclerotic heart disease or coronary heart disease, TIA, and ischaemic stroke. Outcomes were defined by the presence of relevant medical codes in the CPRD GP records,^25^ ICD-10 codes in HES hospitalisation data,^21^ and ICD-10 codes in ONS death data.^22^

Models were adjusted for the cohort matching criteria as well as socioeconomic status (IMD),^23^ ethnicity (white as baseline),^24^ and binary covariates (all based on diagnostic codes) of; atrial fibrillation and flutter, chronic kidney disease (CKD), erectile dysfunction, family history of MI (< 60 years of age and < 55 in 1^st^ degree male relative) and angina (< 60 years of age, < 55 in 1^st^ degree male relative, and < 65 in 1^st^ degree female relative), hypertension, migraine, obesity, severe mental illness (SMI), smoking status (coded yes and no), diabetes mellitus, depression, alcohol misuse, intellectual disability (ID), and dyslipidaemia. As with outcomes, covariates were defined by the presence of medical codes (medcodes) in the CPRD GP records as well as ICD-10 codes in the linked HES data dated prior to each patients first event or end date.^21^

Four models were constructed as follows, with each model in turn adding further covariates to the previous model: Model 1 included the matching criteria only (gender, age, GP practice); Model 2 additionally adjusted for key lifestyle factors (alcohol misuse, smoking status), sociodemographic factors (IMD, ethnicity) and intellectual disability; Model 3 additionally adjusted for obesity (a key cardiometabolic risk factor with major behavioural and environmental determinants); and Model 4 was a fully adjusted model, including all matching factors, sociodemographic factors, and health conditions associated with cardiovascular risk. All covariates met the proportional hazards assumption other than birth year, GP practice ID, family history of MI and angina, alcohol misuse, obesity, chronic kidney disease, hypertension, migraines, and diabetes mellitus. Given that these were adjustment variables rather than exposures of primary interest, mild violations are unlikely to bias the estimates of autism. All covariates met the linearity assumption other than birth year; however, this was a matching variable rather than exposures of primary interest, and such violations are unlikely to bias the estimated effect of autism.

Finally, a gender-stratified analysis, and an analysis excluding those with ID, was performed using the same four Cox regression model designs previously used. Male and female-only groups were formed using the gender recorded in each patient’s GP record, with two genders listed, ‘male’ and ‘female,’ as there were no patients with other genders recorded. The male-only group size was n=111,006, of which n=18,501 were autistic. The female-only group had a size of n=30,666, of which n=5,111 were autistic. Both sets of gender-stratified models excluded gender, and the female-only models also excluded erectile dysfunction. The no-ID group had a size of n=129,396, of which n=21,566 were autistic. The interaction between autism and gender was then tested for each model using the ‘survival’ package in R.

## Results

Both autistic and non-autistic groups were matched and demographically similar to one another in birth year, region, and gender ratio. Based on the characteristics of the autistic group, the study was over 75% male, with mean birth year of 1996 (standard deviation of 12.8 years). While there were participants at GP practices in every region of the UK, the East Midlands had the highest percentage at 34.7%, and greater London had the lowest with 1.25%; this reflects regional variation in use of the Vision GP clinical informatics system that contributed data to the CPRD GOLD database. The complete range of IMD was well represented across the cohort. There was strong evidence (p<0.001) for slight differences in the distribution of IMD between autistic and non-autistic patients, with slightly lower and higher rates of deprivation in each group respectively. The two groups were both majority white, although there were slight differences in distribution of ethnicities (p<0.001), and lower rates of unknown ethnicity in autistic (5.7%) compared to non-autistic (10.3%) patients. The autistic group had a significantly higher prevalence of intellectual disability (8.1% vs 0.2%, p<0.001).

**Table 1:** Description of the Cohort.

|  | *Autistic group*<br>*N=23,612* |  | *Non-Autistic group*<br>*N = 118,060* |  | *P-Value* |
| --- | --- | --- | --- | --- | --- |
| *Mean Birth Year (±SD)* | 1996 | (12.8) | 1996 | (12.8) |  |
| ***Birth Year Ranges:*** |  |  |  |  |  |
| *1905 – 1927* | 9 | (0.04%) | 45 | (0.04%) |  |
| *1928 – 1950* | 170 | (0.72%) | 850 | (0.72%) |  |
| *1951 – 1973* | 1509 | (6.39%) | 7545 | (6.39%) |  |
| *1974 – 1996* | 8309 | (35.2%) | 41,609 | (35.2%) |  |
| *1997 – 2019* | 13,615 | (57.7%) | 68,011 | (57.7%) |  |
| ***Gender:*** |  |  |  |  |  |
| *Male* | 18,501 | (78.4%) | 92,505 | (78.4%) |  |
| *Female* | 5111 | (21.6%) | 25,555 | (21.6%) |  |
| ***IMD Ranges:*** |  |  |  |  | <0.001 |
| *1 – 5 (most deprived)* | 5563 | (23.6%) | 31,644 | (26.8%) |  |
| *6 – 10* | 5816 | (24.6%) | 30,143 | (25.5%) |  |
| *11 – 15* | 6109 | (25.9%) | 28,549 | (24.2%) |  |
| *16 – 20 (least deprived)* | 6124 | (25.9%) | 27,724 | (23.5%) |  |
| ***Ethnicity:*** |  |  |  |  | <0.001 |
| *1 (Asian)* | 918 | (3.89%) | 5875 | (4.98%) |  |
| *2 (Black)* | 856 | (3.63%) | 3784 | (3.21%) |  |
| *3 (Mixed/Multiple)* | 641 | (2.71%) | 2713 | (2.30%) |  |
| *4 (White)* | 19,786 | (83.8%) | 92,838 | (78.6%) |  |
| *5 (Other)* | 70 | (0.30%) | 680 | (0.58%) |  |
| *6 (Unknown)* | 1341 | (5.68%) | 12,170 | (10.3%) |  |
| ***Intellectual Disability*** | 1907 | (8.08%) | 184 | (0.16%) | <0.001 |
| ***GP Practice Region:*** |  |  |  |  |  |
| *1 (Greater London)* | 296 | (1.25%) | 1480 | (1.25%) |  |
| *2 (South East)* | 3031 | (12.8%) | 15,155 | (12.8%) |  |
| *3 (South West)* | 585 | (2.48%) | 2925 | (2.48%) |  |
| *4 (West Midlands)* | 469 | (1.99%) | 2345 | (1.99%) |  |
| *5 (North West)* | 3116 | (13.2%) | 15,580 | (13.2%) |  |
| *6 (North East)* | 1921 | (8.14%) | 9605 | (8.14%) |  |
| *7 (Yorkshire and Humber)* | 3272 | (13.9%) | 16,360 | (13.9%) |  |
| *8 (East Midlands)* | 8201 | (34.7%) | 41,005 | (34.7%) |  |
| *9 (East of England)* | 2721 | (11.5%) | 13,605 | (11.5%) |  |

*IMD, Index of Multiple Deprivation; GP, General Practitioner.*
*Significance calculated using Chi-square tests.*

The autistic group had a higher prevalence of several important risk factors (Table 2). The greatest differences were seen in obesity (5.0% vs 2.6%, p<0.001), SMI (4.5% vs 0.6%, p<0.001), and depression (14.9% vs 6.8%, p<0.001) compared to the non-autistic group. There was also higher prevalence for other key cardiometabolic conditions including hypertension (2.4% vs 2.0%, p=0.001), dyslipidaemia (1.4% vs 1.1%, p<0.001) and diabetes (1.9% vs 1.2%, p<0.001). However, smoking prevalence (13% vs 15%, p<0.001) and cardiovascular family history (0.5% vs 0.7%, p=0.01) were lower in the autistic group, and there was no evidence of a difference in other key risk factors (e.g. migraine and atrial fibrillation and flutter).

Prevalence of cardiovascular outcomes are presented in Table 2. The autistic group had a higher prevalence of stroke (0.26% vs 0.14%, p<0.001), but lower prevalence of cardiac ischaemia (MI 0.16% vs 0.22%, p=0.08; other cardiac ischaemia 0.31% vs 0.43%, p=0.007). There was no significant difference in the prevalence of a first major ischaemic event.

**Table 2:**
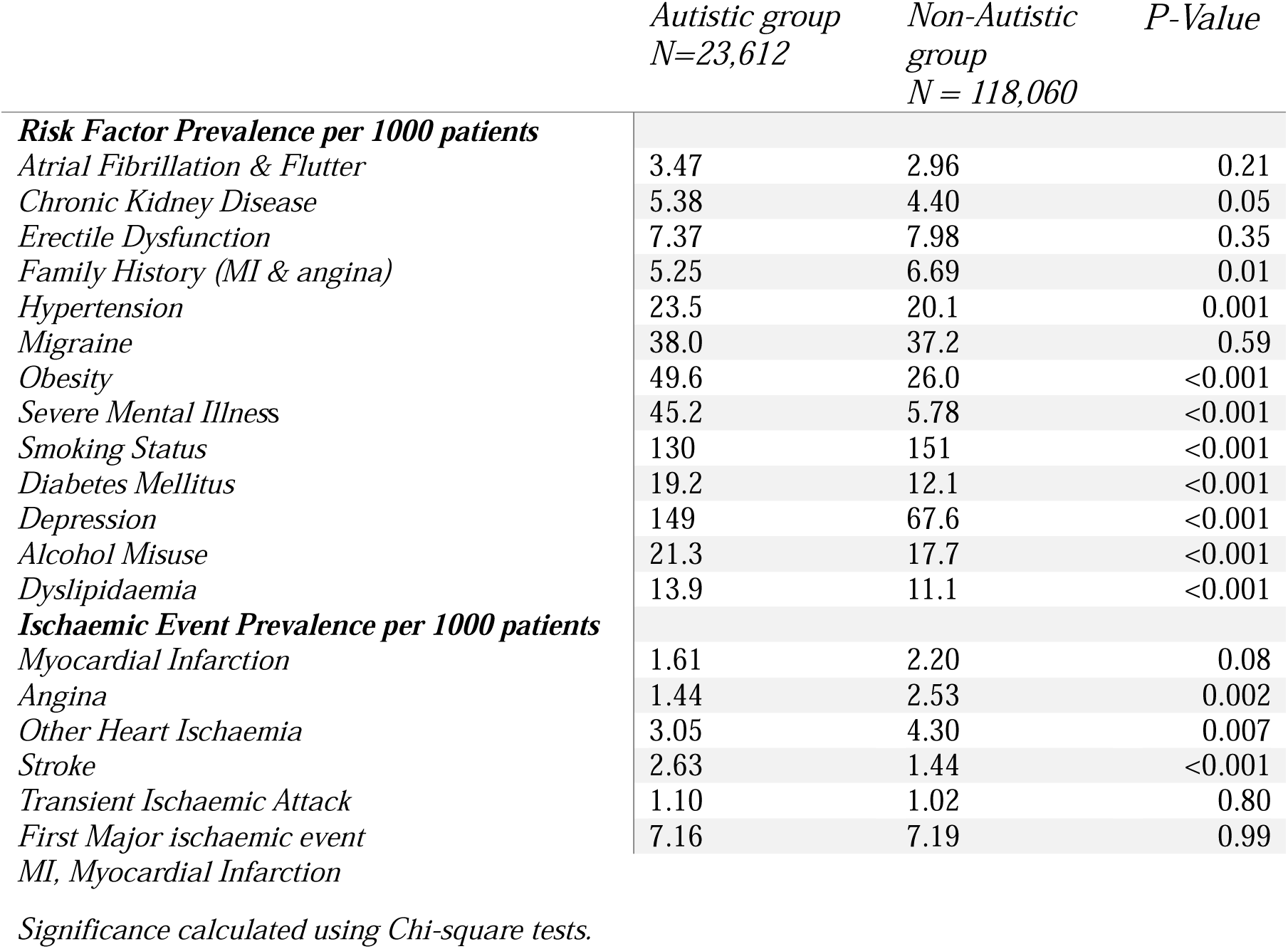
Risk Factor and Outcome Prevalence.

Table 3 reports the association between autism and ischaemic outcomes. In all models, autistic people had a higher risk of major ischaemic events compared to matched non-autistic people during the duration of the study period, with the increased risk being significant in all models other than in the fully adjusted Model 4 (hazard ratio (HR): 1.16; 95% CI: 0.94, 1.44). There was a slight increase of this association when adjusting for fewer factors in Model 2 (HR: 1.29; 95% CI: 1.06, 1.56).

**Table 3:** Hazard Ratios for Autism and Co-Variates Across Cox Models.

|  | *Model 1* |  | *Model 2* |  | *Model 3* |  | *Model 4* |  |
| --- | --- | --- | --- | --- | --- | --- | --- | --- |
|  | *Hazard Ratio* | *P-Value* | *Hazard Ratio* | *P-Value* | *Hazard Ratio* | *P-Value* | *Hazard Ratio* | *P-Value* |
| *Autism* | 1.19 (1.00, 1.42) | 0.05 | 1.29 (1.06, 1.56) | 0.01 | 1.27 (1.04, 1.54) | 0.02 | 1.16 (0.94, 1.44) | 0.17 |
| *IMD* | - | - | 1.02 (1.01, 1.03) | <0.001 | 1.02 (1.01, 1.03) | 0.003 | 1.02 (1.00, 1.03) | 0.008 |
| *Ethnicity 1 (Asian)* | - | - | 1.43 (1.03, 1.99) | 0.03 | 1.43 (1.03, 1.99) | 0.03 | 1.20 (0.86, 1.69) | 0.29 |
| *Ethnicity 2 (Black)* | - | - | 1.52 (1.01, 2.29) | 0.04 | 1.53 (1.03, 2.28) | 0.04 | 1.39 (0.94, 2.07) | 0.10 |
| *Ethnicity 3 (Mixed/Multiple)* | - | - | 1.14 (0.51, 2.55) | 0.74 | 1.16 (0.52, 2.59) | 0.71 | 1.08 (0.51, 2.26) | 0.84 |
| *Ethnicity 5 (Other)* | - | - | 0.46 (0.11, 1.90) | 0.29 | 0.46 (0.11, 1.90) | 0.28 | 0.47 (0.15, 1.47) | 0.19 |
| *Ethnicity 6 (Unknown)* | - | - | 0.27 (0.18, 0.41) | <0.001 | 0.28 (0.19, 0.43) | <0.001 | 0.46 (0.30, 0.70) | <0.001 |
| *Alcohol Misuse* | - | - | 1.61 (1.30, 2.01) | <0.001 | 1.60 (1.29, 1.99) | <0.001 | 1.32 (1.06, 1.66) | 0.01 |
| *Smoking Status* | - | - | 1.85 (1.62, 2.13) | <0.001 | 1.84 (1.60, 2.11) | <0.001 | 1.56 (1.35, 1.80) | <0.001 |
| *Intellectual Disability* | - | - | 0.77 (0.52, 1.13) | 0.18 | 0.77 (0.52, 1.12) | 0.17 | 1.02 (0.68, 1.51) | 0.94 |
| *Obesity* | - | - | - | - | 1.73 (1.44, 2.08) | <0.001 | 1.04 (0.86, 1.26) | 0.66 |
| *Atrial Fibrillation & Flutter* | - | - | - | - | - | - | 1.68 (1.35, 2.10) | <0.001 |
| *CKD* | - | - | - | - | - | - | 1.25 (0.96, 1.62) | 0.09 |
| *Erectile Dysfunction* | - | - | - | - | - | - | 0.92 (0.72, 1.17) | 0.48 |
| *Family History (MI & Angina)* | - | - | - | - | - | - | 5.85 (4.82, 7.11) | <0.001 |
| *Hypertension* | - | - | - | - | - | - | 2.21 (1.85, 2.65) | <0.001 |
| *Migraine* | - | - | - | - | - | - | 1.14 (0.91, 1.44) | 0.25 |
| *SMI<sup>d</sup>* | - | - | - | - | - | - | 1.73 (1.32, 2.25) | <0.001 |
| *Diabetes Mellitus* | - | - | - | - | - | - | 1.33 (1.07, 1.65) | 0.009 |
| *Depression* | - | - | - | - | - | - | 1.20 (1.03, 1.40) | 0.02 |
| *Dyslipidaemia* | - | - | - | - | - | - | 1.97 (1.65, 2.35) | <0.001 |

IMD, Indices of Multiple Deprivation; CKD, Chronic Kidney Disease; MI, Myocardial Infarction; SMI, Severe Mental Illness.
*Model 1: minimally adjusted model containing the matching criteria gender, practice ID, and birth year ( $\pm 2$ years); Model 2: as Model 1, plus adjustment for alcohol misuse, socioeconomic status (IMD 2015), ethnicity (white as baseline), smoking status, and intellectual disability; Model 3: as Model 2, plus adjustment for obesity; Model 4: fully adjusted model containing all matching, demographic, and risk factor covariates.*

The same pattern of risk was observed when stratifying by gender, and after exclusion of patients with ID (Figure 1). Effect sizes were lower and non-significant in males. Effect sizes were substantially higher in females, but confidence intervals were wide reflecting smaller numbers.

**Figure 1:**
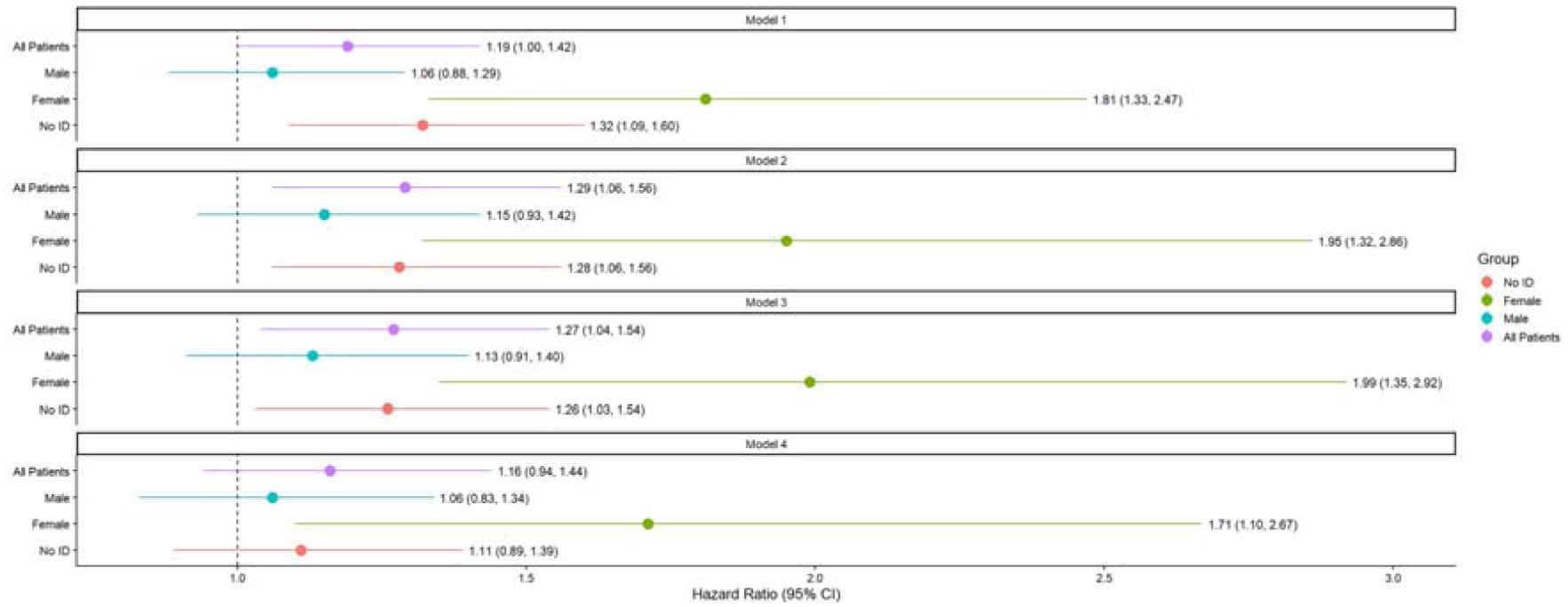
Forest plot of Autism hazard ratios for gender-stratified Cox models and models excluding intellectual disability compared to the whole cohort.

The age at which autistic and non-autistic patients had their first major ischaemic events appeared to vary in distribution, with the peaks occurring in middle age. Autistic people had a lower mean age of major ischaemic events than non-autistic people. This was significant for overall events and for TIAs.

**Figure 2:**
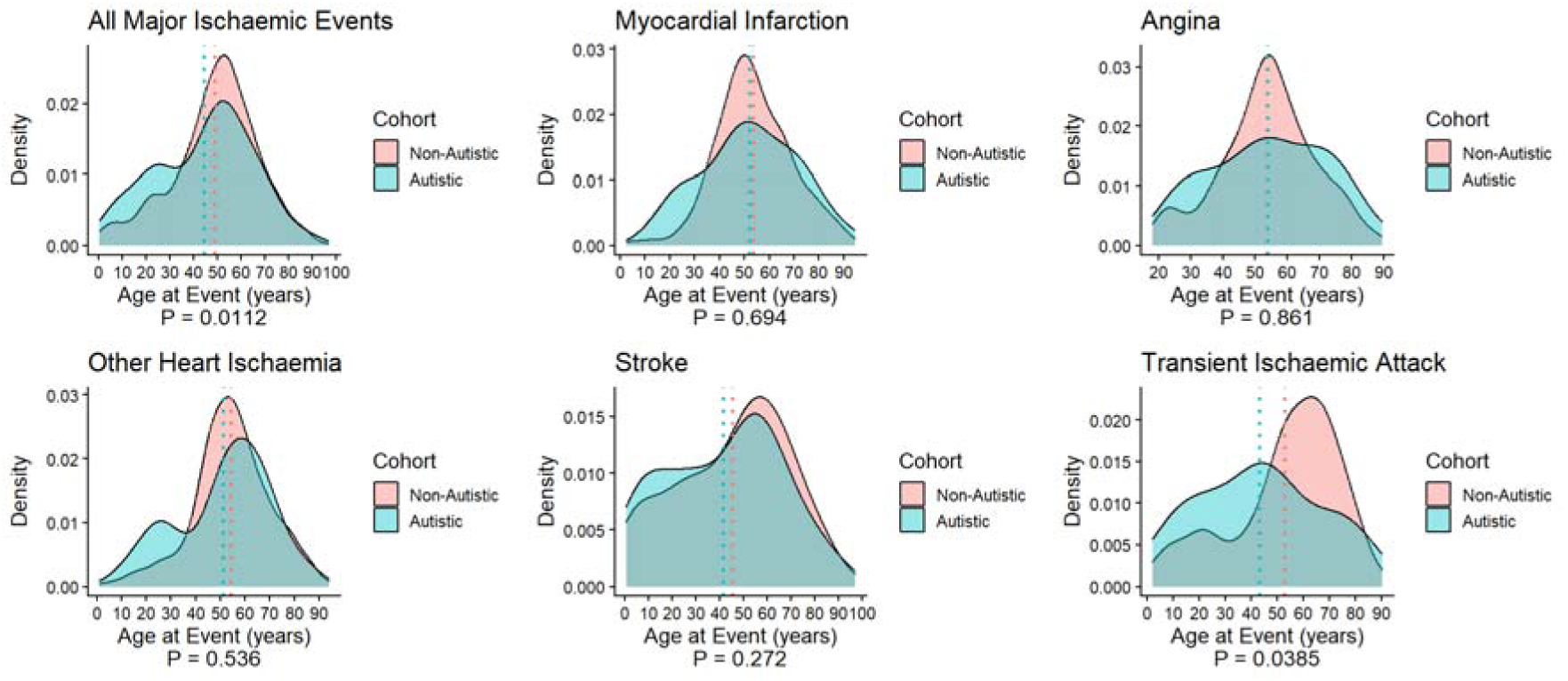
Density plots of age at first major ischaemic event by autism diagnosis and event type.

There was a significant interaction between autism and gender in all models (please see Supplement 5 for the table), with the hazard ratio being highest in the minimally adjusted Model 1 (HR: 0.60; 95% CI: 0.43, 0.83) and lowest in the fully adjusted Model 4 (HR: 0.57; 95% CI: 0.39, 0.84).

## Discussion

### Summary of Key Findings

This study has shown a significantly increased risk of major ischaemic events in the autistic population. This finding warrants further research, especially as this risk remained after accounting for other cardiovascular risk factors and demographics in autistic females but not significantly in the other groups, such as when excluding those with intellectual disability. The autistic population also have these events significantly younger. The gender stratified analysis of event risk showed the significantly increased risk remained for the autistic females, but was no longer significant for males, with autistic females being at a markedly greater risk compared to non-autistic females. The interaction testing between autism and gender (being male) found a buffering effect, with the hazard being significantly lower than expected highlighting this gender difference in risk.

While there was no significant difference between autistic and non-autistic people in the prevalence of a first event, autistic people had a significantly higher prevalence of strokes but significantly lower prevalence of cardiac ischaemia. This suggests that while there are no differences in the composite outcome, autistic and non-autistic people have different ischaemic disease profiles that would benefit from further research.

### Strengths and Limitations

The cox modelling incrementally adds covariates based on whether it is demographic, part-modifiable, or a non-modifiable, to analyse these categories separately. However, a limitation of this model design is that Model 4 covaries for plausible mediators of the autism-cardiovascular disease pathway, such as diabetes and hypertension, as well as confounders.

Although the autistic and non-autistic groups were matched and demographically similar, they were not representative of England’s population. Only 1.25% of each group were registered with a GP practice in Greater London, which is the most populous city in the England according to the 2021-1991 census,^26–29^ with its population in mid-2019 estimated as 8,962,000.^30^ England’s population in the same year was estimated to be 56,286,961,^31^ meaning 15.9% of the population lived in London. This suggests that the autistic population in this cohort may be biased away from London. As well as this, both groups were 78.4% male compared to the general population being 49.4% male,^31^ and over half of both groups were born after 1997 and would be a maximum age of 22 years by the study end date. This means that the autistic population was also younger compared to the general population where 27.2% of people in England were 22 years old or younger in 2019.^31^

While the study population was majority white, all ethnicities were represented in a broadly similar way to what is seen in the general population other than ‘Asian’ and ‘Other’ who are underrepresented in this study.^32^ However, the groups were ethnically significantly different. The study population was also distributed across IMD ranges, but there was a significant difference in distribution, with the non-autistic group being more represented in the least deprived bands. Intellectual disability was significantly more prevalent in the autistic group. This large and diverse study population makes this a strong study, despite the limitations of the population being relatively young.

### Interpretation in the Context of Other Literature

There were significant differences between the autistic and non-autistic population in risk factor and outcome prevalence. The autistic group had an elevated prevalence of cardiometabolic risk factors widely accepted in previous literature,^20^ such as hypertension, obesity, diabetes, and severe mental illness. The higher prevalence of dyslipidaemia observed may also suggest metabolic or dietary differences between the autistic and non-autistic population and is congruent with existing literature,^15^ providing direction on which cardiometabolic risk factors could be targeted to improve the health of autistic patients.

There were two risk factors the autistic population had a significantly reduced prevalence of which were smoking status, which aligns with previous literature,^33^ and family history of an early MI or angina, which appears to be a novel finding. The outcomes to see a significant difference were stroke, which was elevated in the autistic group, and MI, angina, and other ischaemic coronary diseases, which were elevated in the non-autistic group.

It may be the case that autistic people have increased risk of major ischaemic events due to specific genetic or endocrine factors that appear to be more common among autistic people, such as polyendocrine metabolic ovarian syndrome (PMOS).^34–36^ In addition, autistic people are less likely than non-autistic people to meet guidelines for diet, exercise and sleep, and are more likely to be obese or underweight, which are all known environmental risk factors for cardiovascular conditions.^17^

### Implications for Policy and Future Research

As autistic patients are at a higher risk of major ischaemic events and are having them younger, they may benefit from earlier risk management and closer monitoring of their cardiometabolic health, especially as this risk became non-significant after account for cardiometabolic risk factors. More research into the relationship between autism and stroke risk may also be warranted. It would be especially to further research a potential mechanism behind the remaining risk experienced by autistic females. However, in this cohort the number of stroke events was too low to study this outcome individually.

## Conclusion

This retrospective matched cohort study has identified the autistic population, including those without ID, as being at a greater lifetime risk of major ischaemic events, a risk that is greater and remains for autistic females even after for accounting for socioeconomic data and widely accepted cardiometabolic risk factors. This study has also provided insight into the broader cardiometabolic health of the autistic population by providing the prevalence data of the risk factors included in the cox regression modelling. Finally, the age at which these events occurred was analysed, demonstrating that the autistic population are having their first event younger.

## Data Availability

The medcode and ICD-10 code lists used in this study are available in the supplement.

## Acknowledgments

SBC received funding from the Wellcome Trust 214322\Z\18\Z. SBC also received funding from the C (IMI) 2 Joint Undertaking under grant agreement No 777394 for the project AIMS-2-TRIALS. This Joint Undertaking receives support from the European Union’s Horizon 2020 research and innovation programme and EFPIA and Autism Speaks, Autistica, and SFARI. SBC also received funding from the Autism Research Trust (now managed by Autism Action) and from SFARI, the European Commission, the Templeton World Charitable Fund, the Stoneygate Trust, the MRC, the Rosetrees Trust, the Eagles Autism Foundation, the K. Lisa Yang Centre for Autism Research at Cambridge/Yang Tan Collective, the Cambridge University Development and Alumni Relations (CUDAR), the NIHR Cambridge Biomedical Research Centre (NIHR203312), and the NIHR Applied Research Collaboration East of England. Any views expressed are those of the author(s) and not necessarily those of the funders (including IHI-JU2, or the NIHR or the Department of Health and Social Care). The funders had no role in the design of the study, in the collection, analyses, or interpretation of data, in the writing of the manuscript, or in the decision to publish the results. For the purpose of Open Access, the author has applied a CC BY public copyright licence to any Author Accepted Manuscript version arising from this submission.

## References

1. Lai MC, Lombardo MV, Baron-Cohen S. Autism. Lancet. Mar 8 2014;383(9920):896–910. doi:10.1016/S0140-6736(13)61539-1

2. Zeidan J, Fombonne E, Scorah J, et al. Global prevalence of autism: A systematic review update. Autism Res. May 2022;15(5):778–790. doi:10.1002/aur.2696

3. Russell G, Stapley S, Newlove-Delgado T, et al. Time trends in autism diagnosis over 20 years: a UK population-based cohort study. J Child Psychol Psychiatry. Jun 2022;63(6):674–682. doi:10.1111/jcpp.13505

4. O’Nions E, Petersen I, Buckman JEJ, et al. Autism in England: assessing underdiagnosis in a population-based cohort study of prospectively collected primary care data. Lancet Reg Health Eur. Jun 2023;29:100626. doi:10.1016/j.lanepe.2023.100626

5. Schendel DE, Thorsteinsson E. Cumulative Incidence of Autism Into Adulthood for Birth Cohorts in Denmark, 1980-2012. JAMA. Nov 6 2018;320(17):1811–1813. doi:10.1001/jama.2018.11328

6. Raz R, Weisskopf MG, Davidovitch M, Pinto O, Levine H. Differences in autism spectrum disorders incidence by sub-populations in Israel 1992-2009: a total population study. J Autism Dev Disord. Apr 2015;45(4):1062–9. doi:10.1007/s10803-014-2262-z

7. Grosvenor LP, Croen LA, Lynch FL, et al. Autism Diagnosis Among US Children and Adults, 2011-2022. JAMA Netw Open. Oct 1 2024;7(10):e2442218. doi:10.1001/jamanetworkopen.2024.42218

8. Doherty M, Neilson S, O’Sullivan J, et al. Barriers to healthcare and self-reported adverse outcomes for autistic adults: a cross-sectional study. BMJ Open. Feb 22 2022;12(2):e056904. doi:10.1136/bmjopen-2021-056904

9. Mason D, Ingham B, Urbanowicz A, et al. A Systematic Review of What Barriers and Facilitators Prevent and Enable Physical Healthcare Services Access for Autistic Adults. J Autism Dev Disord. Aug 2019;49(8):3387–3400. doi:10.1007/s10803-019-04049-2

10. Babalola T, Sanguedolce G, Dipper L, Botting N. Barriers and Facilitators of Healthcare Access for Autistic Children in the UK: a Systematic Review. Rev J Autism Dev Dis. Dec 2025;12(4):780–808. doi:10.1007/s40489-023-00420-3

11. Hirvikoski T, Mittendorfer-Rutz E, Boman M, Larsson H, Lichtenstein P, Bolte S. Premature mortality in autism spectrum disorder. Br J Psychiatry. Mar 2016;208(3):232–8. doi:10.1192/bjp.bp.114.160192

12. O’Nions E, Lewer D, Petersen I, et al. Estimating life expectancy and years of life lost for autistic people in the UK: a matched cohort study. Lancet Reg Health Eur. Jan 2024;36:100776. doi:10.1016/j.lanepe.2023.100776

13. Mandell D. Dying before their time: Addressing premature mortality among autistic people. Autism. Apr 2018;22(3):234–235. doi:10.1177/1362361318764742

14. Weir E, Allison C, Warrier V, Baron-Cohen S. Increased prevalence of non-communicable physical health conditions among autistic adults. Autism. Apr 2021;25(3):681–694. doi:10.1177/1362361320953652

15. Dhanasekara CS, Ancona D, Cortes L, et al. Association Between Autism Spectrum Disorders and Cardiometabolic Diseases: A Systematic Review and Meta-analysis. JAMA Pediatr. Mar 1 2023;177(3):248–257. doi:10.1001/jamapediatrics.2022.5629

16. Vetri L. Autism and Migraine: An Unexplored Association? Brain Sci. Sep 6 2020;10(9)doi:10.3390/brainsci10090615

17. Weir E, Allison C, Ong KK, Baron-Cohen S. An investigation of the diet, exercise, sleep, BMI, and health outcomes of autistic adults. Mol Autism. May 8 2021;12(1):31. doi:10.1186/s13229-021-00441-x

18. Hill AP, Zuckerman KE, Fombonne E. Obesity and Autism. Pediatrics. Dec 2015;136(6):1051–61. doi:10.1542/peds.2015-1437

19. Endeavour Predict CIC. QRISK3. Accessed 16/04/2025, 2025. https://qrisk.org/

20. Hippisley-Cox J, Coupland C, Brindle P. Development and validation of QRISK3 risk prediction algorithms to estimate future risk of cardiovascular disease: prospective cohort study. BMJ. May 23 2017;357:j2099. doi:10.1136/bmj.j2099

21. (MHRA) MaHPRA. Hospital Episode Statistics (HES) Admitted Patient Care and CPRD primary care data Documentation (set 22/January 2022). https://www.cprd.com/sites/default/files/2024-11/HES_APC_Documentation_v2.9.pdf

22. (MHRA) MaHPRA. ONS death registration data and CPRD primary care data Documentation (set 22/January 2022). https://cprd.com/sites/default/files/2022-02/Documentation_Death_set22_v2.6.pdf

23. Statistics OfN. English indicies of deprivation 2015. Ministry of Housing, Communities & Local Government (2018-2021). 2015;

24. (MHRA) MaHPRA. CPRD Ethnicity Records Documentation. Accessed September 2025, https://www.cprd.com/sites/default/files/2025-09/CPRD_EthnicityRecord_Documentation_v1.2.pdf

25. Datalink CPR. CPRD Gold June 2024. Clinical Practice Research Datalink. 2024;

26. Statistics OfN. Population and household estimates, England and Wales: Census 2021. Accessed 02/12/2025, 2025. https://www.ons.gov.uk/peoplepopulationandcommunity/populationandmigration/populationestimates/bulletins/populationandhouseholdestimatesenglandandwales/census2021

27. Statistics OfN. 2011 Census. Accessed 02/12/2025, 2025. https://www.nomisweb.co.uk/sources/census_2011

28. Statistics OfN. 2001 Census. Accessed 02/12/2025, 2025. https://www.nomisweb.co.uk/sources/census_2001

29. Statistics OfN. 1991 Census. Accessed 02/12/2025, 2025. https://www.nomisweb.co.uk/sources/census_1991

30. London Assembly TMoL. London Population. What is the current population of London? Accessed 02/12/2025, 2025. https://www.london.gov.uk/who-we-are/what-london-assembly-does/questions-mayor/find-an-answer/london-population

31. Statistics OfN. Population estimates for the UK, England and Wales, Scotland and Northern Ireland: mid-2019. Accessed 02/12/2025, 2025. https://www.ons.gov.uk/peoplepopulationandcommunity/populationandmigration/populationestimates/bulletins/annualmidyearpopulationestimates/mid2019estimates

32. Statistics OfN. Population of England and Wales. Accessed 06/05/2026, 2026. https://www.ethnicity-facts-figures.service.gov.uk/uk-population-by-ethnicity/national-and-regional-populations/population-of-england-and-wales/latest/#main-facts-and-figures

33. Bejerot S, Nylander L. Low prevalence of smoking in patients with autism spectrum disorders. Psychiatry Res. Jul 15 2003;119(1-2):177–82. doi:10.1016/s0165-1781(03)00123-9

34. Cherskov A, Pohl A, Allison C, Zhang H, Payne RA, Baron-Cohen S. Polycystic ovary syndrome and autism: A test of the prenatal sex steroid theory. Transl Psychiatry. Aug 1 2018;8(1):136. doi:10.1038/s41398-018-0186-7

35. Nautiyal H, Jaiswar A, Roy KK, Dwivedi S. Deciphering the molecular connections between polycystic ovarian syndrome and autism spectrum disorder using bioinformatic analysis. Horm Behav. Jan 2026;177:105859. doi:10.1016/j.yhbeh.2025.105859

36. Teede HJ, Khomami MB, Morman R, et al. Polyendocrine metabolic ovarian syndrome, the new name for polycystic ovary syndrome: a multistep global consensus process. Lancet. May 12 2026;doi:10.1016/S0140-6736(26)00717-8

