## Supplement 3 for "Increased risk of major ischaemic events among autistic people"

**Supplement 3 – Outcome Medcode and ICD-10 Code Lists:**

| outcome | medcode | clinicalevents | referralevents | testevents | immunisationevents | readcode | readterm | databasebuild |
| --- | --- | --- | --- | --- | --- | --- | --- | --- |
| Myocardial Infarction | 241 | 317591 | 10968 | 75 | 0 | G30..00 | Acute myocardial infarction | Feb-09 |
| Myocardial Infarction | 13566 | 801 | 7 | 0 | 0 | G30..11 | Attack - heart | Feb-09 |
| Myocardial Infarction | 2491 | 3760 | 162 | 1 | 0 | G30..12 | Coronary thrombosis | Feb-09 |
| Myocardial Infarction | 30421 | 267 | 5 | 0 | 0 | G30..13 | Cardiac rupture following myocardial infarction (MI) | Feb-09 |
| Myocardial Infarction | 1204 | 6324 | 300 | 4 | 0 | G30..14 | Heart attack | Feb-09 |
| Myocardial Infarction | 1677 | 120837 | 1125 | 15 | 0 | G30..15 | MI - acute myocardial infarction | Feb-09 |
| Myocardial Infarction | 13571 | 533 | 0 | 0 | 0 | G30..16 | Thrombosis - coronary | Feb-09 |
| Myocardial Infarction | 17689 | 698 | 12 | 0 | 0 | G30..17 | Silent myocardial infarction | Feb-09 |
| Myocardial Infarction | 12139 | 2930 | 22 | 0 | 0 | G300.00 | Acute anterolateral infarction | Feb-09 |
| Myocardial Infarction | 5387 | 3305 | 60 | 0 | 0 | G301.00 | Other specified anterior myocardial infarction | Feb-09 |
| Myocardial Infarction | 40429 | 167 | 0 | 0 | 0 | G301000 | Acute anteroapical infarction | Feb-09 |
| Myocardial Infarction | 17872 | 1781 | 12 | 0 | 0 | G301100 | Acute anteroseptal infarction | Feb-09 |
| Myocardial Infarction | 8935 | 2878 | 20 | 0 | 0 | G302.00 | Acute inferolateral infarction | Feb-09 |
| Myocardial Infarction | 29643 | 895 | 8 | 0 | 0 | G303.00 | Acute inferoposterior infarction | Feb-09 |
| Myocardial Infarction | 23892 | 1350 | 5 | 0 | 0 | G304.00 | Posterior myocardial infarction NOS | Feb-09 |
| Myocardial Infarction | 14898 | 1128 | 9 | 0 | 0 | G305.00 | Lateral myocardial infarction NOS | Feb-09 |
| Myocardial Infarction | 63467 | 83 | 0 | 0 | 0 | G306.00 | True posterior myocardial infarction | Feb-09 |
| Myocardial Infarction | 3704 | 4629 | 195 | 0 | 0 | G307.00 | Acute subendocardial infarction | Feb-09 |
| Myocardial Infarction | 9507 | 1246 | 22 | 0 | 0 | G307000 | Acute non-Q wave infarction | Feb-09 |
| Myocardial Infarction | 10562 | 110679 | 194 | 1 | 0 | G307100 | Acute non-ST segment elevation myocardial infarction | Feb-09 |
| Myocardial Infarction | 1678 | 21874 | 218 | 0 | 0 | G308.00 | Inferior myocardial infarction NOS | Feb-09 |
| Myocardial Infarction | 30330 | 169 | 0 | 0 | 0 | G309.00 | Acute Q-wave infarct | Feb-09 |
| Myocardial Infarction | 17133 | 1144 | 13 | 0 | 0 | G30A.00 | Mural thrombosis | Feb-09 |
| Myocardial Infarction | 32854 | 108 | 0 | 0 | 0 | G30B.00 | Acute posterolateral myocardial infarction | Feb-09 |
| Myocardial Infarction | 29758 | 289 | 5 | 0 | 0 | G30X.00 | Acute transmural myocardial infarction of unspecif site | Feb-09 |
| Myocardial Infarction | 12229 | 46316 | 74 | 0 | 0 | G30X000 | Acute ST segment elevation myocardial infarction | Feb-09 |
| Myocardial Infarction | 34803 | 494 | 1 | 0 | 0 | G30y.00 | Other acute myocardial infarction | Feb-09 |
| Myocardial Infarction | 28736 | 177 | 1 | 0 | 0 | G30y000 | Acute atrial infarction | Feb-09 |
| Myocardial Infarction | 62626 | 5 | 0 | 0 | 0 | G30y100 | Acute papillary muscle infarction | Feb-09 |
| Myocardial Infarction | 41221 | 303 | 8 | 0 | 0 | G30y200 | Acute septal infarction | Feb-09 |
| Myocardial Infarction | 46017 | 1777 | 0 | 0 | 0 | G30yz00 | Other acute myocardial infarction NOS | Feb-09 |
| Myocardial Infarction | 14658 | 116170 | 6259 | 5 | 0 | G30z.00 | Acute myocardial infarction NOS | Feb-09 |
| Myocardial Infarction | 27951 | 329 | 5 | 0 | 0 | G31..00 | Other acute and subacute ischaemic heart disease | Feb-09 |
| Myocardial Infarction | 9413 | 139 | 4 | 0 | 0 | G31y.00 | Other acute and subacute ischaemic heart disease | Feb-09 |
| Myocardial Infarction | 27977 | 106 | 1 | 0 | 0 | G31yz00 | Other acute and subacute ischaemic heart disease NOS | Feb-09 |
| Myocardial Infarction | 23579 | 144 | 4 | 0 | 0 | G310.00 | Postmyocardial infarction syndrome | Feb-09 |
| Myocardial Infarction | 15661 | 728 | 18 | 0 | 0 | G310.11 | Dressler's syndrome | Feb-09 |
| Myocardial Infarction | 36523 | 7053 | 586 | 0 | 0 | G311.00 | Preinfarction syndrome | Feb-09 |
| Myocardial Infarction | 39655 | 5 | 0 | 0 | 0 | G311.12 | Impending infarction | Feb-09 |
| Myocardial Infarction | 61072 | 118 | 6 | 0 | 0 | G311000 | Myocardial infarction aborted | Feb-09 |
| Myocardial Infarction | 55137 | 84 | 0 | 0 | 0 | G311011 | MI - myocardial infarction aborted | Feb-09 |
| Myocardial Infarction | 11983 | 48767 | 264 | 0 | 0 | G311500 | Acute coronary syndrome | Feb-09 |
| Myocardial Infarction | 54251 | 164 | 2 | 0 | 0 | G311z00 | Preinfarction syndrome NOS | Feb-09 |
| Myocardial Infarction | 39449 | 137 | 0 | 0 | 0 | G312.00 | Coronary thrombosis not resulting in myocardial infarction | Feb-09 |
| Myocardial Infarction | 9276 | 3595 | 25 | 0 | 0 | G31y000 | Acute coronary insufficiency | Feb-09 |
| Myocardial Infarction | 68357 | 41 | 0 | 0 | 0 | G31y100 | Microinfarction of heart | Feb-09 |
| Myocardial Infarction | 39693 | 159 | 3 | 0 | 0 | G31y200 | Subendocardial ischaemia | Feb-09 |
| Myocardial Infarction | 21844 | 500 | 5 | 0 | 0 | G31y300 | Transient myocardial ischaemia | Feb-09 |
| Myocardial Infarction | 4017 | 14988 | 279 | 13 | 0 | G32..00 | Old myocardial infarction | Feb-09 |
| Myocardial Infarction | 16408 | 451 | 28 | 0 | 0 | G32..11 | Healed myocardial infarction | Feb-09 |
| Myocardial Infarction | 17464 | 1458 | 36 | 3 | 0 | G32..12 | Personal history of myocardial infarction | Feb-09 |
| Myocardial Infarction | 18842 | 500 | 9 | 0 | 0 | G35..00 | Subsequent myocardial infarction | Feb-09 |
| Myocardial Infarction | 45809 | 83 | 0 | 0 | 0 | G350.00 | Subsequent myocardial infarction of anterior wall | Feb-09 |
| Myocardial Infarction | 38609 | 93 | 1 | 0 | 0 | G351.00 | Subsequent myocardial infarction of inferior wall | Feb-09 |
| Myocardial Infarction | 72562 | 12 | 0 | 0 | 0 | G353.00 | Subsequent myocardial infarction of other sites | Feb-09 |
| Myocardial Infarction | 46166 | 25 | 0 | 0 | 0 | G35X.00 | Subsequent myocardial infarction of unspecified site | Feb-09 |
| Myocardial Infarction | 36423 | 35 | 1 | 0 | 0 | G36..00 | Certain current complication follow acute myocardial infarct | Feb-09 |
| Myocardial Infarction | 24126 | 172 | 1 | 0 | 0 | G360.00 | Haemopericardium/current comp folow acut myocard infarct | Feb-09 |
| Myocardial Infarction | 23708 | 55 | 2 | 0 | 0 | G361.00 | Atrial septal defect/curr comp folow acut myocardal infarct | Feb-09 |
| Myocardial Infarction | 37657 | 67 | 3 | 0 | 0 | G362.00 | Ventric septal defect/curr comp fol acut myocardal infarctn | Feb-09 |
| Myocardial Infarction | 59189 | 7 | 0 | 0 | 0 | G363.00 | Ruptur cardiac wall w'out haemopericard/cur comp fol ac MI | Feb-09 |
| Myocardial Infarction | 59940 | 2 | 0 | 0 | 0 | G364.00 | Ruptur chordae tendinae/curr comp fol acute myocard infarct | Feb-09 |
| Myocardial Infarction | 69474 | 6 | 0 | 0 | 0 | G365.00 | Rupture papillary muscle/curr comp fol acute myocard infarct | Feb-09 |
| Myocardial Infarction | 29553 | 70 | 3 | 0 | 0 | G366.00 | Thrombosis atrium,auric append&vent/curr comp foll acute MI | Feb-09 |
| Myocardial Infarction | 32272 | 318 | 7 | 0 | 0 | G38..00 | Postoperative myocardial infarction | Feb-09 |
| Myocardial Infarction | 46112 | 8 | 0 | 0 | 0 | G380.00 | Postoperative transmural myocardial infarction anterior wall | Feb-09 |
| Myocardial Infarction | 46276 | 51 | 0 | 0 | 0 | G381.00 | Postoperative transmural myocardial infarction inferior wall | Feb-09 |
| Myocardial Infarction | 106812 | 3 | 0 | 0 | 0 | G383.00 | Postoperative transmural myocardial infarction unspec site | Aug-13 |
| Myocardial Infarction | 41835 | 42 | 1 | 0 | 0 | G384.00 | Postoperative subendocardial myocardial infarction | Feb-09 |
| Myocardial Infarction | 68748 | 23 | 0 | 0 | 0 | G38z.00 | Postoperative myocardial infarction, unspecified | Feb-09 |
| Myocardial Infarction | 35119 | 96 | 1 | 0 | 0 | G501.00 | Post infarction pericarditis | Feb-09 |
| Myocardial Infarction | 96838 | 3 | 0 | 0 | 0 | Gyu3400 | [X]Acute transmural myocardial infarction of unspecif site | May-09 |
| Ischaemic Coronary Disease | 240 | 756631 | 11652 | 1653 | 0 | G3...00 | Ischaemic heart disease | Feb-09 |
| Ischaemic Coronary Disease | 24783 | 1082 | 1 | 0 | 0 | G3...11 | Arteriosclerotic heart disease | Feb-09 |
| Ischaemic Coronary Disease | 20416 | 2692 | 9 | 0 | 0 | G3...12 | Atherosclerotic heart disease | Feb-09 |
| Ischaemic Coronary Disease | 1792 | 187726 | 3305 | 10 | 0 | G3...13 | IHD - Ischaemic heart disease | Feb-09 |
| Ischaemic Coronary Disease | 28138 | 977 | 4 | 0 | 0 | G34..00 | Other chronic ischaemic heart disease | Feb-09 |
| Ischaemic Coronary Disease | 1676 | 104221 | 314 | 1 | 0 | G3z..00 | Ischaemic heart disease NOS | Feb-09 |
| Ischaemic Coronary Disease | 15754 | 456 | 13 | 0 | 0 | G34z.00 | Other chronic ischaemic heart disease NOS | Feb-09 |
| Ischaemic Coronary Disease | 5413 | 13313 | 104 | 2 | 0 | G340.00 | Coronary atherosclerosis | Feb-09 |
| Ischaemic Coronary Disease | 1655 | 14825 | 115 | 0 | 0 | G340.11 | Triple vessel disease of the heart | Feb-09 |
| Ischaemic Coronary Disease | 1344 | 64595 | 698 | 11 | 0 | G340.12 | Coronary artery disease | Feb-09 |
| Ischaemic Coronary Disease | 3999 | 8859 | 35 | 0 | 0 | G340000 | Single coronary vessel disease | Feb-09 |
| Ischaemic Coronary Disease | 5254 | 5514 | 21 | 0 | 0 | G340100 | Double coronary vessel disease | Feb-09 |
| Ischaemic Coronary Disease | 36609 | 470 | 1 | 0 | 0 | G342.00 | Atherosclerotic cardiovascular disease | Feb-09 |
| Ischaemic Coronary Disease | 7320 | 3719 | 43 | 0 | 0 | G343.00 | Ischaemic cardiomyopathy | Feb-09 |
| Ischaemic Coronary Disease | 34633 | 95 | 3 | 0 | 0 | G34y.00 | Other specified chronic ischaemic heart disease | Feb-09 |
| Ischaemic Coronary Disease | 24540 | 233 | 2 | 0 | 0 | G34y000 | Chronic coronary insufficiency | Feb-09 |
| Ischaemic Coronary Disease | 23078 | 2328 | 16 | 0 | 0 | G34y100 | Chronic myocardial ischaemia | Feb-09 |
| Ischaemic Coronary Disease | 35713 | 76 | 3 | 0 | 0 | G34yz00 | Other specified chronic ischaemic heart disease NOS | Feb-09 |
| Ischaemic Coronary Disease | 18889 | 1425 | 8 | 0 | 0 | G34z000 | Asymptomatic coronary heart disease | Feb-09 |
| Ischaemic Coronary Disease | 22383 | 1147 | 1 | 0 | 0 | G3y..00 | Other specified ischaemic heart disease | Feb-09 |
| Angina | 1431 | 40998 | 6441 | 3 | 0 | G311.13 | Unstable angina | Feb-09 |
| Angina | 7347 | 37182 | 1726 | 0 | 0 | G311100 | Unstable angina | Feb-09 |
| Angina | 19655 | 879 | 70 | 0 | 0 | G311.14 | Angina at rest | Feb-09 |
| Angina | 17307 | 2633 | 205 | 0 | 0 | G311200 | Angina at rest | Feb-09 |
| Angina | 34328 | 195 | 20 | 0 | 0 | G311300 | Refractory angina | Feb-09 |
| Angina | 18118 | 4243 | 605 | 0 | 0 | G311400 | Worsening angina | Feb-09 |
| Angina | 1430 | 1003830 | 42215 | 224 | 0 | G33..00 | Angina pectoris | Feb-09 |
| Angina | 20095 | 2546 | 207 | 0 | 0 | G330.00 | Angina decubitus | Feb-09 |
| Angina | 18125 | 430 | 21 | 0 | 0 | G330000 | Nocturnal angina | Feb-09 |
| Angina | 29902 | 65 | 1 | 0 | 0 | G330z00 | Angina decubitus NOS | Feb-09 |
| Angina | 12986 | 1207 | 22 | 0 | 0 | G331.00 | Prinzmetal's angina | Feb-09 |
| Angina | 11048 | 173 | 22 | 0 | 0 | G331.11 | Variant angina pectoris | Feb-09 |
| Angina | 36854 | 2113 | 10 | 0 | 0 | G332.00 | Coronary artery spasm | Feb-09 |
| Angina | 28554 | 25537 | 136 | 1 | 0 | G33zz00 | Angina pectoris NOS | Feb-09 |
| Angina | 25842 | 14921 | 83 | 0 | 0 | G33z.00 | Angina pectoris NOS | Feb-09 |
| Angina | 66388 | 69 | 0 | 0 | 0 | G33z000 | Status anginosus | Feb-09 |
| Angina | 7696 | 104 | 0 | 0 | 0 | G33z200 | Syncope anginosa | Feb-09 |
| Angina | 1414 | 8726 | 671 | 2 | 0 | G33z300 | Angina on effort | Feb-09 |
| Angina | 32450 | 2000 | 141 | 0 | 0 | G33z400 | Ischaemic chest pain | Feb-09 |
| Angina | 9555 | 320 | 2 | 0 | 0 | G33z500 | Post infarct angina | Feb-09 |
| Angina | 26863 | 940 | 8 | 0 | 0 | G33z600 | New onset angina | Feb-09 |
| Angina | 12804 | 12363 | 87 | 1 | 0 | G33z700 | Stable angina | Feb-09 |
| Stroke | 23671 | 2064 | 8 | 0 | 0 | G63y000 | Cerebral infarct due to thrombosis of precerebral arteries | Feb-09 |
| Stroke | 24446 | 539 | 5 | 0 | 0 | G63y100 | Cerebral infarction due to embolism of precerebral arteries | Feb-09 |
| Stroke | 8837 | 35835 | 209 | 0 | 0 | G64..00 | Cerebral arterial occlusion | Feb-09 |
| Stroke | 5363 | 41511 | 507 | 0 | 0 | G64..11 | CVA - cerebral artery occlusion | Feb-09 |
| Stroke | 569 | 5632 | 193 | 0 | 0 | G64..12 | Infarction - cerebral | Feb-09 |
| Stroke | 6155 | 7358 | 58 | 0 | 0 | G64..13 | Stroke due to cerebral arterial occlusion | Feb-09 |
| Stroke | 16517 | 4222 | 100 | 0 | 0 | G640.00 | Cerebral thrombosis | Feb-09 |
| Stroke | 36717 | 1108 | 12 | 0 | 0 | G640000 | Cerebral infarction due to thrombosis of cerebral arteries | Feb-09 |
| Stroke | 15019 | 1596 | 63 | 0 | 0 | G641.00 | Cerebral embolism | Feb-09 |
| Stroke | 34758 | 124 | 2 | 0 | 0 | G641.11 | Cerebral embolus | Feb-09 |
| Stroke | 3149 | 48851 | 249 | 0 | 0 | G64z.00 | Cerebral infarction NOS | Feb-09 |
| Stroke | 15252 | 445 | 30 | 0 | 0 | G64z.11 | Brainstem infarction NOS | Feb-09 |
| Stroke | 5602 | 12282 | 70 | 0 | 0 | G64z.12 | Cerebellar infarction | Feb-09 |
| Stroke | 25615 | 1032 | 39 | 0 | 0 | G64z000 | Brainstem infarction | Feb-09 |
| Stroke | 47642 | 79 | 1 | 0 | 0 | G64z100 | Wallenberg syndrome | Feb-09 |
| Stroke | 5185 | 433 | 5 | 0 | 0 | G64z111 | Lateral medullary syndrome | Feb-09 |
| Stroke | 9985 | 3934 | 8 | 0 | 0 | G64z200 | Left sided cerebral infarction | Feb-09 |
| Stroke | 10504 | 3552 | 4 | 0 | 0 | G64z300 | Right sided cerebral infarction | Feb-09 |
| Stroke | 26424 | 2380 | 8 | 0 | 0 | G64z400 | Infarction of basal ganglia | Feb-09 |
| Stroke | 3132 | 5632 | 1661 | 0 | 0 | G65..11 | Drop attack | Feb-09 |
| Stroke | 2417 | 9602 | 434 | 0 | 0 | G65..13 | Vertebro-basilar insufficiency | Feb-09 |
| Stroke | 23942 | 1338 | 42 | 0 | 0 | G650.00 | Basilar artery syndrome | Feb-09 |
| Stroke | 5268 | 4966 | 267 | 2 | 0 | G650.11 | Insufficiency - basilar artery | Feb-09 |
| Stroke | 23465 | 765 | 51 | 0 | 0 | G652.00 | Subclavian steal syndrome | Feb-09 |
| Stroke | 44765 | 131 | 2 | 0 | 0 | G653.00 | Carotid artery syndrome hemispheric | Feb-09 |
| Stroke | 50594 | 11 | 0 | 0 | 0 | G654.00 | Multiple and bilateral precerebral artery syndromes | Feb-09 |
| Stroke | 10794 | 2636 | 37 | 0 | 0 | G656.00 | Vertebrobasilar insufficiency | Feb-09 |
| Stroke | 55247 | 62 | 0 | 0 | 0 | G65z000 | Impending cerebral ischaemia | Feb-09 |
| Stroke | 1469 | 323475 | 24251 | 385 | 0 | G66..00 | Stroke and cerebrovascular accident unspecified | Feb-09 |
| Stroke | 1298 | 173353 | 3828 | 26 | 0 | G66..11 | CVA unspecified | Feb-09 |
| Stroke | 6253 | 14969 | 104 | 0 | 0 | G66..12 | Stroke unspecified | Feb-09 |
| Stroke | 6116 | 24841 | 454 | 0 | 0 | G66..13 | CVA - Cerebrovascular accident unspecified | Feb-09 |
| Stroke | 7780 | 9001 | 106 | 0 | 0 | G667.00 | Left sided CVA | Feb-09 |
| Stroke | 12833 | 7800 | 95 | 0 | 0 | G668.00 | Right sided CVA | Feb-09 |
| Stroke | 40758 | 823 | 2 | 0 | 0 | G6W..00 | Cereb infarct due unsp occlus/stenos precerebr arteries | Feb-09 |
| Stroke | 33543 | 3362 | 4 | 0 | 0 | G6X..00 | Cerebrl infarctn due/unspcf occlusn or sten/cerebrl artrs | Feb-09 |
| Stroke | 91627 | 127 | 0 | 0 | 0 | Gyu6300 | [X]Cerebrl infarctn due/unspcf occlusn or sten/cerebrl artrs | Feb-09 |
| Stroke | 53745 | 995 | 2 | 0 | 0 | Gyu6400 | [X]Other cerebral infarction | Feb-09 |
| Stroke | 90572 | 30 | 0 | 0 | 0 | Gyu6500 | [X]Occlusion and stenosis of other precerebral arteries | Feb-09 |
| Stroke | 92036 | 67 | 0 | 0 | 0 | Gyu6600 | [X]Occlusion and stenosis of other cerebral arteries | Feb-09 |
| Transient Ischaemic Attack | 63746 | 114 | 0 | 0 | 0 | Fyu5500 | [X]Other transnt cerebral ischaemic attacks+related syndroms | Feb-09 |
| Transient Ischaemic Attack | 504 | 235641 | 15990 | 8 | 0 | G65..00 | Transient cerebral ischaemia | Feb-09 |
| Transient Ischaemic Attack | 1433 | 243625 | 10700 | 0 | 0 | G65..12 | Transient ischaemic attack | Feb-09 |
| Transient Ischaemic Attack | 19354 | 444 | 4 | 0 | 0 | G65y.00 | Other transient cerebral ischaemia | Feb-09 |
| Transient Ischaemic Attack | 15788 | 21566 | 71 | 0 | 0 | G65zz00 | Transient cerebral ischaemia NOS | Feb-09 |
| Transient Ischaemic Attack | 1895 | 4458 | 75 | 0 | 0 | G65z.00 | Transient cerebral ischaemia NOS | Feb-09 |
|  | 16507 | 227 | 10 | 0 | 0 | G65z100 | Intermittent cerebral ischaemia | Feb-09 |
| Transient Ischaemic Attack | 101251 | 660 | 7 | 0 | 0 | ZV12D00 | [V]Personal history of transient ischaemic attack | Jan-11 |

| outcome | ICD-10 code |
| --- | --- |
| Myocardial Infarction | I21 |
| Myocardial Infarction | I22 |
| Myocardial Infarction | I23 |
| Myocardial Infarction | I24.1 |
| Myocardial Infarction | I25.2 |
| Ischaemic Coronary Disease | I24 |
| Ischaemic Coronary Disease | I25 |
| Stroke | I63 |
| Stroke | I69.3 |
| Stroke | G46.3 |
| Stroke | G46.4 |
| Stroke | G46.5 |
| Stroke | G46.6 |
| Stroke | G46.7 |
| Stroke | G46.8 |
| Transient Ischaemic Attack | I65 |
| Transient Ischaemic Attack | I66 |
| Transient Ischaemic Attack | G45 |
| Transient Ischaemic Attack | G46.0 |
| Transient Ischaemic Attack | G46.1 |
| Transient Ischaemic Attack | G46.2 |
