## Supplement 5 for "Increased risk of major ischaemic events among autistic people"

**Supplement 5 – Interaction Testing of Autism and Gender:**

|  | Model 1 | | Model 2 | | Model 3 | | Model 4 | |
| --- | --- | --- | --- | --- | --- | --- | --- | --- |
|  | *Hazard Ratio* | *P-Value* | *Hazard Ratio* | *P-Value* | *Hazard Ratio* | *P-Value* | *Hazard Ratio* | *P-Value* |
| Autism:Gender | 0.60 (0.43, 0.83) | 0.002 | 0.58 (0.41, 0.81) | 0.002 | 0.57 (0.41, 0.80) | 0.001 | 0.57 (0.39, 0.84) | 0.004 |
