## Supplementary figures and images for "Increased risk of major ischaemic events among autistic people"

### Supplement 1

**Supplement 1 – Methodology Flow Chart:**

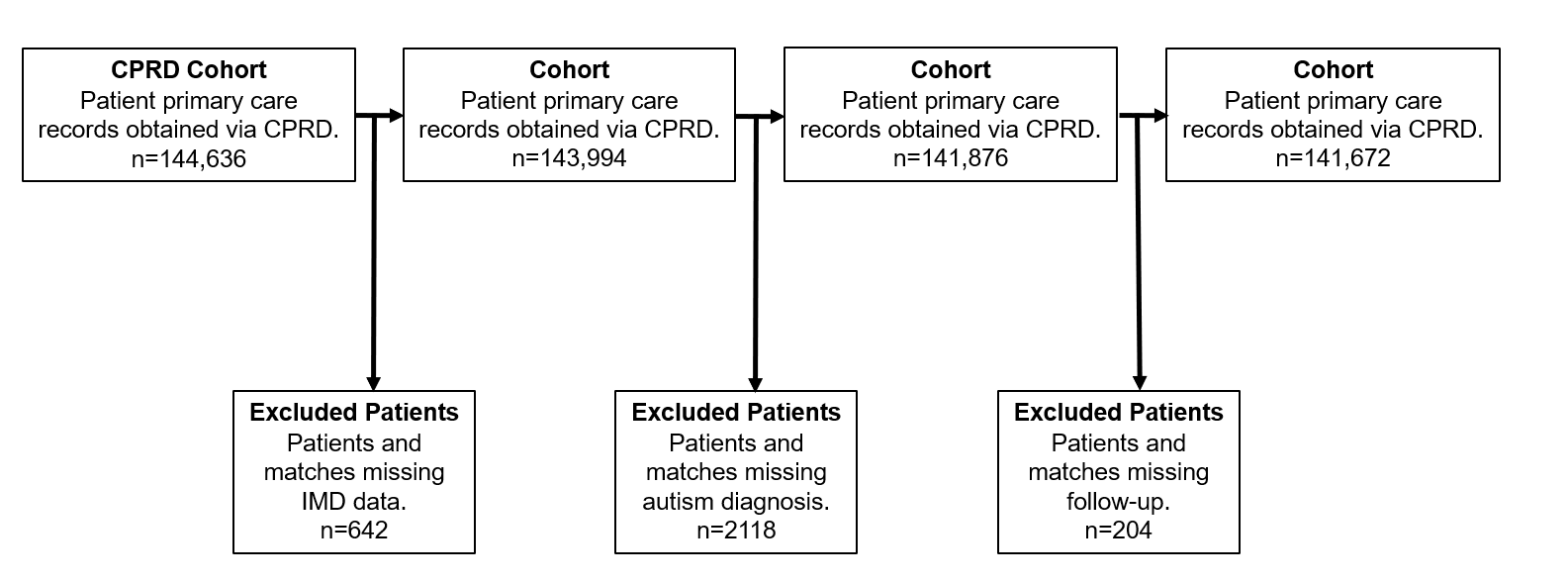
