## Supplement 4 for "Increased risk of major ischaemic events among autistic people"

**Supplement 4 – Co-Variate Medcode and ICD-10 Code List:**

| exposure | medcode | clinicalevents | referralevents | testevents | immunisationevents | readcode | readterm | databasebuild |
| --- | --- | --- | --- | --- | --- | --- | --- | --- |
| Atrial Fibrillation and Flutter | 90189 | 1046 | 0 | 0 | 0 | 9Os2.00 | Atrial fibrillation monitoring third letter | Feb-09 |
| Atrial Fibrillation and Flutter | 1664 | 595832 | 24353 | 103 | 0 | G573000 | Atrial fibrillation | Feb-09 |
| Atrial Fibrillation and Flutter | 96076 | 859 | 7 | 0 | 0 | G573500 | Persistent atrial fibrillation | Feb-09 |
| Atrial Fibrillation and Flutter | 90190 | 389 | 0 | 0 | 0 | 9Os3.00 | Atrial fibrillation monitoring verbal invite | Feb-09 |
| Atrial Fibrillation and Flutter | 28994 | 32159 | 101 | 0 | 0 | 212R.00 | Atrial fibrillation resolved | Feb-09 |
| Atrial Fibrillation and Flutter | 23437 | 2179 | 38 | 0 | 0 | G573z00 | Atrial fibrillation and flutter NOS | Feb-09 |
| Atrial Fibrillation and Flutter | 35127 | 259 | 0 | 0 | 0 | G573300 | Non-rheumatic atrial fibrillation | Feb-09 |
| Atrial Fibrillation and Flutter | 90188 | 3771 | 1 | 0 | 0 | 9Os1.00 | Atrial fibrillation monitoring second letter | Feb-09 |
| Atrial Fibrillation and Flutter | 90187 | 26611 | 5 | 0 | 0 | 9Os0.00 | Atrial fibrillation monitoring first letter | Feb-09 |
| Atrial Fibrillation and Flutter | 3757 | 10592 | 1239 | 122571 | 0 | 3272 | ECG: atrial fibrillation | Feb-09 |
| Atrial Fibrillation and Flutter | 57832 | 6903 | 157 | 0 | 0 | 9Os..00 | Atrial fibrillation monitoring administration | Feb-09 |
| Atrial Fibrillation and Flutter | 6345 | 23050 | 1316 | 8 | 0 | 14AN.00 | H/O: atrial fibrillation | Feb-09 |
| Atrial Fibrillation and Flutter | 90191 | 782 | 0 | 0 | 0 | 9Os4.00 | Atrial fibrillation monitoring telephone invite | Feb-09 |
| Atrial Fibrillation and Flutter | 45773 | 60839 | 56 | 0 | 0 | 6A9..00 | Atrial fibrillation annual review | Feb-09 |
| Atrial Fibrillation and Flutter | 63350 | 294 | 0 | 0 | 0 | 9hF..00 | Exception reporting: atrial fibrillation quality indicators | Feb-09 |
| Atrial Fibrillation and Flutter | 96277 | 489 | 3 | 0 | 0 | G573400 | Permanent atrial fibrillation | Apr-09 |
| Atrial Fibrillation and Flutter | 39114 | 6317 | 0 | 0 | 0 | 9hF1.00 | Excepted from atrial fibrillation qual indic: Inform dissent | Feb-09 |
| Atrial Fibrillation and Flutter | 18746 | 68310 | 1527 | 0 | 0 | 662S.00 | Atrial fibrillation monitoring | Feb-09 |
| Atrial Fibrillation and Flutter | 1268 | 99770 | 3304 | 3 | 0 | G573200 | Paroxysmal atrial fibrillation | Feb-09 |
| Atrial Fibrillation and Flutter | 2212 | 207816 | 14211 | 9 | 0 | G573.00 | Atrial fibrillation and flutter | Feb-09 |
| Atrial Fibrillation and Flutter | 9479 | 169 | 3 | 0 | 0 | 7936A00 | Implant intravenous pacemaker for atrial fibrillation | Feb-09 |
| Atrial Fibrillation and Flutter | 105554 | 1875 | 2 | 0 | 0 | 8CMW200 | Atrial fibrillation care pathway | Jan-13 |
| Chronic Kidney Disease | 95508 | 544 | 6 | 0 | 0 | 1Z1K.00 | Chronic kidney disease stage 5 with proteinuria | Feb-09 |
| Chronic Kidney Disease | 95177 | 6343 | 27 | 0 | 0 | 1Z1G.00 | Chronic kidney disease stage 3B without proteinuria | Feb-09 |
| Chronic Kidney Disease | 94965 | 36110 | 52 | 1 | 0 | 1Z15.00 | Chronic kidney disease stage 3A | Feb-09 |
| Chronic Kidney Disease | 95408 | 2933 | 23 | 0 | 0 | 1Z1D.00 | Chronic kidney disease stage 3A with proteinuria | Feb-09 |
| Chronic Kidney Disease | 95178 | 2224 | 24 | 0 | 0 | 1Z1F.00 | Chronic kidney disease stage 3B with proteinuria | Feb-09 |
| Chronic Kidney Disease | 95122 | 1954 | 71 | 0 | 0 | 1Z1H.00 | Chronic kidney disease stage 4 with proteinuria | Feb-09 |
| Chronic Kidney Disease | 95405 | 212 | 4 | 0 | 0 | 1Z1L.00 | Chronic kidney disease stage 5 without proteinuria | Feb-09 |
| Chronic Kidney Disease | 95406 | 2146 | 55 | 0 | 0 | 1Z1J.00 | Chronic kidney disease stage 4 without proteinuria | Feb-09 |
| Chronic Kidney Disease | 12479 | 73861 | 3169 | 0 | 0 | 1Z13.00 | Chronic kidney disease stage 4 | Feb-09 |
| Chronic Kidney Disease | 95175 | 17513 | 16 | 0 | 0 | 1Z1E.00 | Chronic kidney disease stage 3A without proteinuria | Feb-09 |
| Chronic Kidney Disease | 105151 | 735 | 5 | 0 | 0 | K055.00 | Chronic kidney disease stage 5 | Nov-12 |
| Chronic Kidney Disease | 12585 | 15680 | 252 | 0 | 0 | 1Z14.00 | Chronic kidney disease stage 5 | Feb-09 |
| Chronic Kidney Disease | 95179 | 13502 | 65 | 0 | 0 | 1Z16.00 | Chronic kidney disease stage 3B | Feb-09 |
| Chronic Kidney Disease | 104619 | 32383 | 46 | 0 | 0 | K053.00 | Chronic kidney disease stage 3 | Jul-12 |
| Chronic Kidney Disease | 12566 | 661565 | 2784 | 0 | 0 | 1Z12.00 | Chronic kidney disease stage 3 | Feb-09 |
| Chronic Kidney Disease | 94793 | 4407 | 29 | 0 | 0 | 1Z1B.00 | Chronic kidney disease stage 3 with proteinuria | Feb-09 |
| Chronic Kidney Disease | 95123 | 15911 | 10 | 0 | 0 | 1Z1C.00 | Chronic kidney disease stage 3 without proteinuria | Feb-09 |
| Chronic Kidney Disease | 104963 | 2645 | 20 | 0 | 0 | K054.00 | Chronic kidney disease stage 4 | Sep-12 |
| Chronic Kidney Disease | 17253 | 925 | 19 | 0 | 0 | 8L50.00 | Renal transplant planned | Feb-09 |
| Chronic Kidney Disease | 105811 | 67 | 0 | 0 | 0 | SP08R00 | Renal transplant rejection | Feb-13 |
| Chronic Kidney Disease | 18774 | 301 | 8 | 0 | 0 | TB00111 | Renal transplant with complication, without blame | Feb-09 |
| Chronic Kidney Disease | 107000 | 4 | 0 | 0 | 0 | SP08F00 | Acute rejection of renal transplant - grade II | Sep-13 |
| Chronic Kidney Disease | 105787 | 7 | 0 | 0 | 0 | 7B00600 | Xenograft renal transplant | Feb-13 |
| Chronic Kidney Disease | 105724 | 21 | 0 | 0 | 0 | SP08N00 | Unexplained episode of renal transplant dysfunction | Jan-13 |
| Chronic Kidney Disease | 104960 | 30 | 0 | 0 | 0 | SP08E00 | Acute rejection of renal transplant - grade I | Sep-12 |
| Chronic Kidney Disease | 114407 | 5 | 0 | 0 | 0 | SP08C00 | Accelerated rejection of renal transplant | Nov-19 |
| Chronic Kidney Disease | 106620 | 14 | 0 | 0 | 0 | SP08J00 | Chronic rejection of renal transplant | Aug-13 |
| Chronic Kidney Disease | 104905 | 6 | 0 | 0 | 0 | SP08D00 | Acute-on-chronic rejection of renal transplant | Sep-12 |
| Chronic Kidney Disease | 106866 | 6 | 0 | 0 | 0 | SP08W00 | Vascular complication of renal transplant | Aug-13 |
| Chronic Kidney Disease | 108437 | 8 | 0 | 0 | 0 | SP08V00 | Very mild acute rejection of renal transplant | Aug-14 |
| Chronic Kidney Disease | 104201 | 40 | 0 | 0 | 0 | SP08H00 | Acute rejection of renal transplant | Jun-12 |
| Chronic Kidney Disease | 26862 | 301 | 15 | 0 | 0 | 7B06300 | Exploration of renal transplant | Feb-09 |
| Chronic Kidney Disease | 107752 | 6 | 0 | 0 | 0 | SP08T00 | Urological complication of renal transplant | Mar-14 |
| Chronic Kidney Disease | 104630 | 2 | 0 | 0 | 0 | SP08G00 | Acute rejection of renal transplant - grade III | Jul-12 |
| Chronic Kidney Disease | 105328 | 271 | 0 | 0 | 0 | 7B00212 | Cadaveric renal transplant | Dec-12 |
| Chronic Kidney Disease | 53852 | 311 | 1 | 0 | 0 | K05..12 | End stage renal failure | Feb-09 |
| Chronic Kidney Disease | 6712 | 6089 | 70 | 1 | 0 | K050.00 | End stage renal failure | Feb-09 |
| Chronic Kidney Disease | 8330 | 762 | 5 | 0 | 0 | K0D..00 | End-stage renal disease | Feb-09 |
| Chronic Kidney Disease | 47672 | 97 | 3 | 0 | 0 | K01x400 | Nephrotic syndrome in systemic lupus erythematosus | Feb-09 |
| Chronic Kidney Disease | 9840 | 40 | 0 | 0 | 0 | K010.00 | Nephrotic syndrome with proliferative glomerulonephritis | Feb-09 |
| Chronic Kidney Disease | 23913 | 82 | 1 | 0 | 0 | K014.00 | Nephrotic syndrome, minor glomerular abnormality | Feb-09 |
| Chronic Kidney Disease | 27427 | 521 | 9 | 0 | 0 | K01z.00 | Nephrotic syndrome NOS | Feb-09 |
| Chronic Kidney Disease | 99644 | 4 | 0 | 0 | 0 | K012.00 | Nephrotic syndrome+membranoproliferative glomerulonephritis | Jun-10 |
| Chronic Kidney Disease | 19316 | 193 | 6 | 0 | 0 | K016.00 | Nephrotic syndrome, diffuse membranous glomerulonephritis | Feb-09 |
| Chronic Kidney Disease | 17365 | 69 | 0 | 0 | 0 | K01B.00 | Nephrotic syndrome, diffuse crescentic glomerulonephritis | Feb-09 |
| Chronic Kidney Disease | 56987 | 16 | 0 | 0 | 0 | K01A.00 | Nephrotic syndrome, dense deposit disease | Feb-09 |
| Chronic Kidney Disease | 58750 | 14 | 0 | 0 | 0 | K01x300 | Nephrotic syndrome in polyarteritis nodosa | Feb-09 |
| Chronic Kidney Disease | 47922 | 103 | 2 | 0 | 0 | K01x000 | Nephrotic syndrome in amyloidosis | Feb-09 |
| Chronic Kidney Disease | 2471 | 404 | 8 | 0 | 0 | K01x100 | Nephrotic syndrome in diabetes mellitus | Feb-09 |
| Chronic Kidney Disease | 99201 | 1 | 0 | 0 | 0 | K01x200 | Nephrotic syndrome in malaria | Apr-10 |
| Chronic Kidney Disease | 108816 | 2 | 0 | 0 | 0 | K01x.00 | Nephrotic syndrome in diseases EC | Nov-14 |
| Chronic Kidney Disease | 112548 | 1 | 0 | 0 | 0 | K01w111 | Nephrotic syndrome with pseudohermaphroditism | Apr-18 |
| Chronic Kidney Disease | 1803 | 673 | 9 | 0 | 0 | K011.00 | Nephrotic syndrome with membranous glomerulonephritis | Feb-09 |
| Chronic Kidney Disease | 94373 | 7 | 0 | 0 | 0 | K01y.00 | Nephrotic syndrome with other pathological kidney lesions | Feb-09 |
| Chronic Kidney Disease | 29634 | 244 | 4 | 0 | 0 | K013.00 | Nephrotic syndrome with minimal change glomerulonephritis | Feb-09 |
| Chronic Kidney Disease | 22852 | 323 | 24 | 0 | 0 | K015.00 | Nephrotic syndrome, focal and segmental glomerular lesions | Feb-09 |
| Chronic Kidney Disease | 2999 | 21031 | 980 | 5 | 0 | K01..00 | Nephrotic syndrome | Feb-09 |
| Chronic Kidney Disease | 57926 | 124 | 0 | 0 | 0 | K013.12 | Steroid sensitive nephrotic syndrome | Feb-09 |
| Chronic Kidney Disease | 63786 | 37 | 0 | 0 | 0 | K01w.00 | Congenital nephrotic syndrome | Feb-09 |
| Chronic Kidney Disease | 40349 | 9 | 0 | 0 | 0 | K013.11 | Lipoid nephrosis | Feb-09 |
| Chronic Kidney Disease | 72303 | 3 | 0 | 0 | 0 | K01w000 | Finnish nephrosis syndrome | Feb-09 |
| Chronic Kidney Disease | 45499 | 9 | 0 | 0 | 0 | K01x111 | Kimmelstiel - Wilson disease | Feb-09 |
| Chronic Kidney Disease | 7804 | 2979 | 81 | 0 | 0 | K02..00 | Chronic glomerulonephritis | Feb-09 |
| Chronic Kidney Disease | 34998 | 125 | 0 | 0 | 0 | K020.00 | Chronic proliferative glomerulonephritis | Feb-09 |
| Chronic Kidney Disease | 61494 | 52 | 0 | 0 | 0 | K022.00 | Chronic membranoproliferative glomerulonephritis | Feb-09 |
| Chronic Kidney Disease | 60857 | 26 | 0 | 0 | 0 | K0A3700 | Chronic nephritic syn diffuse crescentic glomerulonephritis | Feb-09 |
| Chronic Kidney Disease | 10809 | 527 | 9 | 0 | 0 | K021.00 | Chronic membranous glomerulonephritis | Feb-09 |
| Chronic Kidney Disease | 65064 | 5 | 0 | 0 | 0 | K023.00 | Chronic rapidly progressive glomerulonephritis | Feb-09 |
| Chronic Kidney Disease | 65400 | 8 | 0 | 0 | 0 | K02y300 | Chronic diffuse glomerulonephritis | Feb-09 |
| Chronic Kidney Disease | 73026 | 8 | 0 | 0 | 0 | K0A3500 | Chronic neph syn difus mesangiocapillary glomerulonephritis | Feb-09 |
| Chronic Kidney Disease | 56893 | 16 | 0 | 0 | 0 | K0A3300 | Chron neph syn difus mesangial prolifrtiv glomerulonephritis | Feb-09 |
| Chronic Kidney Disease | 15097 | 261 | 7 | 0 | 0 | K02z.00 | Chronic glomerulonephritis NOS | Feb-09 |
| Chronic Kidney Disease | 57168 | 19 | 0 | 0 | 0 | K0A3200 | Chron nephritic syndrom difuse membranous glomerulonephritis | Feb-09 |
| Chronic Kidney Disease | 60960 | 18 | 0 | 0 | 0 | K02y.00 | Other chronic glomerulonephritis | Feb-09 |
| Chronic Kidney Disease | 21947 | 173 | 1 | 0 | 0 | K017.00 | Nephrotic syn difus mesangial prolifertiv glomerulonephritis | Feb-09 |
| Chronic Kidney Disease | 63615 | 20 | 0 | 0 | 0 | K02yz00 | Other chronic glomerulonephritis NOS | Feb-09 |
| Chronic Kidney Disease | 97758 | 3 | 0 | 0 | 0 | K02y000 | Chronic glomerulonephritis + diseases EC | Oct-09 |
| Chronic Kidney Disease | 4669 | 232 | 7 | 0 | 0 | K02y200 | Chronic focal glomerulonephritis | Feb-09 |
| Chronic Kidney Disease | 10647 | 516 | 10 | 0 | 0 | K02..11 | Nephritis - chronic | Feb-09 |
| Chronic Kidney Disease | 512 | 79754 | 4893 | 14 | 0 | K05..00 | Chronic renal failure | Feb-09 |
| Chronic Kidney Disease | 4654 | 6848 | 127 | 0 | 0 | K100.00 | Chronic pyelonephritis | Feb-09 |
| Chronic Kidney Disease | 57568 | 6 | 1 | 0 | 0 | K100100 | Chronic pyelonephritis with medullary necrosis | Feb-09 |
| Chronic Kidney Disease | 99631 | 4 | 0 | 0 | 0 | K100000 | Chronic pyelonephritis without medullary necrosis | Jun-10 |
| Chronic Kidney Disease | 48855 | 40 | 0 | 0 | 0 | K100500 | Chronic obstructive pyelonephritis | Feb-09 |
| Chronic Kidney Disease | 48111 | 329 | 1 | 0 | 0 | K100z00 | Chronic pyelonephritis NOS | Feb-09 |
| Chronic Kidney Disease | 2939 | 413 | 8 | 0 | 0 | K100600 | Calculous pyelonephritis | Feb-09 |
| Chronic Kidney Disease | 21158 | 294 | 5 | 0 | 0 | K100200 | Chronic pyelitis | Feb-09 |
| Chronic Kidney Disease | 5911 | 3064 | 32 | 0 | 0 | ZV42000 | [V]Kidney transplanted | Feb-09 |
| Chronic Kidney Disease | 54990 | 96 | 2 | 0 | 0 | TB00100 | Kidney transplant with complication, without blame | Feb-09 |
| Chronic Kidney Disease | 11553 | 506 | 9 | 0 | 0 | SP08300 | Kidney transplant failure and rejection | Feb-09 |
| Erectile Dysfunction | 17894 | 2308 | 31 | 0 | 0 | K27y100 | Impotence of organic origin | Feb-09 |
| Erectile Dysfunction | 40725 | 22 | 1 | 0 | 0 | ZG43600 | Advice on technique for impotence | Feb-09 |
| Erectile Dysfunction | 94343 | 2568 | 30 | 0 | 0 | 1ABC.00 | Cannot sustain an erection | Feb-09 |
| Erectile Dysfunction | 81439 | 238 | 45 | 0 | 0 | 7C25F00 | Operations on penis for erectile dysfunction NEC | Feb-09 |
| Erectile Dysfunction | 92310 | 7 | 0 | 0 | 0 | Z9E9.00 | Provision of device for impotence | Feb-09 |
| Erectile Dysfunction | 12066 | 6609 | 296 | 0 | 0 | Eu52212 | [X]Male erectile disorder | Feb-09 |
| Erectile Dysfunction | 17639 | 1364 | 19 | 0 | 0 | Eu52213 | [X]Psychogenic impotence | Feb-09 |
| Erectile Dysfunction | 33494 | 372 | 6 | 0 | 0 | Eu52200 | [X]Failure of genital response | Feb-09 |
| Erectile Dysfunction | 3838 | 560134 | 17423 | 262 | 0 | E227311 | Erectile dysfunction | Feb-09 |
| Erectile Dysfunction | 710 | 212166 | 14589 | 34 | 0 | E227300 | Impotence | Feb-09 |
| Erectile Dysfunction | 37391 | 40 | 4 | 0 | 0 | 7A6G000 | Revascularisation for impotence | Feb-09 |
| Erectile Dysfunction | 12867 | 4347 | 41 | 0 | 0 | 7C25E00 | Treatment of erectile dysfunction NEC | Feb-09 |
| Erectile Dysfunction | 102274 | 139127 | 908 | 0 | 0 | 1D1B.00 | C/O erectile dysfunction | May-11 |
| Erectile Dysfunction | 20833 | 255 | 22 | 0 | 0 | 7C25B00 | Penile injection to produce erection | Feb-09 |
| Erectile Dysfunction | 94821 | 3226 | 75 | 0 | 0 | 1ABB.00 | Cannot get an erection | Feb-09 |
| Erectile Dysfunction | 103966 | 26 | 2 | 0 | 0 | 8BB4.00 | Erect dysf unresponsiv to phosphodiesterase-5 inhibitor | Apr-12 |
| Erectile Dysfunction | 12790 | 2269 | 3809 | 0 | 0 | 8HTj.00 | Referral to erectile dysfunction clinic | Feb-09 |
| Erectile Dysfunction | 41382 | 63 | 2 | 0 | 0 | 7A6G500 | Ligation of penile veins for impotence | Feb-09 |
| Erectile Dysfunction | 106360 | 534 | 2 | 0 | 0 | K27y700 | Erectile dysfunction due to diabetes mellitus | Jun-13 |
| Family History of Young Myocardial Infarcion and Angina | 30789 | 4523 | 2 | 0 | 0 | 12CL.00 | FH: Angina in 1st degree female relative <65 years | Feb-09 |
| Family History of Young Myocardial Infarcion and Angina | 26653 | 6141 | 0 | 0 | 0 | 12CM.00 | FH: Angina in 1st degree male relative <55 years | Feb-09 |
| Family History of Young Myocardial Infarcion and Angina | 18661 | 12146 | 3 | 0 | 0 | 12CP.00 | FH: Myocardial infarct in 1st degree male relative <55 years | Feb-09 |
| Family History of Young Myocardial Infarcion and Angina | 19127 | 5264 | 1 | 0 | 0 | 12CN.00 | FH: Myocardial infarct in 1st degree female relative <65 yrs | Feb-09 |
| Hypertension | 16292 | 4743 | 61 | 1 | 0 | G21..00 | Hypertensive heart disease | Feb-09 |
| Hypertension | 72030 | 4 | 0 | 0 | 0 | L122100 | Other pre-existing hypertension in preg/childb/puerp - deliv | Feb-09 |
| Hypertension | 34744 | 208 | 2 | 0 | 0 | G244.00 | Hypertension secondary to endocrine disorders | Feb-09 |
| Hypertension | 204 | 1865402 | 23171 | 2209 | 0 | G2...00 | Hypertensive disease | Feb-09 |
| Hypertension | 8732 | 45512 | 242 | 0 | 0 | G2...11 | BP - hypertensive disease | Feb-09 |
| Hypertension | 44549 | 29 | 1 | 0 | 0 | L128.00 | Pre-exist hypertension compl preg childbirth and puerperium | Feb-09 |
| Hypertension | 63000 | 7 | 0 | 0 | 0 | G231.00 | Benign hypertensive heart and renal disease | Feb-09 |
| Hypertension | 25371 | 184 | 1 | 0 | 0 | G241000 | Secondary benign renovascular hypertension | Feb-09 |
| Hypertension | 57288 | 110 | 1 | 0 | 0 | G241.00 | Secondary benign hypertension | Feb-09 |
| Hypertension | 61166 | 108 | 1 | 0 | 0 | G21z000 | Hypertensive heart disease NOS without CCF | Feb-09 |
| Hypertension | 93055 | 7 | 0 | 0 | 0 | L127z00 | Pre-eclampsia or eclampsia + pre-existing hypertension NOS | Feb-09 |
| Hypertension | 50157 | 45 | 0 | 0 | 0 | G210.00 | Malignant hypertensive heart disease | Feb-09 |
| Hypertension | 63164 | 78 | 0 | 0 | 0 | U60C500 | [X]Oth antihyperten drug caus advers eff in therap use, NEC | Feb-09 |
| Hypertension | 2666 | 202145 | 2767 | 16 | 0 | 14A2.00 | H/O: hypertension | Feb-09 |
| Hypertension | 16059 | 582 | 26 | 0 | 0 | G24z.00 | Secondary hypertension NOS | Feb-09 |
| Hypertension | 15106 | 401 | 13 | 0 | 0 | G22z.00 | Hypertensive renal disease NOS | Feb-09 |
| Hypertension | 43935 | 67 | 0 | 0 | 0 | G221.00 | Benign hypertensive renal disease | Feb-09 |
| Hypertension | 96743 | 1 | 0 | 0 | 0 | L122300 | Other pre-exist hypertension in preg/childb/puerp-not deliv | Apr-09 |
| Hypertension | 62718 | 112 | 0 | 0 | 0 | G21z100 | Hypertensive heart disease NOS with CCF | Feb-09 |
| Hypertension | 52127 | 94 | 0 | 0 | 0 | G211100 | Benign hypertensive heart disease with CCF | Feb-09 |
| Hypertension | 10818 | 271215 | 2695 | 9 | 0 | G20z.00 | Essential hypertension NOS | Feb-09 |
| Hypertension | 68659 | 10 | 0 | 0 | 0 | G23z.00 | Hypertensive heart and renal disease NOS | Feb-09 |
| Hypertension | 52621 | 39 | 4 | 0 | 0 | L128200 | Pre-exist 2ndry hypertens comp preg childbth and puerperium | Feb-09 |
| Hypertension | 67232 | 16 | 0 | 0 | 0 | G230.00 | Malignant hypertensive heart and renal disease | Feb-09 |
| Hypertension | 66567 | 18 | 0 | 0 | 0 | L122.00 | Other pre-existing hypertension in preg/childbirth/puerp | Feb-09 |
| Hypertension | 6702 | 7059 | 282 | 4 | 0 | F421300 | Hypertensive retinopathy | Feb-09 |
| Hypertension | 51635 | 95 | 0 | 0 | 0 | G241z00 | Secondary benign hypertension NOS | Feb-09 |
| Hypertension | 29310 | 637 | 26 | 1 | 0 | G22z.11 | Renal hypertension | Feb-09 |
| Hypertension | 3979 | 837 | 42 | 9 | 0 | G672.00 | Hypertensive encephalopathy | Feb-09 |
| Hypertension | 11056 | 183085 | 42 | 0 | 0 | 8BL0.00 | Patient on maximal tolerated antihypertensive therapy | Feb-09 |
| Hypertension | 16173 | 558 | 28 | 0 | 0 | G21zz00 | Hypertensive heart disease NOS | Feb-09 |
| Hypertension | 52427 | 65 | 0 | 0 | 0 | G211.00 | Benign hypertensive heart disease | Feb-09 |
| Hypertension | 13188 | 38174 | 38 | 0 | 0 | 662G.00 | Hypertensive treatm.changed | Feb-09 |
| Hypertension | 61660 | 21 | 0 | 0 | 0 | G211000 | Benign hypertensive heart disease without CCF | Feb-09 |
| Hypertension | 31387 | 298 | 11 | 0 | 0 | G24z000 | Secondary renovascular hypertension NOS | Feb-09 |
| Hypertension | 85944 | 4 | 0 | 0 | 0 | 7Q01.00 | High cost hypertension drugs | Feb-09 |
| Hypertension | 22333 | 511 | 5 | 0 | 0 | 8I3N.00 | Hypertension treatment refused | Feb-09 |
| Hypertension | 31341 | 384 | 1 | 0 | 0 | G24z100 | Hypertension secondary to drug | Feb-09 |
| Hypertension | 3425 | 54939 | 800 | 1 | 0 | 662O.00 | On treatment for hypertension | Feb-09 |
| Hypertension | 7057 | 68340 | 229 | 0 | 0 | G2z..00 | Hypertensive disease NOS | Feb-09 |
| Hypertension | 18590 | 4944 | 19 | 0 | 0 | 662b.00 | Moderate hypertension control | Feb-09 |
| Hypertension | 28684 | 57 | 3 | 0 | 0 | G233.00 | Hypertensive heart and renal disease with renal failure | Feb-09 |
| Hypertension | 18057 | 7213 | 1 | 0 | 0 | 8B26.00 | Antihypertensive therapy | Feb-09 |
| Hypertension | 19070 | 1048298 | 90 | 0 | 0 | 662d.00 | Hypertension annual review | Feb-09 |
| Hypertension | 57987 | 10 | 1 | 0 | 0 | G234.00 | Hyperten heart&renal dis+both(congestv)heart and renal fail | Feb-09 |
| Hypertension | 21837 | 35 | 4 | 0 | 0 | G232.00 | Hypertensive heart&renal dis wth (congestive) heart failure | Feb-09 |
| Hypertension | 16565 | 91281 | 33 | 0 | 0 | 6627 | Good hypertension control | Feb-09 |
| Hypertension | 66645 | 282 | 0 | 0 | 0 | 9OI9.00 | Hypertens.monitor deleted | Feb-09 |
| Hypertension | 21826 | 9391 | 23 | 0 | 0 | 662F.00 | Hypertension treatm. started | Feb-09 |
| Hypertension | 37086 | 113 | 2 | 0 | 0 | F404200 | Blind hypertensive eye | Feb-09 |
| Hypertension | 43664 | 107 | 1 | 0 | 0 | L127.00 | Pre-eclampsia or eclampsia with pre-existing hypertension | Feb-09 |
| Hypertension | 32976 | 388 | 0 | 0 | 0 | 6146200 | Hypertension induced by oral contraceptive pill | Feb-09 |
| Hypertension | 20497 | 1421 | 2 | 0 | 0 | TJC7z00 | Adverse reaction to antihypertensives NOS | Feb-09 |
| Hypertension | 1894 | 59893 | 116 | 0 | 0 | G201.00 | Benign essential hypertension | Feb-09 |
| Hypertension | 44350 | 117 | 0 | 0 | 0 | U60C51A | [X] Adverse reaction to antihypertensives NOS | Feb-09 |
| Hypertension | 63466 | 76 | 0 | 0 | 0 | G23..00 | Hypertensive heart and renal disease | Feb-09 |
| Hypertension | 3712 | 98760 | 722 | 7 | 0 | G20z.11 | Hypertension NOS | Feb-09 |
| Hypertension | 31755 | 126 | 7 | 0 | 0 | G240.00 | Secondary malignant hypertension | Feb-09 |
| Hypertension | 39649 | 157 | 11 | 0 | 0 | G220.00 | Malignant hypertensive renal disease | Feb-09 |
| Hypertension | 62432 | 18 | 1 | 0 | 0 | L122z00 | Other pre-existing hypertension in preg/childb/puerp NOS | Feb-09 |
| Hypertension | 8857 | 1794 | 272 | 1 | 0 | G21z011 | Cardiomegaly - hypertensive | Feb-09 |
| Hypertension | 59383 | 27 | 1 | 0 | 0 | G240000 | Secondary malignant renovascular hypertension | Feb-09 |
| Hypertension | 31464 | 161 | 2 | 0 | 0 | G21z.00 | Hypertensive heart disease NOS | Feb-09 |
| Hypertension | 72668 | 7 | 0 | 0 | 0 | G210100 | Malignant hypertensive heart disease with CCF | Feb-09 |
| Hypertension | 31816 | 359 | 14 | 0 | 0 | G672.11 | Hypertensive crisis | Feb-09 |
| Hypertension | 18765 | 1994 | 8 | 0 | 0 | G2y..00 | Other specified hypertensive disease | Feb-09 |
| Hypertension | 7329 | 4478 | 26 | 0 | 0 | G24..00 | Secondary hypertension | Feb-09 |
| Hypertension | 3269 | 30335 | 62 | 0 | 0 | 2126100 | Hypertension resolved | Feb-09 |
| Hypertension | 15377 | 5074 | 467 | 0 | 0 | G200.00 | Malignant essential hypertension | Feb-09 |
| Hypertension | 18482 | 189034 | 173 | 0 | 0 | 662c.00 | Hypertension six month review | Feb-09 |
| Hypertension | 95334 | 5 | 0 | 0 | 0 | G210000 | Malignant hypertensive heart disease without CCF | Feb-09 |
| Hypertension | 19342 | 10981 | 1 | 0 | 0 | 212K.00 | Hypertension resolved | Feb-09 |
| Hypertension | 21660 | 1223 | 1 | 0 | 0 | TJC7.00 | Adverse reaction to other antihypertensives | Feb-09 |
| Hypertension | 32423 | 109 | 0 | 0 | 0 | G222.00 | Hypertensive renal disease with renal failure | Feb-09 |
| Hypertension | 69753 | 240 | 1 | 0 | 0 | Gyu2.00 | [X]Hypertensive diseases | Feb-09 |
| Hypertension | 95359 | 7 | 0 | 0 | 0 | 662r.00 | Trial withdrawal of antihypertensive therapy | Feb-09 |
| Hypertension | 799 | 4197252 | 17507 | 38 | 0 | G20..00 | Essential hypertension | Feb-09 |
| Hypertension | 73293 | 9 | 0 | 0 | 0 | G240z00 | Secondary malignant hypertension NOS | Feb-09 |
| Hypertension | 30770 | 83 | 0 | 0 | 0 | U60C511 | [X] Adverse reaction to other antihypertensives | Feb-09 |
| Hypertension | 4372 | 21286 | 216 | 4 | 0 | G202.00 | Systolic hypertension | Feb-09 |
| Hypertension | 83473 | 1074 | 15 | 0 | 0 | G203.00 | Diastolic hypertension | Feb-09 |
| Hypertension | 4668 | 2554 | 63 | 0 | 0 | G22..00 | Hypertensive renal disease | Feb-09 |
| Hypertension | 42229 | 340 | 0 | 0 | 0 | G24zz00 | Secondary hypertension NOS | Feb-09 |
| Hypertension | 97533 | 10 | 0 | 0 | 0 | Gyu2100 | [X]Hypertension secondary to other renal disorders | Aug-09 |
| Hypertension | 8296 | 7024 | 30 | 0 | 0 | 6624 | Borderline hyperten:yearly obs | Feb-09 |
| Hypertension | 27511 | 53138 | 128 | 1 | 0 | 6628 | Poor hypertension control | Feb-09 |
| Hypertension | 73586 | 11 | 0 | 0 | 0 | L122000 | Other pre-existing hypertension in preg/childb/puerp unspec | Feb-09 |
| Hypertension | 60655 | 1 | 2 | 0 | 0 | L128000 | Pre-exist hyperten heart dis compl preg childbth+puerperium | Feb-09 |
| Hypertension | 351 | 262754 | 7147 | 336 | 0 | G20..11 | High blood pressure | Feb-09 |
| Migraine | 27930 | 962 | 6 | 0 | 0 | F26y300 | Complicated migraine | Feb-09 |
| Migraine | 2861 | 12429 | 228 | 0 | 0 | F262200 | Abdominal migraine | Feb-09 |
| Migraine | 103451 | 3090 | 21 | 0 | 0 | 1474000 | H/O migraine with aura | Dec-11 |
| Migraine | 5029 | 84605 | 852 | 0 | 0 | 1474 | H/O: migraine | Feb-09 |
| Migraine | 11138 | 9182 | 72 | 0 | 0 | K584.11 | Migraine - menstrual | Feb-09 |
| Migraine | 65262 | 14 | 1 | 0 | 0 | F26y111 | Moebius' ophthalmoplegic migraine | Feb-09 |
| Migraine | 103602 | 912 | 0 | 1 | 0 | F261.11 | Migraine without aura | Jan-12 |
| Migraine | 9004 | 1898 | 37 | 0 | 0 | F262300 | Basilar migraine | Feb-09 |
| Migraine | 41497 | 6055 | 1 | 0 | 0 | F261z00 | Common migraine NOS | Feb-09 |
| Migraine | 23621 | 1064 | 23 | 0 | 0 | F262z00 | Migraine variant NOS | Feb-09 |
| Migraine | 2424 | 8772 | 25 | 0 | 0 | F261.00 | Common migraine | Feb-09 |
| Migraine | 14700 | 38585 | 284 | 1 | 0 | F26z.00 | Migraine NOS | Feb-09 |
| Migraine | 5509 | 1116 | 28 | 0 | 0 | F262.00 | Migraine variants | Feb-09 |
| Migraine | 22685 | 411 | 3 | 0 | 0 | F26y200 | Status migrainosus | Feb-09 |
| Migraine | 28092 | 857 | 4 | 0 | 0 | F26yz00 | Other forms of migraine NOS | Feb-09 |
| Migraine | 53813 | 237 | 5 | 0 | 0 | Fyu5300 | [X]Other migraine | Feb-09 |
| Migraine | 11389 | 17275 | 74 | 0 | 0 | 8B6N.00 | Migraine prophylaxis | Feb-09 |
| Migraine | 3220 | 27350 | 111 | 0 | 0 | F260.00 | Classical migraine | Feb-09 |
| Migraine | 103973 | 39 | 0 | 0 | 0 | F262800 | Migraine induced by oestrogen contraceptive | May-12 |
| Migraine | 10583 | 3466 | 63 | 0 | 0 | F262400 | Ophthalmic migraine | Feb-09 |
| Migraine | 9633 | 3438 | 68 | 0 | 0 | F261000 | Atypical migraine | Feb-09 |
| Migraine | 161 | 1621622 | 21733 | 34 | 0 | F26..00 | Migraine | Feb-09 |
| Migraine | 103502 | 4994 | 18 | 0 | 0 | F260.11 | Migraine with aura | Jan-12 |
| Migraine | 28031 | 812 | 6 | 0 | 0 | F26y.00 | Other forms of migraine | Feb-09 |
| Migraine | 3658 | 9779 | 415 | 0 | 0 | F26y000 | Hemiplegic migraine | Feb-09 |
| Migraine | 12511 | 1758 | 37 | 0 | 0 | F26y100 | Ophthalmoplegic migraine | Feb-09 |
| Migraine | 17762 | 1030 | 10 | 0 | 0 | R090D00 | [D]Abdominal migraine | Feb-09 |
| Migraine | 6433 | 1966 | 19 | 0 | 0 | 1967 | Abdominal migraine - symptom | Feb-09 |
| Obesity | 104421 | 2 | 0 | 0 | 0 | C380700 | Lifelong obesity | Jun-12 |
| Obesity | 25968 | 304 | 2 | 0 | 0 | C380500 | Generalised obesity | Feb-09 |
| Obesity | 56107 | 10 | 0 | 0 | 0 | 7633000 | Bypass of jejunum by anastomosis of jejunum to jejunum | Feb-09 |
| Obesity | 24755 | 181 | 15 | 0 | 0 | C38y.11 | Pickwickian syndrome | Feb-09 |
| Obesity | 22556 | 74286 | 300 | 5 | 0 | 22K7.00 | Body mass index 40+ - severely obese | Feb-09 |
| Obesity | 64712 | 59 | 0 | 0 | 0 | 66C5.00 | Treatment of obesity changed | Feb-09 |
| Obesity | 64123 | 5 | 0 | 0 | 0 | 7633y00 | Other specified bypass of jejunum | Feb-09 |
| Obesity | 55586 | 113 | 0 | 0 | 0 | 9OK5.00 | Obesity monitoring 2nd letter | Feb-09 |
| Obesity | 10728 | 4325 | 129 | 0 | 0 | ZC2CM00 | Dietary advice for obesity | Feb-09 |
| Obesity | 67516 | 455 | 0 | 0 | 0 | 9OK2.00 | Refuses obesity monitoring | Feb-09 |
| Obesity | 52034 | 2288 | 1 | 0 | 0 | 9OK1.00 | Attends obesity monitoring | Feb-09 |
| Obesity | 32843 | 1778 | 32 | 0 | 0 | 9OK..00 | Obesity monitoring admin. | Feb-09 |
| Obesity | 67517 | 165 | 0 | 0 | 0 | 9OK8.00 | Obesity monitor phone invite | Feb-09 |
| Obesity | 38799 | 1255 | 2 | 0 | 0 | C380000 | Obesity due to excess calories | Feb-09 |
| Obesity | 49250 | 47 | 0 | 0 | 0 | C380100 | Drug-induced obesity | Feb-09 |
| Obesity | 38658 | 7373 | 9 | 0 | 0 | 66C1.00 | Initial obesity assessment | Feb-09 |
| Obesity | 40153 | 4620 | 15 | 0 | 0 | 66CZ.00 | Obesity monitoring NOS | Feb-09 |
| Obesity | 11401 | 1426 | 31 | 0 | 0 | C38z000 | Simple obesity NOS | Feb-09 |
| Obesity | 8854 | 20725 | 765 | 0 | 0 | C380300 | Morbid obesity | Feb-09 |
| Obesity | 52735 | 1587 | 1 | 0 | 0 | 9OKZ.00 | Obesity monitoring admin.NOS | Feb-09 |
| Obesity | 90600 | 10 | 0 | 0 | 0 | 7613500 | Partitioning of stomach NEC | Feb-09 |
| Obesity | 29538 | 13356 | 3 | 0 | 0 | 66C2.00 | Follow-up obesity assessment | Feb-09 |
| Obesity | 47439 | 11677 | 5 | 0 | 0 | 9OKA.00 | Obesity monitoring check done | Feb-09 |
| Obesity | 38059 | 362 | 3 | 0 | 0 | C380200 | Extreme obesity with alveolar hypoventilation | Feb-09 |
| Obesity | 70950 | 198 | 0 | 0 | 0 | 9OK7.00 | Obesity monitoring verbal inv. | Feb-09 |
| Obesity | 7984 | 27994 | 905 | 2 | 0 | 22A5.11 | O/E - obese | Feb-09 |
| Obesity | 52782 | 2 | 0 | 0 | 0 | Cyu7.00 | [X]Obesity and other hyperalimentation | Feb-09 |
| Obesity | 11461 | 177988 | 971 | 0 | 0 | 66C..00 | Obesity monitoring | Feb-09 |
| Obesity | 38632 | 1758 | 0 | 0 | 0 | 66C6.00 | Treatment of obesity started | Feb-09 |
| Obesity | 21744 | 192 | 35 | 0 | 0 | 9OK..11 | Obesity clinic administration | Feb-09 |
| Obesity | 22695 | 2070 | 8 | 0 | 0 | C380400 | Central obesity | Feb-09 |
| Obesity | 17477 | 1464 | 32 | 0 | 0 | ZV65319 | [V]Dietary counselling in obesity | Feb-09 |
| Obesity | 88474 | 816 | 18 | 0 | 0 | 7613300 | Partitioning of stomach using band | Feb-09 |
| Obesity | 52703 | 324 | 1 | 0 | 0 | 212Q.00 | Obesity resolved | Feb-09 |
| Obesity | 70898 | 22 | 0 | 0 | 0 | C38z.00 | Obesity and other hyperalimentation NOS | Feb-09 |
| Obesity | 3176 | 9431 | 1060 | 4 | 0 | 66C4.00 | Has seen dietician - obesity | Feb-09 |
| Obesity | 69757 | 25 | 1 | 0 | 0 | Cyu7000 | [X]Other obesity | Feb-09 |
| Obesity | 40977 | 78 | 0 | 0 | 0 | 7614100 | Bypass of stomach by anastomosis of stomach to duodenum | Feb-09 |
| Obesity | 16196 | 7311 | 225 | 3 | 0 | 1444 | H/O: obesity | Feb-09 |
| Obesity | 49409 | 2730 | 4 | 0 | 0 | 9OK4.00 | Obesity monitoring 1st letter | Feb-09 |
| Obesity | 55585 | 35 | 0 | 0 | 0 | 9OK6.00 | Obesity monitoring 3rd letter | Feb-09 |
| Obesity | 52036 | 250 | 0 | 0 | 0 | 9OK3.00 | Obesity monitoring default | Feb-09 |
| Obesity | 430 | 602272 | 27338 | 125 | 0 | C380.00 | Obesity | Feb-09 |
| Obesity | 104129 | 2 | 0 | 0 | 0 | C380600 | Adult-onset obesity | May-12 |
| Obesity | 13278 | 468105 | 852 | 76 | 0 | 22K5.00 | Body mass index 30+ - obesity | Feb-09 |
| Obesity | 59780 | 5635 | 110 | 0 | 0 | 222A.00 | O/E - obese | Feb-09 |
| Obesity | 38294 | 57 | 6 | 0 | 0 | C38y000 | Pickwickian syndrome | Feb-09 |
| Obesity | 103574 | 1211 | 6 | 0 | 0 | C38y011 | Obesity hypoventilation syndrome | Jan-12 |
| Obesity | 17444 | 84 | 88 | 0 | 0 | 66CE.00 | Reason for obesity therapy - occupational | Feb-09 |
| Obesity | 66406 | 664 | 0 | 0 | 0 | C38..00 | Obesity and other hyperalimentation | Feb-09 |
| Severe Mental Illness | 101720 | 2295 | 3 | 0 | 0 | Eu22300 | [X]Paranoid state in remission | Feb-11 |
| Severe Mental Illness | 101987 | 2112 | 1 | 0 | 0 | Eu26.00 | [X]Nonorganic psychosis in remission | Apr-11 |
| Severe Mental Illness | 102311 | 6 | 0 | 0 | 0 | E105000 | Unspecified latent schizophrenia | Jun-11 |
| Severe Mental Illness | 102427 | 14 | 0 | 0 | 0 | E102500 | Catatonic schizophrenia in remission | Jun-11 |
| Severe Mental Illness | 102446 | 3 | 0 | 0 | 0 | E105z00 | Latent schizophrenia NOS | Jun-11 |
| Severe Mental Illness | 103915 | 812 | 3 | 0 | 0 | Eu31900 | [X]Bipolar affective disorder type II | Mar-12 |
| Severe Mental Illness | 104051 | 62 | 0 | 0 | 0 | Eu31911 | [X]Bipolar II disorder | May-12 |
| Severe Mental Illness | 104065 | 93 | 0 | 0 | 0 | Eu31800 | [X]Bipolar affective disorder type I | May-12 |
| Severe Mental Illness | 104760 | 5 | 0 | 0 | 0 | E103100 | Subchronic paranoid schizophrenia | Aug-12 |
| Severe Mental Illness | 104763 | 3 | 0 | 0 | 0 | Eu25211 | [X]Cyclic schizophrenia | Aug-12 |
| Severe Mental Illness | 10575 | 354 | 1 | 0 | 0 | E107z00 | Schizo-affective schizophrenia NOS | Feb-09 |
| Severe Mental Illness | 107222 | 2 | 0 | 0 | 0 | E102400 | Acute exacerbation of chronic catatonic schizophrenia | Nov-13 |
| Severe Mental Illness | 109485 | 2 | 0 | 0 | 0 | E110500 | Single manic episode in partial or unspecified remission | Apr-15 |
| Severe Mental Illness | 109634 | 1 | 0 | 0 | 0 | E121.11 | Sander's disease | May-15 |
| Severe Mental Illness | 11055 | 585 | 4 | 0 | 0 | Eu25100 | [X]Schizoaffective disorder, depressive type | Feb-09 |
| Severe Mental Illness | 11172 | 2442 | 225 | 0 | 0 | Eu22012 | [X]Paranoid state | Feb-09 |
| Severe Mental Illness | 11244 | 1964 | 19 | 0 | 0 | Eu2z.00 | [X]Unspecified nonorganic psychosis | Feb-09 |
| Severe Mental Illness | 113612 | 5 | 0 | 0 | 0 | 8G13100 | CBTp - cognitive behavioural therapy for psychosis | Apr-19 |
| Severe Mental Illness | 11548 | 2356 | 103 | 0 | 0 | 146D.00 | H/O: manic depressive disorder | Feb-09 |
| Severe Mental Illness | 11596 | 1062 | 2 | 0 | 0 | E11y000 | Unspecified manic-depressive psychoses | Feb-09 |
| Severe Mental Illness | 12173 | 3790 | 64 | 0 | 0 | Eu30.00 | [X]Manic episode | Feb-09 |
| Severe Mental Illness | 12771 | 8055 | 479 | 0 | 0 | E12z.00 | Paranoid psychosis NOS | Feb-09 |
| Severe Mental Illness | 12777 | 6197 | 415 | 0 | 0 | 146H.00 | H/O: psychosis | Feb-09 |
| Severe Mental Illness | 12831 | 1094 | 67 | 0 | 0 | E115.11 | Manic-depressive - now depressed | Feb-09 |
| Severe Mental Illness | 14728 | 4022 | 579 | 0 | 0 | E110100 | Single manic episode, mild | Feb-09 |
| Severe Mental Illness | 14743 | 3671 | 633 | 0 | 0 | E120.00 | Simple paranoid state | Feb-09 |
| Severe Mental Illness | 14784 | 7358 | 566 | 0 | 0 | E117.00 | Unspecified bipolar affective disorder | Feb-09 |
| Severe Mental Illness | 1494 | 37992 | 1182 | 0 | 0 | E103.00 | Paranoid schizophrenia | Feb-09 |
| Severe Mental Illness | 14965 | 13219 | 1712 | 0 | 0 | E13z.00 | Nonorganic psychosis NOS | Feb-09 |
| Severe Mental Illness | 14971 | 2685 | 203 | 0 | 0 | E122.00 | Paraphrenia | Feb-09 |
| Severe Mental Illness | 1531 | 9125 | 776 | 14 | 0 | Eu31.11 | [X]Manic-depressive illness | Feb-09 |
| Severe Mental Illness | 15733 | 4061 | 110 | 0 | 0 | E100000 | Unspecified schizophrenia | Feb-09 |
| Severe Mental Illness | 15923 | 110 | 4 | 0 | 0 | E115000 | Bipolar affective disorder, currently depressed, unspecified | Feb-09 |
| Severe Mental Illness | 15958 | 2503 | 82 | 0 | 0 | E1...00 | Non-organic psychoses | Feb-09 |
| Severe Mental Illness | 16347 | 38 | 1 | 0 | 0 | E114300 | Bipolar affect disord, currently manic, severe, no psychosis | Feb-09 |
| Severe Mental Illness | 16537 | 65 | 2 | 0 | 0 | E1y..00 | Other specified non-organic psychoses | Feb-09 |
| Severe Mental Illness | 16562 | 558 | 9 | 0 | 0 | Eu31300 | [X]Bipolar affect disorder cur epi mild or moderate depressn | Feb-09 |
| Severe Mental Illness | 16764 | 2887 | 36 | 0 | 0 | Eu20000 | [X]Paranoid schizophrenia | Feb-09 |
| Severe Mental Illness | 16808 | 1453 | 19 | 0 | 0 | Eu31000 | [X]Bipolar affective disorder, current episode hypomanic | Feb-09 |
| Severe Mental Illness | 16905 | 71 | 2 | 0 | 0 | Eu25011 | [X]Schizoaffective psychosis, manic type | Feb-09 |
| Severe Mental Illness | 17281 | 2265 | 57 | 0 | 0 | Eu2..00 | [X]Schizophrenia, schizotypal and delusional disorders | Feb-09 |
| Severe Mental Illness | 17385 | 688 | 53 | 0 | 0 | E114.11 | Manic-depressive - now manic | Feb-09 |
| Severe Mental Illness | 18053 | 176 | 21 | 0 | 0 | Eu20y13 | [X]Schizophrenifrm psychos NOS | Feb-09 |
| Severe Mental Illness | 18909 | 1521 | 56 | 0 | 0 | E110.11 | Hypomanic psychoses | Feb-09 |
| Severe Mental Illness | 19345 | 2548 | 86 | 0 | 0 | 212T.00 | Psychosis, schizophrenia + bipolar affective disord resolved | Feb-09 |
| Severe Mental Illness | 19967 | 317 | 18 | 0 | 0 | E111000 | Recurrent manic episodes, unspecified | Feb-09 |
| Severe Mental Illness | 20110 | 1190 | 217 | 0 | 0 | E110000 | Single manic episode, unspecified | Feb-09 |
| Severe Mental Illness | 20572 | 139 | 7 | 0 | 0 | Eu20211 | [X]Catatonic stupor | Feb-09 |
| Severe Mental Illness | 20785 | 70 | 0 | 0 | 0 | Eu20400 | [X]Post-schizophrenic depression | Feb-09 |
| Severe Mental Illness | 2113 | 14446 | 799 | 1 | 0 | Eu22011 | [X]Paranoid psychosis | Feb-09 |
| Severe Mental Illness | 2117 | 8333 | 612 | 0 | 0 | E107.00 | Schizo-affective schizophrenia | Feb-09 |
| Severe Mental Illness | 21455 | 50 | 2 | 0 | 0 | Eu23012 | [X]Cycloid psychosis | Feb-09 |
| Severe Mental Illness | 22080 | 140 | 7 | 0 | 0 | ZV11112 | [V]Personal history of manic-depressive psychosis | Feb-09 |
| Severe Mental Illness | 22104 | 1521 | 28 | 0 | 0 | ZV11000 | [V]Personal history of schizophrenia | Feb-09 |
| Severe Mental Illness | 22188 | 619 | 20 | 0 | 0 | E1z..00 | Non-organic psychosis NOS | Feb-09 |
| Severe Mental Illness | 22644 | 441 | 80 | 0 | 0 | 286..11 | Poor insight into psychotic condition | Feb-09 |
| Severe Mental Illness | 22713 | 688 | 61 | 0 | 0 | 1S42.00 | Manic mood | Feb-09 |
| Severe Mental Illness | 23616 | 293 | 0 | 0 | 0 | E100100 | Subchronic schizophrenia | Feb-09 |
| Severe Mental Illness | 23713 | 157 | 9 | 0 | 0 | Eu31400 | [X]Bipol aff disord, curr epis sev depress, no psychot symp | Feb-09 |
| Severe Mental Illness | 23963 | 312 | 5 | 0 | 0 | ZV11111 | [V]Personal history of manic-depressive psychosis | Feb-09 |
| Severe Mental Illness | 24107 | 711 | 23 | 0 | 0 | Eu20511 | [X]Chronic undifferentiated schizophrenia | Feb-09 |
| Severe Mental Illness | 24171 | 836 | 9 | 0 | 0 | E113400 | Recurrent major depressive episodes, severe, with psychosis | Feb-09 |
| Severe Mental Illness | 24230 | 465 | 0 | 0 | 0 | E117600 | Unspecified bipolar affective disorder, in full remission | Feb-09 |
| Severe Mental Illness | 24345 | 93 | 1 | 0 | 0 | E134.00 | Psychogenic paranoid psychosis | Feb-09 |
| Severe Mental Illness | 24640 | 29 | 1 | 0 | 0 | E110200 | Single manic episode, moderate | Feb-09 |
| Severe Mental Illness | 24689 | 44 | 0 | 0 | 0 | E116100 | Mixed bipolar affective disorder, mild | Feb-09 |
| Severe Mental Illness | 25546 | 388 | 14 | 0 | 0 | E102.00 | Catatonic schizophrenia | Feb-09 |
| Severe Mental Illness | 25697 | 3374 | 33 | 0 | 0 | E113300 | Recurrent major depressive episodes, severe, no psychosis | Feb-09 |
| Severe Mental Illness | 26143 | 14 | 4 | 0 | 0 | Eu23112 | [X]Cycloid psychosis with symptoms of schizophrenia | Feb-09 |
| Severe Mental Illness | 26161 | 857 | 28 | 0 | 0 | E11..13 | Manic psychoses | Feb-09 |
| Severe Mental Illness | 26227 | 346 | 6 | 0 | 0 | E111.00 | Recurrent manic episodes | Feb-09 |
| Severe Mental Illness | 26299 | 92 | 0 | 0 | 0 | Eu31100 | [X]Bipolar affect disorder cur epi manic wout psychotic symp | Feb-09 |
| Severe Mental Illness | 2741 | 18669 | 1241 | 2 | 0 | Eu30000 | [X]Hypomania | Feb-09 |
| Severe Mental Illness | 27584 | 2506 | 2 | 0 | 0 | Eu31700 | [X]Bipolar affective disorder, currently in remission | Feb-09 |
| Severe Mental Illness | 27739 | 36 | 0 | 0 | 0 | E111200 | Recurrent manic episodes, moderate | Feb-09 |
| Severe Mental Illness | 27770 | 209 | 8 | 0 | 0 | Eu23312 | [X]Psychogenic paranoid psychosis | Feb-09 |
| Severe Mental Illness | 27890 | 93 | 1 | 0 | 0 | E115200 | Bipolar affective disorder, currently depressed, moderate | Feb-09 |
| Severe Mental Illness | 27986 | 502 | 11 | 0 | 0 | E117z00 | Unspecified bipolar affective disorder, NOS | Feb-09 |
| Severe Mental Illness | 28168 | 110 | 2 | 0 | 0 | Eu44.14 | [X]Hysterical psychosis | Feb-09 |
| Severe Mental Illness | 28277 | 218 | 1 | 0 | 0 | Eu31200 | [X]Bipolar affect disorder cur epi manic with psychotic symp | Feb-09 |
| Severe Mental Illness | 28562 | 1014 | 9 | 0 | 0 | Eu22.00 | [X]Persistent delusional disorders | Feb-09 |
| Severe Mental Illness | 28677 | 159 | 1 | 0 | 0 | Eu33312 | [X]Manic-depress psychosis,depressed type+psychotic symptoms | Feb-09 |
| Severe Mental Illness | 29451 | 147 | 7 | 0 | 0 | Eu33213 | [X]Manic-depress psychosis,depressd,no psychotic symptoms | Feb-09 |
| Severe Mental Illness | 29651 | 535 | 20 | 0 | 0 | Eu23z12 | [X]Reactive psychosis | Feb-09 |
| Severe Mental Illness | 29937 | 288 | 4 | 0 | 0 | E131.00 | Acute hysterical psychosis | Feb-09 |
| Severe Mental Illness | 30282 | 382 | 48 | 0 | 0 | ZRby100 | Profile of mood states, bipolar | Feb-09 |
| Severe Mental Illness | 30619 | 546 | 5 | 0 | 0 | E101.00 | Hebephrenic schizophrenia | Feb-09 |
| Severe Mental Illness | 30985 | 546 | 6 | 0 | 0 | Eu2y.00 | [X]Other nonorganic psychotic disorders | Feb-09 |
| Severe Mental Illness | 31316 | 2131 | 52 | 0 | 0 | E116.00 | Mixed bipolar affective disorder | Feb-09 |
| Severe Mental Illness | 31362 | 415 | 5 | 0 | 0 | E103200 | Chronic paranoid schizophrenia | Feb-09 |
| Severe Mental Illness | 31455 | 202 | 4 | 0 | 0 | E12yz00 | Other paranoid states NOS | Feb-09 |
| Severe Mental Illness | 31493 | 2 | 0 | 0 | 0 | Eu20214 | [X]Schizophrenic flexibilatis cerea | Feb-09 |
| Severe Mental Illness | 31535 | 735 | 2 | 0 | 0 | E116000 | Mixed bipolar affective disorder, unspecified | Feb-09 |
| Severe Mental Illness | 31589 | 175 | 3 | 0 | 0 | E12y.00 | Other paranoid states | Feb-09 |
| Severe Mental Illness | 31633 | 291 | 6 | 0 | 0 | Eu3z.11 | [X]Affective psychosis NOS | Feb-09 |
| Severe Mental Illness | 31707 | 189 | 2 | 0 | 0 | Eu23z11 | [X]Brief reactive psychosis NOS | Feb-09 |
| Severe Mental Illness | 31738 | 162 | 10 | 0 | 0 | Eu2y.11 | [X]Chronic hallucinatory psychosis | Feb-09 |
| Severe Mental Illness | 31757 | 63 | 3 | 0 | 0 | Eu33314 | [X]Recurr severe episodes/psychogenic depressive psychosis | Feb-09 |
| Severe Mental Illness | 32088 | 67 | 0 | 0 | 0 | Eu30y00 | [X]Other manic episodes | Feb-09 |
| Severe Mental Illness | 32159 | 547 | 5 | 0 | 0 | E112400 | Single major depressive episode, severe, with psychosis | Feb-09 |
| Severe Mental Illness | 32222 | 1299 | 22 | 0 | 0 | E100.00 | Simple schizophrenia | Feb-09 |
| Severe Mental Illness | 32295 | 44 | 0 | 0 | 0 | E111400 | Recurrent manic episodes, severe, with psychosis | Feb-09 |
| Severe Mental Illness | 33338 | 108 | 16 | 0 | 0 | E10y000 | Atypical schizophrenia | Feb-09 |
| Severe Mental Illness | 33383 | 167 | 2 | 0 | 0 | E103000 | Unspecified paranoid schizophrenia | Feb-09 |
| Severe Mental Illness | 33410 | 325 | 8 | 0 | 0 | Eu25z11 | [X]Schizoaffective psychosis NOS | Feb-09 |
| Severe Mental Illness | 33425 | 405 | 2 | 0 | 0 | E11zz00 | Other affective psychosis NOS | Feb-09 |
| Severe Mental Illness | 33426 | 83 | 1 | 0 | 0 | E11yz00 | Other and unspecified manic-depressive psychoses NOS | Feb-09 |
| Severe Mental Illness | 33693 | 309 | 1 | 0 | 0 | Eu25200 | [X]Schizoaffective disorder, mixed type | Feb-09 |
| Severe Mental Illness | 33751 | 967 | 5 | 0 | 0 | Eu31z00 | [X]Bipolar affective disorder, unspecified | Feb-09 |
| Severe Mental Illness | 33847 | 221 | 0 | 0 | 0 | Eu25000 | [X]Schizoaffective disorder, manic type | Feb-09 |
| Severe Mental Illness | 34236 | 5555 | 103 | 0 | 0 | Eu20.00 | [X]Schizophrenia | Feb-09 |
| Severe Mental Illness | 34389 | 2876 | 41 | 0 | 0 | Eu22000 | [X]Delusional disorder | Feb-09 |
| Severe Mental Illness | 34966 | 278 | 2 | 0 | 0 | Eu20z00 | [X]Schizophrenia, unspecified | Feb-09 |
| Severe Mental Illness | 35274 | 169 | 8 | 0 | 0 | Eu25111 | [X]Schizoaffective psychosis, depressive type | Feb-09 |
| Severe Mental Illness | 35607 | 38 | 0 | 0 | 0 | E115300 | Bipolar affect disord, now depressed, severe, no psychosis | Feb-09 |
| Severe Mental Illness | 35734 | 88 | 1 | 0 | 0 | E115100 | Bipolar affective disorder, currently depressed, mild | Feb-09 |
| Severe Mental Illness | 35738 | 129 | 1 | 0 | 0 | E114000 | Bipolar affective disorder, currently manic, unspecified | Feb-09 |
| Severe Mental Illness | 35848 | 232 | 6 | 0 | 0 | Eu20600 | [X]Simple schizophrenia | Feb-09 |
| Severe Mental Illness | 35877 | 113 | 3 | 0 | 0 | Eu20213 | [X]Schizophrenic catatonia | Feb-09 |
| Severe Mental Illness | 36126 | 65 | 1 | 0 | 0 | E114100 | Bipolar affective disorder, currently manic, mild | Feb-09 |
| Severe Mental Illness | 36172 | 543 | 0 | 0 | 0 | E103500 | Paranoid schizophrenia in remission | Feb-09 |
| Severe Mental Illness | 36611 | 118 | 2 | 0 | 0 | E110z00 | Manic disorder, single episode NOS | Feb-09 |
| Severe Mental Illness | 3702 | 772 | 48 | 0 | 0 | E114.00 | Bipolar affective disorder, currently manic | Feb-09 |
| Severe Mental Illness | 37070 | 1674 | 13 | 0 | 0 | E110.00 | Manic disorder, single episode | Feb-09 |
| Severe Mental Illness | 37178 | 192 | 1 | 0 | 0 | E111600 | Recurrent manic episodes, in full remission | Feb-09 |
| Severe Mental Illness | 37296 | 124 | 3 | 0 | 0 | E115z00 | Bipolar affective disorder, currently depressed, NOS | Feb-09 |
| Severe Mental Illness | 37580 | 183 | 5 | 0 | 0 | Eu25212 | [X]Mixed schizophrenic and affective psychosis | Feb-09 |
| Severe Mental Illness | 37681 | 1016 | 1 | 0 | 0 | Eu25z00 | [X]Schizoaffective disorder, unspecified | Feb-09 |
| Severe Mental Illness | 37764 | 107 | 7 | 0 | 0 | Eu33316 | [X]Recurrent severe episodes/reactive depressive psychosis | Feb-09 |
| Severe Mental Illness | 38063 | 441 | 1 | 0 | 0 | E106.00 | Residual schizophrenia | Feb-09 |
| Severe Mental Illness | 3890 | 2325 | 316 | 1 | 0 | E121.00 | Chronic paranoid psychosis | Feb-09 |
| Severe Mental Illness | 39062 | 151 | 1 | 0 | 0 | E10y.00 | Other schizophrenia | Feb-09 |
| Severe Mental Illness | 39316 | 333 | 1 | 0 | 0 | Eu21.00 | [X]Schizotypal disorder | Feb-09 |
| Severe Mental Illness | 3984 | 4475 | 248 | 0 | 0 | E100200 | Chronic schizophrenic | Feb-09 |
| Severe Mental Illness | 40386 | 40 | 0 | 0 | 0 | Eu21.15 | [X]Prodromal schizophrenia | Feb-09 |
| Severe Mental Illness | 40981 | 181 | 7 | 0 | 0 | Eu22y11 | [X]Delusional dysmorphophobia | Feb-09 |
| Severe Mental Illness | 41022 | 61 | 0 | 0 | 0 | Eu25112 | [X]Schizophreniform psychosis, depressive type | Feb-09 |
| Severe Mental Illness | 4261 | 13722 | 1151 | 0 | 0 | E12..00 | Paranoid states | Feb-09 |
| Severe Mental Illness | 43093 | 14 | 1 | 0 | 0 | E110300 | Single manic episode, severe without mention of psychosis | Feb-09 |
| Severe Mental Illness | 43405 | 45 | 0 | 0 | 0 | Eu20100 | [X]Hebephrenic schizophrenia | Feb-09 |
| Severe Mental Illness | 43800 | 138 | 4 | 0 | 0 | E107200 | Chronic schizo-affective schizophrenia | Feb-09 |
| Severe Mental Illness | 4390 | 489 | 84 | 0 | 0 | 285..11 | Psychotic condition, insight present | Feb-09 |
| Severe Mental Illness | 44498 | 140 | 2 | 0 | 0 | E100400 | Acute exacerbation of chronic schizophrenia | Feb-09 |
| Severe Mental Illness | 44513 | 312 | 3 | 0 | 0 | Eu30z00 | [X]Manic episode, unspecified | Feb-09 |
| Severe Mental Illness | 44693 | 178 | 4 | 0 | 0 | Eu31600 | [X]Bipolar affective disorder, current episode mixed | Feb-09 |
| Severe Mental Illness | 46415 | 300 | 1 | 0 | 0 | E111z00 | Recurrent manic episode NOS | Feb-09 |
| Severe Mental Illness | 46425 | 25 | 0 | 0 | 0 | E111100 | Recurrent manic episodes, mild | Feb-09 |
| Severe Mental Illness | 46434 | 64 | 1 | 0 | 0 | E114200 | Bipolar affective disorder, currently manic, moderate | Feb-09 |
| Severe Mental Illness | 4677 | 828 | 41 | 0 | 0 | E115.00 | Bipolar affective disorder, currently depressed | Feb-09 |
| Severe Mental Illness | 4732 | 146 | 2 | 1 | 0 | Eu31500 | [X]Bipolar affect dis cur epi severe depres with psyc symp | Feb-09 |
| Severe Mental Illness | 47947 | 91 | 0 | 0 | 0 | Eu22013 | [X]Paraphrenia - late | Feb-09 |
| Severe Mental Illness | 48054 | 14 | 0 | 0 | 0 | E101z00 | Hebephrenic schizophrenia NOS | Feb-09 |
| Severe Mental Illness | 4843 | 15364 | 2052 | 0 | 0 | Eu22015 | [X]Paranoia | Feb-09 |
| Severe Mental Illness | 49223 | 161 | 1 | 0 | 0 | Eu22z00 | [X]Persistent delusional disorder, unspecified | Feb-09 |
| Severe Mental Illness | 49420 | 59 | 1 | 0 | 0 | Eu20y00 | [X]Other schizophrenia | Feb-09 |
| Severe Mental Illness | 49761 | 77 | 0 | 0 | 0 | E10yz00 | Other schizophrenia NOS | Feb-09 |
| Severe Mental Illness | 49763 | 141 | 0 | 0 | 0 | E117000 | Unspecified bipolar affective disorder, unspecified | Feb-09 |
| Severe Mental Illness | 49852 | 11 | 0 | 0 | 0 | Eu21.16 | [X]Pseudoneurotic schizophrenia | Feb-09 |
| Severe Mental Illness | 50060 | 24 | 0 | 0 | 0 | Eu20011 | [X]Paraphrenic schizophrenia | Feb-09 |
| Severe Mental Illness | 50218 | 60 | 3 | 0 | 0 | E110400 | Single manic episode, severe, with psychosis | Feb-09 |
| Severe Mental Illness | 50248 | 22 | 0 | 0 | 0 | Eu22y12 | [X]Involutional paranoid state | Feb-09 |
| Severe Mental Illness | 51032 | 270 | 9 | 0 | 0 | Eu31y12 | [X]Recurrent manic episodes | Feb-09 |
| Severe Mental Illness | 51322 | 23 | 0 | 0 | 0 | E103300 | Acute exacerbation of subchronic paranoid schizophrenia | Feb-09 |
| Severe Mental Illness | 51903 | 58 | 1 | 0 | 0 | Eu25012 | [X]Schizophreniform psychosis, manic type | Feb-09 |
| Severe Mental Illness | 53032 | 147 | 0 | 0 | 0 | E103400 | Acute exacerbation of chronic paranoid schizophrenia | Feb-09 |
| Severe Mental Illness | 53625 | 124 | 0 | 0 | 0 | E100z00 | Simple schizophrenia NOS | Feb-09 |
| Severe Mental Illness | 53840 | 197 | 1 | 0 | 0 | Eu31y00 | [X]Other bipolar affective disorders | Feb-09 |
| Severe Mental Illness | 53985 | 9 | 1 | 0 | 0 | Eu20111 | [X]Disorganised schizophrenia | Feb-09 |
| Severe Mental Illness | 54195 | 63 | 0 | 0 | 0 | E116400 | Mixed bipolar affective disorder, severe, with psychosis | Feb-09 |
| Severe Mental Illness | 54387 | 22 | 0 | 0 | 0 | Eu21.12 | [X]Borderline schizophrenia | Feb-09 |
| Severe Mental Illness | 55064 | 161 | 0 | 0 | 0 | E116600 | Mixed bipolar affective disorder, in full remission | Feb-09 |
| Severe Mental Illness | 55221 | 58 | 2 | 0 | 0 | Eu22111 | [X]Capgras syndrome | Feb-09 |
| Severe Mental Illness | 55236 | 14 | 2 | 0 | 0 | Eu22y13 | [X]Paranoia querulans | Feb-09 |
| Severe Mental Illness | 55829 | 74 | 1 | 0 | 0 | E114400 | Bipolar affect disord, currently manic,severe with psychosis | Feb-09 |
| Severe Mental Illness | 56260 | 134 | 5 | 0 | 0 | 13Y3.00 | Manic-depression association member | Feb-09 |
| Severe Mental Illness | 56438 | 324 | 0 | 0 | 0 | E107500 | Schizo-affective schizophrenia in remission | Feb-09 |
| Severe Mental Illness | 57465 | 138 | 0 | 0 | 0 | E115600 | Bipolar affective disorder, now depressed, in full remission | Feb-09 |
| Severe Mental Illness | 57605 | 110 | 2 | 0 | 0 | E114z00 | Bipolar affective disorder, currently manic, NOS | Feb-09 |
| Severe Mental Illness | 57666 | 20 | 0 | 0 | 0 | E100300 | Acute exacerbation of subchronic schizophrenia | Feb-09 |
| Severe Mental Illness | 58532 | 73 | 0 | 0 | 0 | Eu25y00 | [X]Other schizoaffective disorders | Feb-09 |
| Severe Mental Illness | 58687 | 2094 | 2 | 0 | 0 | E100500 | Schizophrenia in remission | Feb-09 |
| Severe Mental Illness | 58716 | 6 | 0 | 0 | 0 | E102000 | Unspecified catatonic schizophrenia | Feb-09 |
| Severe Mental Illness | 58862 | 198 | 1 | 0 | 0 | E107000 | Unspecified schizo-affective schizophrenia | Feb-09 |
| Severe Mental Illness | 58863 | 3 | 0 | 0 | 0 | E111500 | Recurrent manic episodes, partial or unspecified remission | Feb-09 |
| Severe Mental Illness | 58866 | 10 | 0 | 0 | 0 | E107300 | Acute exacerbation subchronic schizo-affective schizophrenia | Feb-09 |
| Severe Mental Illness | 59011 | 13 | 0 | 0 | 0 | E114500 | Bipolar affect disord,currently manic, part/unspec remission | Feb-09 |
| Severe Mental Illness | 60013 | 77 | 0 | 0 | 0 | Eu20300 | [X]Undifferentiated schizophrenia | Feb-09 |
| Severe Mental Illness | 60178 | 405 | 4 | 0 | 0 | E11y.00 | Other and unspecified manic-depressive psychoses | Feb-09 |
| Severe Mental Illness | 61098 | 12 | 0 | 0 | 0 | E107100 | Subchronic schizo-affective schizophrenia | Feb-09 |
| Severe Mental Illness | 61501 | 35 | 0 | 0 | 0 | Eu20200 | [X]Catatonic schizophrenia | Feb-09 |
| Severe Mental Illness | 62405 | 8 | 0 | 0 | 0 | Eu22100 | [X]Delusional misidentification syndrome | Feb-09 |
| Severe Mental Illness | 62449 | 6 | 0 | 0 | 0 | Eu21.14 | [X]Prepsychotic schizophrenia | Feb-09 |
| Severe Mental Illness | 63150 | 65 | 0 | 0 | 0 | E116200 | Mixed bipolar affective disorder, moderate | Feb-09 |
| Severe Mental Illness | 6325 | 12696 | 581 | 0 | 0 | 1464 | H/O: schizophrenia | Feb-09 |
| Severe Mental Illness | 63284 | 31 | 0 | 0 | 0 | E116300 | Mixed bipolar affective disorder, severe, without psychosis | Feb-09 |
| Severe Mental Illness | 63478 | 33 | 0 | 0 | 0 | E107400 | Acute exacerbation of chronic schizo-affective schizophrenia | Feb-09 |
| Severe Mental Illness | 63583 | 256 | 1 | 0 | 0 | E116z00 | Mixed bipolar affective disorder, NOS | Feb-09 |
| Severe Mental Illness | 63651 | 10 | 0 | 0 | 0 | E116500 | Mixed bipolar affective disorder, partial/unspec remission | Feb-09 |
| Severe Mental Illness | 63698 | 17 | 0 | 0 | 0 | E117100 | Unspecified bipolar affective disorder, mild | Feb-09 |
| Severe Mental Illness | 63701 | 23 | 0 | 0 | 0 | E115400 | Bipolar affect disord, now depressed, severe with psychosis | Feb-09 |
| Severe Mental Illness | 63784 | 94 | 1 | 0 | 0 | E114600 | Bipolar affective disorder, currently manic, full remission | Feb-09 |
| Severe Mental Illness | 63867 | 13 | 0 | 0 | 0 | E102z00 | Catatonic schizophrenia NOS | Feb-09 |
| Severe Mental Illness | 64264 | 39 | 0 | 0 | 0 | Eu20500 | [X]Residual schizophrenia | Feb-09 |
| Severe Mental Illness | 64533 | 4 | 0 | 0 | 0 | Eu20212 | [X]Schizophrenic catalepsy | Feb-09 |
| Severe Mental Illness | 64993 | 2 | 0 | 0 | 0 | Eu21.13 | [X]Latent schizophrenia | Feb-09 |
| Severe Mental Illness | 65127 | 9 | 0 | 0 | 0 | Eu22014 | [X]Sensitiver Beziehungswahn | Feb-09 |
| Severe Mental Illness | 65811 | 35 | 0 | 0 | 0 | E111300 | Recurrent manic episodes, severe without mention psychosis | Feb-09 |
| Severe Mental Illness | 66077 | 48 | 0 | 0 | 0 | Eu22y00 | [X]Other persistent delusional disorders | Feb-09 |
| Severe Mental Illness | 66153 | 28 | 1 | 0 | 0 | Eu31.13 | [X]Manic-depressive reaction | Feb-09 |
| Severe Mental Illness | 66410 | 28 | 0 | 0 | 0 | E105.00 | Latent schizophrenia | Feb-09 |
| Severe Mental Illness | 66506 | 2 | 0 | 0 | 0 | E101000 | Unspecified hebephrenic schizophrenia | Feb-09 |
| Severe Mental Illness | 66766 | 10 | 1 | 0 | 0 | E12y000 | Paranoia querulans | Feb-09 |
| Severe Mental Illness | 6710 | 1972 | 35 | 0 | 0 | Eu31.12 | [X]Manic-depressive psychosis | Feb-09 |
| Severe Mental Illness | 67768 | 19 | 0 | 0 | 0 | E101500 | Hebephrenic schizophrenia in remission | Feb-09 |
| Severe Mental Illness | 68326 | 17 | 0 | 0 | 0 | E117400 | Unspecified bipolar affective disorder,severe with psychosis | Feb-09 |
| Severe Mental Illness | 68647 | 21 | 0 | 0 | 0 | E117200 | Unspecified bipolar affective disorder, moderate | Feb-09 |
| Severe Mental Illness | 6874 | 71198 | 2376 | 0 | 0 | Eu31.00 | [X]Bipolar affective disorder | Feb-09 |
| Severe Mental Illness | 694 | 26935 | 1876 | 3 | 0 | Eu2z.11 | [X]Psychosis NOS | Feb-09 |
| Severe Mental Illness | 70000 | 883 | 0 | 0 | 0 | E110600 | Single manic episode in full remission | Feb-09 |
| Severe Mental Illness | 70399 | 27 | 0 | 0 | 0 | E11y300 | Other mixed manic-depressive psychoses | Feb-09 |
| Severe Mental Illness | 70721 | 8 | 0 | 0 | 0 | E117500 | Unspecified bipolar affect disord, partial/unspec remission | Feb-09 |
| Severe Mental Illness | 70925 | 6 | 0 | 0 | 0 | E11y100 | Atypical manic disorder | Feb-09 |
| Severe Mental Illness | 71250 | 1 | 0 | 0 | 0 | Eu21.17 | [X]Pseudopsychopathic schizophrenia | Feb-09 |
| Severe Mental Illness | 72026 | 11 | 0 | 0 | 0 | E115500 | Bipolar affect disord, now depressed, part/unspec remission | Feb-09 |
| Severe Mental Illness | 73295 | 8 | 0 | 0 | 0 | E100.11 | Schizophrenia simplex | Feb-09 |
| Severe Mental Illness | 73423 | 5 | 0 | 0 | 0 | E117300 | Unspecified bipolar affective disorder, severe, no psychosis | Feb-09 |
| Severe Mental Illness | 73924 | 190 | 1 | 0 | 0 | Eu31y11 | [X]Bipolar II disorder | Feb-09 |
| Severe Mental Illness | 8407 | 21618 | 244 | 0 | 0 | E10z.00 | Schizophrenia NOS | Feb-09 |
| Severe Mental Illness | 8478 | 2317 | 54 | 0 | 0 | E130.00 | Reactive depressive psychosis | Feb-09 |
| Severe Mental Illness | 85102 | 550 | 32 | 0 | 0 | 212V.00 | Bipolar affective disorder resolved | Feb-09 |
| Severe Mental Illness | 854 | 73828 | 4299 | 7 | 0 | E10..00 | Schizophrenic disorders | Feb-09 |
| Severe Mental Illness | 8567 | 9575 | 606 | 0 | 0 | E11..11 | Bipolar psychoses | Feb-09 |
| Severe Mental Illness | 85972 | 428 | 0 | 0 | 0 | 212X.00 | Psychosis resolved | Feb-09 |
| Severe Mental Illness | 88275 | 144 | 0 | 0 | 0 | 212W.00 | Schizophrenia resolved | Feb-09 |
| Severe Mental Illness | 91511 | 7 | 0 | 0 | 0 | Eu21.11 | [X]Latent schizophrenic reaction | Feb-09 |
| Severe Mental Illness | 91547 | 5 | 0 | 0 | 0 | Eu20311 | [X]Atypical schizophrenia | Feb-09 |
| Severe Mental Illness | 9281 | 664 | 4 | 0 | 0 | E103z00 | Paranoid schizophrenia NOS | Feb-09 |
| Severe Mental Illness | 92994 | 3 | 0 | 0 | 0 | E10y.11 | Cenesthopathic schizophrenia | Feb-09 |
| Severe Mental Illness | 94001 | 20 | 0 | 0 | 0 | Eu20y12 | [X]Schizophreniform disord NOS | Feb-09 |
| Severe Mental Illness | 9422 | 14088 | 356 | 0 | 0 | Eu25.00 | [X]Schizoaffective disorders | Feb-09 |
| Severe Mental Illness | 94299 | 4 | 0 | 0 | 0 | E105200 | Chronic latent schizophrenia | Feb-09 |
| Severe Mental Illness | 9521 | 1155 | 77 | 0 | 0 | Eu30.11 | [X]Bipolar disorder, single manic episode | Feb-09 |
| Severe Mental Illness | 96883 | 28 | 0 | 0 | 0 | E105500 | Latent schizophrenia in remission | May-09 |
| Severe Mental Illness | 97919 | 1 | 0 | 0 | 0 | E101400 | Acute exacerbation of chronic hebephrenic schizophrenia | Oct-09 |
| Severe Mental Illness | 98821 | 12 | 1 | 0 | 0 | Eu22200 | [X]Cotard syndrome | Mar-10 |
| Severe Mental Illness | 99000 | 1 | 0 | 0 | 0 | E107.11 | Cyclic schizophrenia | Apr-10 |
| Severe Mental Illness | 99070 | 1 | 0 | 0 | 0 | E10y100 | Coenesthopathic schizophrenia | Apr-10 |
| Severe Mental Illness | 99199 | 2 | 0 | 0 | 0 | E102100 | Subchronic catatonic schizophrenia | Apr-10 |
| Smoking Status | 1822 | 52126 | 35 | 1 | 0 | 1376 | Very heavy smoker - 40+cigs/d | Feb-09 |
| Smoking Status | 34126 | 87703 | 703 | 0 | 0 | 13p0.00 | Negotiated date for cessation of smoking | Feb-09 |
| Smoking Status | 100099 | 123260 | 4393 | 0 | 0 | 8IAj.00 | Smoking cessation advice declined | Jul-10 |
| Smoking Status | 10742 | 101352 | 28880 | 0 | 0 | 8HTK.00 | Referral to stop-smoking clinic | Feb-09 |
| Smoking Status | 12966 | 8340 | 1 | 0 | 0 | 137V.00 | Smoking reduced | Feb-09 |
| Smoking Status | 59866 | 8 | 0 | 0 | 0 | ZRh4.00 | Reasons for smoking scale | Feb-09 |
| Smoking Status | 66409 | 59 | 0 | 0 | 0 | 8I2I.00 | Nicotine replacement therapy contraindicated | Feb-09 |
| Smoking Status | 32572 | 5286 | 0 | 0 | 0 | 8B3Y.00 | Over the counter nicotine replacement therapy | Feb-09 |
| Smoking Status | 97643 | 363 | 0 | 0 | 0 | 38DH.00 | Fagerstrom test for nicotine dependence | Sep-09 |
| Smoking Status | 63666 | 2902 | 0 | 0 | 0 | ZRBm200 | Fagerstrom test for nicotine dependence | Feb-09 |
| Smoking Status | 63299 | 27 | 0 | 0 | 0 | ZRBm211 | FTND - Fagerstrom test for nicotine dependence | Feb-09 |
| Smoking Status | 98245 | 1914 | 12 | 0 | 0 | 8HBM.00 | Stop smoking face to face follow-up | Dec-09 |
| Smoking Status | 12942 | 72108 | 37 | 0 | 0 | 137..11 | Smoker - amount smoked | Feb-09 |
| Smoking Status | 67178 | 792 | 0 | 0 | 0 | 8BP3.00 | Nicotine replacement therapy provided by community pharmacis | Feb-09 |
| Smoking Status | 31114 | 21026 | 23 | 0 | 0 | 137b.00 | Ready to stop smoking | Feb-09 |
| Smoking Status | 12952 | 12868 | 10 | 0 | 0 | 137Q.00 | Smoking started | Feb-09 |
| Smoking Status | 49418 | 8 | 0 | 0 | 0 | ZRh4.11 | RFS - Reasons for smoking scale | Feb-09 |
| Smoking Status | 12947 | 83635 | 4 | 0 | 0 | 137H.00 | Pipe smoker | Feb-09 |
| Smoking Status | 90522 | 10302 | 1 | 0 | 0 | 745Hz00 | Smoking cessation therapy NOS | Feb-09 |
| Smoking Status | 94958 | 14818 | 0 | 0 | 0 | 745H400 | Smoking cessation drug therapy | Feb-09 |
| Smoking Status | 103507 | 14057 | 56 | 0 | 0 | 8CdB.00 | Stop smoking service opportunity signposted | Jan-12 |
| Smoking Status | 46321 | 278 | 0 | 0 | 0 | 137f.00 | Reason for restarting smoking | Feb-09 |
| Smoking Status | 41405 | 1439 | 0 | 0 | 0 | 13p3.00 | Smoking status at 52 weeks | Feb-09 |
| Smoking Status | 91708 | 3397 | 0 | 0 | 0 | 745Hy00 | Other specified smoking cessation therapy | Feb-09 |
| Smoking Status | 12240 | 250198 | 1535 | 1 | 0 | 137G.00 | Trying to give up smoking | Feb-09 |
| Smoking Status | 81440 | 17389 | 0 | 0 | 0 | 745H000 | Nicotine replacement therapy using nicotine patches | Feb-09 |
| Smoking Status | 93 | 10053150 | 1181 | 190 | 0 | 137P.00 | Cigarette smoker | Feb-09 |
| Smoking Status | 12944 | 766234 | 58 | 3 | 0 | 1373 | Light smoker - 1-9 cigs/day | Feb-09 |
| Smoking Status | 102361 | 15627 | 57 | 0 | 0 | 9NS0200 | Referral for smoking cessation service offered | Jun-11 |
| Smoking Status | 12964 | 15553 | 12 | 0 | 0 | 137C.00 | Keeps trying to stop smoking | Feb-09 |
| Smoking Status | 34127 | 38693 | 564 | 0 | 0 | 13p1.00 | Smoking status at 4 weeks | Feb-09 |
| Smoking Status | 104310 | 5351 | 0 | 0 | 0 | 9ko..11 | Current smoker annual review | Jun-12 |
| Smoking Status | 1823 | 435204 | 666 | 4 | 0 | 137P.11 | Smoker | Feb-09 |
| Smoking Status | 38112 | 35431 | 3 | 0 | 0 | 13p5.00 | Smoking cessation programme start date | Feb-09 |
| Smoking Status | 47273 | 25 | 0 | 0 | 0 | ZRaM.00 | Motives for smoking scale | Feb-09 |
| Smoking Status | 74907 | 124540 | 204 | 1317 | 0 | 745H.00 | Smoking cessation therapy | Feb-09 |
| Smoking Status | 28886 | 15343 | 0 | 1740 | 0 | 13p6.00 | Carbon monoxide reading at 4 weeks | Feb-09 |
| Smoking Status | 12951 | 10812 | 2 | 0 | 0 | 137Q.11 | Smoking restarted | Feb-09 |
| Smoking Status | 41979 | 1701 | 0 | 0 | 0 | 137e.00 | Smoking restarted | Feb-09 |
| Smoking Status | 30423 | 28002 | 5 | 0 | 0 | 137c.00 | Thinking about stopping smoking | Feb-09 |
| Smoking Status | 101764 | 5828 | 0 | 0 | 0 | 13p5000 | Practice based smoking cessation programme start date | Feb-11 |
| Smoking Status | 101338 | 5086 | 0 | 0 | 0 | 137m.00 | Failed attempt to stop smoking | Jan-11 |
| Smoking Status | 103208 | 230 | 0 | 0 | 0 | 13p7.00 | Smoking status at 12 weeks | Oct-11 |
| Smoking Status | 11356 | 314173 | 36 | 6 | 0 | 9N2k.00 | Seen by smoking cessation advisor | Feb-09 |
| Smoking Status | 12945 | 141770 | 8 | 1 | 0 | 137M.00 | Rolls own cigarettes | Feb-09 |
| Smoking Status | 62686 | 8564 | 0 | 0 | 0 | 137h.00 | Minutes from waking to first tobacco consumption | Feb-09 |
| Smoking Status | 25106 | 14148 | 1 | 0 | 0 | 8B3f.00 | Nicotine replacement therapy provided free | Feb-09 |
| Smoking Status | 1878 | 1143006 | 146 | 12 | 0 | 1374 | Moderate smoker - 10-19 cigs/d | Feb-09 |
| Smoking Status | 91513 | 3 | 0 | 0 | 0 | ZRao.00 | Occasions for smoking scale | Feb-09 |
| Smoking Status | 10558 | 2779707 | 450 | 33 | 0 | 137R.00 | Current smoker | Feb-09 |
| Smoking Status | 34374 | 7089 | 209 | 0 | 0 | 13p2.00 | Smoking status between 4 and 52 weeks | Feb-09 |
| Smoking Status | 41042 | 1574 | 19 | 0 | 0 | 8CAg.00 | Smoking cessation advice provided by community pharmacist | Feb-09 |
| Smoking Status | 12943 | 86666 | 13 | 0 | 0 | 137J.00 | Cigar smoker | Feb-09 |
| Smoking Status | 30762 | 39536 | 0 | 5 | 0 | 137d.00 | Not interested in stopping smoking | Feb-09 |
| Smoking Status | 9045 | 11510 | 69 | 1 | 0 | ZG23300 | Advice on smoking | Feb-09 |
| Smoking Status | 12941 | 204567 | 6 | 0 | 0 | 1372.11 | Occasional smoker | Feb-09 |
| Smoking Status | 98137 | 29160 | 7 | 0 | 0 | 67H6.00 | Brief intervention for smoking cessation | Dec-09 |
| Smoking Status | 18573 | 221359 | 22135 | 20 | 0 | 8H7i.00 | Referral to smoking cessation advisor | Feb-09 |
| Smoking Status | 9833 | 91420 | 23 | 0 | 0 | 8B2B.00 | Nicotine replacement therapy | Feb-09 |
| Smoking Status | 7622 | 12293900 | 13432 | 14 | 0 | 8CAL.00 | Smoking cessation advice | Feb-09 |
| Smoking Status | 89464 | 1622 | 0 | 0 | 0 | 745H300 | Nicotine replacement therapy using nicotine lozenges | Feb-09 |
| Smoking Status | 98347 | 239 | 0 | 0 | 0 | 9ko..00 | Current smoker annual review - enhanced services admin | Jan-10 |
| Smoking Status | 102951 | 307 | 0 | 0 | 0 | 13p8.00 | Lost to smoking cessation follow-up | Sep-11 |
| Smoking Status | 3568 | 578428 | 202 | 8 | 0 | 1375 | Heavy smoker - 20-39 cigs/day | Feb-09 |
| Smoking Status | 10211 | 157560 | 704 | 0 | 0 | 13p..00 | Smoking cessation milestones | Feb-09 |
| Smoking Status | 85975 | 1939 | 2 | 0 | 0 | 745H100 | Nicotine replacement therapy using nicotine gum | Feb-09 |
| Smoking Status | 85274 | 0 | 0 | 0 | 0 | K6091 |  | Feb-09 |
| Smoking Status | 70746 | 48 | 0 | 0 | 0 | E251100 | Tobacco dependence, continuous | Feb-09 |
| Smoking Status | 105710 | 206 | 0 | 0 | 0 | 8HBP.00 | Smoking cessation 12 week follow-up | Jan-13 |
| Smoking Status | 12958 | 151025 | 18 | 0 | 0 | 1372 | Trivial smoker - < 1 cig/day | Feb-09 |
| Smoking Status | 98154 | 46430 | 4351 | 4 | 0 | 8HkQ.00 | Referral to NHS stop smoking service | Dec-09 |
| Smoking Status | 10898 | 12795 | 0 | 23 | 0 | 13p4.00 | Smoking free weeks | Feb-09 |
| Smoking Status | 72706 | 381 | 0 | 0 | 0 | E251300 | Tobacco dependence in remission | Feb-09 |
| Smoking Status | 35055 | 125 | 0 | 0 | 0 | ZV6D800 | [V]Tobacco abuse counselling | Feb-09 |
| Smoking Status | 12954 | 198 | 1 | 0 | 0 | ZV4K000 | [V]Tobacco use | Feb-09 |
| Smoking Status | 32687 | 8870 | 0 | 0 | 0 | E251.00 | Tobacco dependence | Feb-09 |
| Diabetes Mellitus | 10418 | 599 | 10 | 0 | 0 | C10ED00 | Type 1 diabetes mellitus with nephropathy | Feb-09 |
| Diabetes Mellitus | 10692 | 9945 | 129 | 0 | 0 | C10EM00 | Type 1 diabetes mellitus with ketoacidosis | Feb-09 |
| Diabetes Mellitus | 12640 | 2571 | 73 | 0 | 0 | C10FC00 | Type 2 diabetes mellitus with nephropathy | Feb-09 |
| Diabetes Mellitus | 12736 | 79 | 5 | 0 | 0 | C10F500 | Type 2 diabetes mellitus with gangrene | Feb-09 |
| Diabetes Mellitus | 1407 | 20637 | 146 | 0 | 0 | C10FJ00 | Insulin treated Type 2 diabetes mellitus | Feb-09 |
| Diabetes Mellitus | 1549 | 162313 | 3200 | 2 | 0 | C10E.00 | Type 1 diabetes mellitus | Feb-09 |
| Diabetes Mellitus | 17545 | 7 | 1 | 0 | 0 | C108F11 | Type I diabetes mellitus with diabetic cataract | Feb-09 |
| Diabetes Mellitus | 17858 | 7535 | 194 | 0 | 0 | C108.12 | Type 1 diabetes mellitus | Feb-09 |
| Diabetes Mellitus | 17859 | 53448 | 1200 | 2 | 0 | C109.12 | Type 2 diabetes mellitus | Feb-09 |
| Diabetes Mellitus | 18143 | 1 | 0 | 0 | 0 | C109G11 | Type II diabetes mellitus with arthropathy | Feb-09 |
| Diabetes Mellitus | 18209 | 24 | 2 | 0 | 0 | C109012 | Type 2 diabetes mellitus with renal complications | Feb-09 |
| Diabetes Mellitus | 18219 | 6064 | 133 | 0 | 0 | C109.13 | Type II diabetes mellitus | Feb-09 |
| Diabetes Mellitus | 18230 | 3 | 1 | 0 | 0 | C108J12 | Type 1 diabetes mellitus with neuropathic arthropathy | Feb-09 |
| Diabetes Mellitus | 18264 | 97 | 4 | 0 | 0 | C109J12 | Insulin treated Type II diabetes mellitus | Feb-09 |
| Diabetes Mellitus | 18278 | 6071 | 52 | 0 | 0 | C109J00 | Insulin treated Type 2 diabetes mellitus | Feb-09 |
| Diabetes Mellitus | 18387 | 807 | 18 | 0 | 0 | C10E700 | Type 1 diabetes mellitus with retinopathy | Feb-09 |
| Diabetes Mellitus | 18390 | 12666 | 67 | 0 | 0 | C10FM00 | Type 2 diabetes mellitus with persistent microalbuminuria | Feb-09 |
| Diabetes Mellitus | 18425 | 342 | 6 | 0 | 0 | C10FB00 | Type 2 diabetes mellitus with polyneuropathy | Feb-09 |
| Diabetes Mellitus | 18496 | 2735 | 48 | 2 | 0 | C10F600 | Type 2 diabetes mellitus with retinopathy | Feb-09 |
| Diabetes Mellitus | 18642 | 14 | 0 | 0 | 0 | C10EH00 | Type 1 diabetes mellitus with arthropathy | Feb-09 |
| Diabetes Mellitus | 18683 | 72 | 1 | 0 | 0 | C10E500 | Type 1 diabetes mellitus with ulcer | Feb-09 |
| Diabetes Mellitus | 18777 | 294 | 20 | 0 | 0 | C10F000 | Type 2 diabetes mellitus with renal complications | Feb-09 |
| Diabetes Mellitus | 21983 | 5 | 0 | 0 | 0 | C108012 | Type 1 diabetes mellitus with renal complications | Feb-09 |
| Diabetes Mellitus | 22871 | 99 | 4 | 0 | 0 | C10EP00 | Type 1 diabetes mellitus with exudative maculopathy | Feb-09 |
| Diabetes Mellitus | 22884 | 11876 | 12 | 0 | 0 | C10F.11 | Type II diabetes mellitus | Feb-09 |
| Diabetes Mellitus | 24423 | 1090 | 16 | 0 | 0 | C108.13 | Type I diabetes mellitus | Feb-09 |
| Diabetes Mellitus | 24458 | 96 | 5 | 0 | 0 | C109711 | Type II diabetes mellitus - poor control | Feb-09 |
| Diabetes Mellitus | 24836 | 14 | 1 | 0 | 0 | C109C12 | Type 2 diabetes mellitus with nephropathy | Feb-09 |
| Diabetes Mellitus | 25591 | 303 | 5 | 0 | 0 | C10FQ00 | Type 2 diabetes mellitus with exudative maculopathy | Feb-09 |
| Diabetes Mellitus | 25627 | 2031 | 95 | 0 | 0 | C10F700 | Type 2 diabetes mellitus - poor control | Feb-09 |
| Diabetes Mellitus | 26054 | 6476 | 64 | 3 | 0 | C10FL00 | Type 2 diabetes mellitus with persistent proteinuria | Feb-09 |
| Diabetes Mellitus | 30294 | 674 | 7 | 0 | 0 | C10EL00 | Type 1 diabetes mellitus with persistent microalbuminuria | Feb-09 |
| Diabetes Mellitus | 30323 | 781 | 5 | 0 | 0 | C10EK00 | Type 1 diabetes mellitus with persistent proteinuria | Feb-09 |
| Diabetes Mellitus | 32627 | 1468 | 56 | 0 | 0 | C10FN00 | Type 2 diabetes mellitus with ketoacidosis | Feb-09 |
| Diabetes Mellitus | 34268 | 212 | 2 | 0 | 0 | C10F200 | Type 2 diabetes mellitus with neurological complications | Feb-09 |
| Diabetes Mellitus | 34450 | 940 | 15 | 0 | 0 | C10FK00 | Hyperosmolar non-ketotic state in type 2 diabetes mellitus | Feb-09 |
| Diabetes Mellitus | 35288 | 587 | 39 | 0 | 0 | C10E800 | Type 1 diabetes mellitus - poor control | Feb-09 |
| Diabetes Mellitus | 35385 | 291 | 7 | 0 | 0 | C10FH00 | Type 2 diabetes mellitus with neuropathic arthropathy | Feb-09 |
| Diabetes Mellitus | 36633 | 112 | 0 | 0 | 0 | C109K00 | Hyperosmolar non-ketotic state in type 2 diabetes mellitus | Feb-09 |
| Diabetes Mellitus | 37806 | 233 | 3 | 0 | 0 | C10FF00 | Type 2 diabetes mellitus with peripheral angiopathy | Feb-09 |
| Diabetes Mellitus | 38161 | 25 | 1 | 0 | 0 | C108711 | Type I diabetes mellitus with retinopathy | Feb-09 |
| Diabetes Mellitus | 39070 | 137 | 3 | 0 | 0 | C10EE00 | Type 1 diabetes mellitus with hypoglycaemic coma | Feb-09 |
| Diabetes Mellitus | 40682 | 135 | 1 | 0 | 0 | C10E900 | Type 1 diabetes mellitus maturity onset | Feb-09 |
| Diabetes Mellitus | 40837 | 371 | 32 | 0 | 0 | C10EN00 | Type 1 diabetes mellitus with ketoacidotic coma | Feb-09 |
| Diabetes Mellitus | 41049 | 15 | 0 | 0 | 0 | C108712 | Type 1 diabetes mellitus with retinopathy | Feb-09 |
| Diabetes Mellitus | 42729 | 16 | 0 | 0 | 0 | C108E11 | Type I diabetes mellitus with hypoglycaemic coma | Feb-09 |
| Diabetes Mellitus | 42762 | 48 | 8 | 0 | 0 | C109612 | Type 2 diabetes mellitus with retinopathy | Feb-09 |
| Diabetes Mellitus | 42831 | 32 | 1 | 0 | 0 | C10E200 | Type 1 diabetes mellitus with neurological complications | Feb-09 |
| Diabetes Mellitus | 43227 | 10 | 0 | 0 | 0 | C10F311 | Type II diabetes mellitus with multiple complications | Feb-09 |
| Diabetes Mellitus | 43921 | 70 | 2 | 0 | 0 | C10E400 | Unstable type 1 diabetes mellitus | Feb-09 |
| Diabetes Mellitus | 44779 | 13 | 1 | 0 | 0 | C109E12 | Type 2 diabetes mellitus with diabetic cataract | Feb-09 |
| Diabetes Mellitus | 44982 | 253 | 6 | 0 | 0 | C10FE00 | Type 2 diabetes mellitus with diabetic cataract | Feb-09 |
| Diabetes Mellitus | 45913 | 95 | 3 | 0 | 0 | C109712 | Type 2 diabetes mellitus - poor control | Feb-09 |
| Diabetes Mellitus | 45914 | 22 | 0 | 0 | 0 | C108812 | Type 1 diabetes mellitus - poor control | Feb-09 |
| Diabetes Mellitus | 45919 | 17 | 0 | 0 | 0 | C109212 | Type 2 diabetes mellitus with neurological complications | Feb-09 |
| Diabetes Mellitus | 46150 | 12 | 0 | 0 | 0 | C109512 | Type 2 diabetes mellitus with gangrene | Feb-09 |
| Diabetes Mellitus | 46301 | 41 | 0 | 0 | 0 | C10EC00 | Type 1 diabetes mellitus with polyneuropathy | Feb-09 |
| Diabetes Mellitus | 46850 | 21 | 0 | 0 | 0 | C108811 | Type I diabetes mellitus - poor control | Feb-09 |
| Diabetes Mellitus | 46917 | 167 | 3 | 0 | 0 | C10FD00 | Type 2 diabetes mellitus with hypoglycaemic coma | Feb-09 |
| Diabetes Mellitus | 47315 | 122 | 0 | 0 | 0 | C10F711 | Type II diabetes mellitus - poor control | Feb-09 |
| Diabetes Mellitus | 47321 | 124 | 1 | 0 | 0 | C10F100 | Type 2 diabetes mellitus with ophthalmic complications | Feb-09 |
| Diabetes Mellitus | 47409 | 5 | 0 | 0 | 0 | C109B11 | Type II diabetes mellitus with polyneuropathy | Feb-09 |
| Diabetes Mellitus | 47582 | 72 | 3 | 0 | 0 | C10E000 | Type 1 diabetes mellitus with renal complications | Feb-09 |
| Diabetes Mellitus | 47649 | 37 | 2 | 0 | 0 | C10E100 | Type 1 diabetes mellitus with ophthalmic complications | Feb-09 |
| Diabetes Mellitus | 47650 | 52 | 0 | 0 | 0 | C10E300 | Type 1 diabetes mellitus with multiple complications | Feb-09 |
| Diabetes Mellitus | 47816 | 9 | 3 | 0 | 0 | C109H11 | Type II diabetes mellitus with neuropathic arthropathy | Feb-09 |
| Diabetes Mellitus | 47954 | 1045 | 6 | 0 | 0 | C10F900 | Type 2 diabetes mellitus without complication | Feb-09 |
| Diabetes Mellitus | 48192 | 15 | 4 | 0 | 0 | C109E11 | Type II diabetes mellitus with diabetic cataract | Feb-09 |
| Diabetes Mellitus | 49074 | 188 | 5 | 0 | 0 | C10F400 | Type 2 diabetes mellitus with ulcer | Feb-09 |
| Diabetes Mellitus | 49146 | 2 | 0 | 0 | 0 | C108211 | Type I diabetes mellitus with neurological complications | Feb-09 |
| Diabetes Mellitus | 49554 | 24 | 0 | 0 | 0 | C10EF00 | Type 1 diabetes mellitus with diabetic cataract | Feb-09 |
| Diabetes Mellitus | 49655 | 71 | 0 | 0 | 0 | C10F611 | Type II diabetes mellitus with retinopathy | Feb-09 |
| Diabetes Mellitus | 49869 | 1 | 0 | 0 | 0 | C109G12 | Type 2 diabetes mellitus with arthropathy | Feb-09 |
| Diabetes Mellitus | 49949 | 14 | 1 | 0 | 0 | C10E411 | Unstable type I diabetes mellitus | Feb-09 |
| Diabetes Mellitus | 50225 | 11 | 4 | 0 | 0 | C109011 | Type II diabetes mellitus with renal complications | Feb-09 |
| Diabetes Mellitus | 50527 | 49 | 0 | 0 | 0 | C10FB11 | Type II diabetes mellitus with polyneuropathy | Feb-09 |
| Diabetes Mellitus | 50813 | 1 | 0 | 0 | 0 | C109A11 | Type II diabetes mellitus with mononeuropathy | Feb-09 |
| Diabetes Mellitus | 51756 | 44 | 1 | 0 | 0 | C10FP00 | Type 2 diabetes mellitus with ketoacidotic coma | Feb-09 |
| Diabetes Mellitus | 51957 | 13 | 1 | 0 | 0 | C108511 | Type I diabetes mellitus with ulcer | Feb-09 |
| Diabetes Mellitus | 53392 | 331 | 0 | 0 | 0 | C10F911 | Type II diabetes mellitus without complication | Feb-09 |
| Diabetes Mellitus | 54008 | 88 | 1 | 0 | 0 | C10EJ00 | Type 1 diabetes mellitus with neuropathic arthropathy | Feb-09 |
| Diabetes Mellitus | 54899 | 4 | 0 | 0 | 0 | C109F11 | Type II diabetes mellitus with peripheral angiopathy | Feb-09 |
| Diabetes Mellitus | 55075 | 24 | 1 | 0 | 0 | C109411 | Type II diabetes mellitus with ulcer | Feb-09 |
| Diabetes Mellitus | 55239 | 798 | 10 | 0 | 0 | C10EQ00 | Type 1 diabetes mellitus with gastroparesis | Feb-09 |
| Diabetes Mellitus | 56268 | 2 | 0 | 0 | 0 | C109D11 | Type II diabetes mellitus with hypoglycaemic coma | Feb-09 |
| Diabetes Mellitus | 57278 | 10 | 0 | 0 | 0 | C10F011 | Type II diabetes mellitus with renal complications | Feb-09 |
| Diabetes Mellitus | 58604 | 37 | 12 | 0 | 0 | C109611 | Type II diabetes mellitus with retinopathy | Feb-09 |
| Diabetes Mellitus | 59253 | 103 | 2 | 0 | 0 | C10FG00 | Type 2 diabetes mellitus with arthropathy | Feb-09 |
| Diabetes Mellitus | 59725 | 6 | 0 | 0 | 0 | C109111 | Type II diabetes mellitus with ophthalmic complications | Feb-09 |
| Diabetes Mellitus | 60107 | 4 | 0 | 0 | 0 | C108411 | Unstable type I diabetes mellitus | Feb-09 |
| Diabetes Mellitus | 60208 | 3 | 0 | 0 | 0 | C108J11 | Type I diabetes mellitus with neuropathic arthropathy | Feb-09 |
| Diabetes Mellitus | 60699 | 5 | 0 | 0 | 0 | C109F12 | Type 2 diabetes mellitus with peripheral angiopathy | Feb-09 |
| Diabetes Mellitus | 60796 | 67 | 0 | 0 | 0 | C10FL11 | Type II diabetes mellitus with persistent proteinuria | Feb-09 |
| Diabetes Mellitus | 61071 | 9 | 1 | 0 | 0 | C109D12 | Type 2 diabetes mellitus with hypoglycaemic coma | Feb-09 |
| Diabetes Mellitus | 61344 | 6 | 0 | 0 | 0 | C108011 | Type I diabetes mellitus with renal complications | Feb-09 |
| Diabetes Mellitus | 61829 | 6 | 0 | 0 | 0 | C108212 | Type 1 diabetes mellitus with neurological complications | Feb-09 |
| Diabetes Mellitus | 62107 | 10 | 1 | 0 | 0 | C109511 | Type II diabetes mellitus with gangrene | Feb-09 |
| Diabetes Mellitus | 62209 | 90 | 1 | 0 | 0 | C10EM11 | Type I diabetes mellitus with ketoacidosis | Feb-09 |
| Diabetes Mellitus | 62352 | 1 | 0 | 0 | 0 | C108H11 | Type I diabetes mellitus with arthropathy | Feb-09 |
| Diabetes Mellitus | 62613 | 4 | 0 | 0 | 0 | C10EA11 | Type I diabetes mellitus without complication | Feb-09 |
| Diabetes Mellitus | 62674 | 86 | 1 | 0 | 0 | C10FA00 | Type 2 diabetes mellitus with mononeuropathy | Feb-09 |
| Diabetes Mellitus | 63017 | 1 | 0 | 0 | 0 | C108911 | Type I diabetes mellitus maturity onset | Feb-09 |
| Diabetes Mellitus | 63690 | 423 | 7 | 0 | 0 | C10FR00 | Type 2 diabetes mellitus with gastroparesis | Feb-09 |
| Diabetes Mellitus | 64571 | 7 | 1 | 0 | 0 | C109C11 | Type II diabetes mellitus with nephropathy | Feb-09 |
| Diabetes Mellitus | 64668 | 164 | 0 | 0 | 0 | C10FJ11 | Insulin treated Type II diabetes mellitus | Feb-09 |
| Diabetes Mellitus | 65267 | 55 | 2 | 0 | 0 | C10F300 | Type 2 diabetes mellitus with multiple complications | Feb-09 |
| Diabetes Mellitus | 65704 | 18 | 3 | 0 | 0 | C109412 | Type 2 diabetes mellitus with ulcer | Feb-09 |
| Diabetes Mellitus | 66145 | 1 | 0 | 0 | 0 | C10EN11 | Type I diabetes mellitus with ketoacidotic coma | Feb-09 |
| Diabetes Mellitus | 66872 | 8 | 0 | 0 | 0 | C108D11 | Type I diabetes mellitus with nephropathy | Feb-09 |
| Diabetes Mellitus | 66965 | 9 | 0 | 0 | 0 | C109H12 | Type 2 diabetes mellitus with neuropathic arthropathy | Feb-09 |
| Diabetes Mellitus | 67905 | 5 | 0 | 0 | 0 | C109211 | Type II diabetes mellitus with neurological complications | Feb-09 |
| Diabetes Mellitus | 68105 | 17 | 0 | 0 | 0 | C10EB00 | Type 1 diabetes mellitus with mononeuropathy | Feb-09 |
| Diabetes Mellitus | 68390 | 15 | 0 | 0 | 0 | C108512 | Type 1 diabetes mellitus with ulcer | Feb-09 |
| Diabetes Mellitus | 69676 | 74 | 1 | 0 | 0 | C10EA00 | Type 1 diabetes mellitus without complication | Feb-09 |
| Diabetes Mellitus | 69993 | 22 | 0 | 0 | 0 | C10E600 | Type 1 diabetes mellitus with gangrene | Feb-09 |
| Diabetes Mellitus | 70316 | 40 | 0 | 0 | 0 | C109112 | Type 2 diabetes mellitus with ophthalmic complications | Feb-09 |
| Diabetes Mellitus | 70766 | 6 | 0 | 0 | 0 | C108E12 | Type 1 diabetes mellitus with hypoglycaemic coma | Feb-09 |
| Diabetes Mellitus | 758 | 1606815 | 17577 | 21 | 0 | C10F.00 | Type 2 diabetes mellitus | Feb-09 |
| Diabetes Mellitus | 85991 | 10 | 0 | 0 | 0 | C10FM11 | Type II diabetes mellitus with persistent microalbuminuria | Feb-09 |
| Diabetes Mellitus | 91646 | 7 | 0 | 0 | 0 | C10F411 | Type II diabetes mellitus with ulcer | Feb-09 |
| Diabetes Mellitus | 91942 | 6 | 0 | 0 | 0 | C10E311 | Type I diabetes mellitus with multiple complications | Feb-09 |
| Diabetes Mellitus | 91943 | 1 | 0 | 0 | 0 | C10EC11 | Type I diabetes mellitus with polyneuropathy | Feb-09 |
| Diabetes Mellitus | 93468 | 8 | 0 | 0 | 0 | C10EG00 | Type 1 diabetes mellitus with peripheral angiopathy | Feb-09 |
| Diabetes Mellitus | 93727 | 181 | 1 | 0 | 0 | C10FE11 | Type II diabetes mellitus with diabetic cataract | Feb-09 |
| Diabetes Mellitus | 93878 | 12 | 0 | 0 | 0 | C10E511 | Type I diabetes mellitus with ulcer | Feb-09 |
| Diabetes Mellitus | 95343 | 30 | 0 | 0 | 0 | C10E711 | Type I diabetes mellitus with retinopathy | Feb-09 |
| Diabetes Mellitus | 95351 | 40 | 0 | 0 | 0 | C10FA11 | Type II diabetes mellitus with mononeuropathy | Feb-09 |
| Diabetes Mellitus | 95992 | 1 | 0 | 0 | 0 | C108A11 | Type I diabetes mellitus without complication | Feb-09 |
| Diabetes Mellitus | 96235 | 35 | 0 | 0 | 0 | C10E911 | Type I diabetes mellitus maturity onset | Apr-09 |
| Diabetes Mellitus | 97446 | 5 | 0 | 0 | 0 | C108912 | Type 1 diabetes mellitus maturity onset | Aug-09 |
| Diabetes Mellitus | 97474 | 7 | 0 | 0 | 0 | C108412 | Unstable type 1 diabetes mellitus | Aug-09 |
| Diabetes Mellitus | 97894 | 3 | 0 | 0 | 0 | C10EP11 | Type I diabetes mellitus with exudative maculopathy | Oct-09 |
| Diabetes Mellitus | 98616 | 3 | 0 | 0 | 0 | C10F211 | Type II diabetes mellitus with neurological complications | Mar-10 |
| Diabetes Mellitus | 98723 | 50 | 0 | 0 | 0 | C10FD11 | Type II diabetes mellitus with hypoglycaemic coma | Mar-10 |
| Depression | 33469 | 2195 | 41 | 0 | 0 | Eu33200 | [X]Recurr depress disorder cur epi severe without psyc sympt | Feb-09 |
| Depression | 15099 | 21921 | 763 | 0 | 0 | E113.00 | Recurrent major depressive episode | Feb-09 |
| Depression | 7011 | 2881 | 288 | 0 | 0 | E112z00 | Single major depressive episode NOS | Feb-09 |
| Depression | 15220 | 8375 | 317 | 0 | 0 | Eu34114 | [X]Persistant anxiety depression | Feb-09 |
| Depression | 8584 | 1625 | 19 | 0 | 0 | Eu34111 | [X]Depressive neurosis | Feb-09 |
| Depression | 7737 | 6350 | 21 | 0 | 0 | Eu34113 | [X]Neurotic depression | Feb-09 |
| Depression | 42931 | 22725 | 3 | 0 | 0 | 9HA0.00 | On depression register | Feb-09 |
| Depression | 34390 | 1917 | 18 | 0 | 0 | E112000 | Single major depressive episode, unspecified | Feb-09 |
| Depression | 1131 | 311974 | 8026 | 2 | 0 | E204.00 | Neurotic depression reactive type | Feb-09 |
| Depression | 102632 | 1036 | 1 | 0 | 0 | 6659000 | Antidepressant drug treatment started | Jul-11 |
| Depression | 100977 | 7530 | 176 | 0 | 0 | 1JJ..00 | Suspected depression | Nov-10 |
| Depression | 98414 | 3024 | 17 | 0 | 0 | Eu32700 | [X]Major depression, severe without psychotic symptoms | Jan-10 |
| Depression | 2560 | 6909 | 411 | 0 | 0 | E11..12 | Depressive psychoses | Feb-09 |
| Depression | 11717 | 54544 | 872 | 0 | 0 | Eu32000 | [X]Mild depressive episode | Feb-09 |
| Depression | 32841 | 2926 | 1197 | 0 | 0 | 8HHq.00 | Referral for guided self-help for depression | Feb-09 |
| Depression | 96995 | 12 | 0 | 0 | 0 | 9kQ..00 | On full dose long term treatment depression - enh serv admin | Jun-09 |
| Depression | 72966 | 12782 | 12 | 0 | 0 | 9Ov1.00 | Depression monitoring second letter | Feb-09 |
| Depression | 24171 | 836 | 9 | 0 | 0 | E113400 | Recurrent major depressive episodes, severe, with psychosis | Feb-09 |
| Depression | 10015 | 642810 | 18493 | 0 | 0 | 1BT..00 | Depressed mood | Feb-09 |
| Depression | 22116 | 839 | 2 | 0 | 0 | Eu33400 | [X]Recurrent depressive disorder, currently in remission | Feb-09 |
| Depression | 7749 | 12327 | 73 | 0 | 0 | Eu41211 | [X]Mild anxiety depression | Feb-09 |
| Depression | 47009 | 1319 | 11 | 0 | 0 | Eu33300 | [X]Recurrent depress disorder cur epi severe with psyc symp | Feb-09 |
| Depression | 33426 | 83 | 1 | 0 | 0 | E11yz00 | Other and unspecified manic-depressive psychoses NOS | Feb-09 |
| Depression | 324 | 2136186 | 94133 | 94 | 0 | E2B..00 | Depressive disorder NEC | Feb-09 |
| Depression | 27491 | 253 | 14 | 0 | 0 | E11y200 | Atypical depressive disorder | Feb-09 |
| Depression | 1996 | 553328 | 18307 | 0 | 0 | 1B17.00 | Depressed | Feb-09 |
| Depression | 30483 | 57451 | 9 | 0 | 0 | 8CAa.00 | Patient given advice about management of depression | Feb-09 |
| Depression | 15219 | 3910 | 43 | 0 | 0 | E112300 | Single major depressive episode, severe, without psychosis | Feb-09 |
| Depression | 2970 | 156645 | 2360 | 0 | 0 | Eu32z00 | [X]Depressive episode, unspecified | Feb-09 |
| Depression | 98252 | 3030 | 16 | 0 | 0 | Eu32600 | [X]Major depression, moderately severe | Dec-09 |
| Depression | 56273 | 240 | 9 | 0 | 0 | E113500 | Recurrent major depressive episodes,partial/unspec remission | Feb-09 |
| Depression | 10610 | 58650 | 1093 | 0 | 0 | E112.00 | Single major depressive episode | Feb-09 |
| Depression | 9796 | 311684 | 13038 | 0 | 0 | 1B1U.00 | Symptoms of depression | Feb-09 |
| Depression | 15155 | 19317 | 173 | 0 | 0 | E112200 | Single major depressive episode, moderate | Feb-09 |
| Depression | 16632 | 2900 | 122 | 0 | 0 | E291.00 | Prolonged depressive reaction | Feb-09 |
| Depression | 6932 | 107071 | 5134 | 0 | 0 | E113.11 | Endogenous depression - recurrent | Feb-09 |
| Depression | 47731 | 343 | 3 | 0 | 0 | Eu33y00 | [X]Other recurrent depressive disorders | Feb-09 |
| Depression | 71009 | 41833 | 31 | 0 | 0 | 9Ov0.00 | Depression monitoring first letter | Feb-09 |
| Depression | 32941 | 423 | 27 | 0 | 0 | Eu33313 | [X]Recurr severe episodes/major depression+psychotic symptom | Feb-09 |
| Depression | 28677 | 159 | 1 | 0 | 0 | Eu33312 | [X]Manic-depress psychosis,depressed type+psychotic symptoms | Feb-09 |
| Depression | 23731 | 669 | 32 | 0 | 0 | Eu33311 | [X]Endogenous depression with psychotic symptoms | Feb-09 |
| Depression | 37764 | 107 | 7 | 0 | 0 | Eu33316 | [X]Recurrent severe episodes/reactive depressive psychosis | Feb-09 |
| Depression | 16861 | 609 | 20 | 0 | 0 | Eu33315 | [X]Recurrent severe episodes of psychotic depression | Feb-09 |
| Depression | 31757 | 63 | 3 | 0 | 0 | Eu33314 | [X]Recurr severe episodes/psychogenic depressive psychosis | Feb-09 |
| Depression | 4824 | 372645 | 15432 | 0 | 0 | 1B17.11 | C/O - feeling depressed | Feb-09 |
| Depression | 47701 | 11 | 1 | 0 | 0 | U606815 | [X] Adverse reaction to CNS muscle-tone depressants NOS | Feb-09 |
| Depression | 96038 | 1 | 0 | 0 | 0 | ZRLfI00 | Health of the Nation Outcome Scale item 7 - depressed mood | Feb-09 |
| Depression | 98346 | 972 | 1 | 0 | 0 | Eu32500 | [X]Major depression, mild | Jan-10 |
| Depression | 14709 | 19630 | 320 | 0 | 0 | E113200 | Recurrent major depressive episodes, moderate | Feb-09 |
| Depression | 43324 | 325 | 5 | 0 | 0 | E112500 | Single major depressive episode, partial or unspec remission | Feb-09 |
| Depression | 24117 | 346 | 20 | 0 | 0 | Eu32311 | [X]Single episode of major depression and psychotic symptoms | Feb-09 |
| Depression | 28863 | 242 | 4 | 0 | 0 | Eu32314 | [X]Single episode of reactive depressive psychosis | Feb-09 |
| Depression | 6950 | 84374 | 4736 | 0 | 0 | E112.13 | Endogenous depression first episode | Feb-09 |
| Depression | 6546 | 20549 | 386 | 0 | 0 | E112.12 | Endogenous depression first episode | Feb-09 |
| Depression | 5879 | 51877 | 3859 | 0 | 0 | E112.11 | Agitated depression | Feb-09 |
| Depression | 595 | 77747 | 2802 | 0 | 0 | E112.14 | Endogenous depression | Feb-09 |
| Depression | 10438 | 36804 | 1739 | 0 | 0 | 1B1U.11 | Depressive symptoms | Feb-09 |
| Depression | 10667 | 21623 | 201 | 0 | 0 | Eu32400 | [X]Mild depression | Feb-09 |
| Depression | 32845 | 126 | 5 | 0 | 0 | Eu92000 | [X]Depressive conduct disorder | Feb-09 |
| Depression | 12399 | 32023 | 93 | 0 | 0 | 9H90.00 | Depression annual review | Feb-09 |
| Depression | 1055 | 54058 | 3113 | 0 | 0 | E135.00 | Agitated depression | Feb-09 |
| Depression | 2716 | 419549 | 12998 | 13 | 0 | 1465 | H/O: depression | Feb-09 |
| Depression | 36616 | 48 | 2 | 0 | 0 | Eu33z11 | [X]Monopolar depression NOS | Feb-09 |
| Depression | 3291 | 21957 | 486 | 0 | 0 | Eu32z12 | [X]Depressive disorder NOS | Feb-09 |
| Depression | 9083 | 64503 | 1280 | 0 | 0 | 1285 | FH: Depression | Feb-09 |
| Depression | 19439 | 80062 | 101 | 0 | 0 | 212S.00 | Depression resolved | Feb-09 |
| Depression | 8902 | 4588 | 78 | 0 | 0 | Eu33.13 | [X]Recurrent episodes of reactive depression | Feb-09 |
| Depression | 4639 | 563425 | 6293 | 0 | 0 | Eu32.00 | [X]Depressive episode | Feb-09 |
| Depression | 8478 | 2317 | 54 | 0 | 0 | E130.00 | Reactive depressive psychosis | Feb-09 |
| Depression | 25697 | 3374 | 33 | 0 | 0 | E113300 | Recurrent major depressive episodes, severe, no psychosis | Feb-09 |
| Depression | 19696 | 915 | 29 | 0 | 0 | Eu33.12 | [X]Recurrent episodes of psychogenic depression | Feb-09 |
| Depression | 8851 | 3308 | 38 | 0 | 0 | Eu33.11 | [X]Recurrent episodes of depressive reaction | Feb-09 |
| Depression | 28756 | 154 | 2 | 0 | 0 | Eu33.14 | [X]Seasonal depressive disorder | Feb-09 |
| Depression | 32159 | 547 | 5 | 0 | 0 | E112400 | Single major depressive episode, severe, with psychosis | Feb-09 |
| Depression | 9183 | 3891 | 81 | 0 | 0 | E11z200 | Masked depression | Feb-09 |
| Depression | 30583 | 53882 | 83 | 0 | 0 | 9k4..00 | Depression - enhanced services administration | Feb-09 |
| Depression | 42104 | 11 | 6 | 351 | 0 | 32E4.00 | ECG: S-T depression | Feb-09 |
| Depression | 12099 | 6667 | 86 | 0 | 0 | Eu32300 | [X]Severe depressive episode with psychotic symptoms | Feb-09 |
| Depression | 4323 | 72098 | 2061 | 2 | 0 | E2B1.00 | Chronic depression | Feb-09 |
| Depression | 35671 | 767 | 12 | 0 | 0 | E113000 | Recurrent major depressive episodes, unspecified | Feb-09 |
| Depression | 25563 | 2174 | 11 | 0 | 0 | E113z00 | Recurrent major depressive episode NOS | Feb-09 |
| Depression | 52678 | 89 | 4 | 0 | 0 | Eu32312 | [X]Single episode of psychogenic depressive psychosis | Feb-09 |
| Depression | 44300 | 3419 | 23 | 0 | 0 | Eu33z00 | [X]Recurrent depressive disorder, unspecified | Feb-09 |
| Depression | 29784 | 3183 | 17 | 0 | 0 | Eu33000 | [X]Recurrent depressive disorder, current episode mild | Feb-09 |
| Depression | 29527 | 322 | 4 | 0 | 0 | R007z13 | [D]Postoperative depression | Feb-09 |
| Depression | 9055 | 10658 | 146 | 0 | 0 | Eu32.11 | [X]Single episode of depressive reaction | Feb-09 |
| Depression | 7604 | 8886 | 137 | 0 | 0 | Eu32.13 | [X]Single episode of reactive depression | Feb-09 |
| Depression | 18510 | 705 | 25 | 0 | 0 | Eu32.12 | [X]Single episode of psychogenic depression | Feb-09 |
| Depression | 17770 | 2862 | 88 | 0 | 0 | E130.11 | Psychotic reactive depression | Feb-09 |
| Depression | 56609 | 152 | 1 | 0 | 0 | Eu32y12 | [X]Single episode of masked depression NOS | Feb-09 |
| Depression | 10720 | 512 | 6 | 0 | 0 | Eu32y11 | [X]Atypical depression | Feb-09 |
| Depression | 5987 | 110440 | 2202 | 0 | 0 | Eu32z14 | [X] Reactive depression NOS | Feb-09 |
| Depression | 543 | 958613 | 24261 | 1 | 0 | Eu32z11 | [X]Depression NOS | Feb-09 |
| Depression | 28248 | 646 | 21 | 0 | 0 | Eu32z13 | [X]Prolonged single episode of reactive depression | Feb-09 |
| Depression | 3292 | 60397 | 637 | 0 | 0 | Eu33.00 | [X]Recurrent depressive disorder | Feb-09 |
| Depression | 9667 | 32593 | 615 | 0 | 0 | Eu32200 | [X]Severe depressive episode without psychotic symptoms | Feb-09 |
| Depression | 91105 | 6961 | 10 | 0 | 0 | 9Ov2.00 | Depression monitoring third letter | Feb-09 |
| Depression | 55384 | 525 | 1 | 0 | 0 | E113600 | Recurrent major depressive episodes, in full remission | Feb-09 |
| Depression | 73991 | 30 | 0 | 0 | 0 | Eu33214 | [X]Vital depression, recurrent without psychotic symptoms | Feb-09 |
| Depression | 11252 | 470 | 17 | 0 | 0 | Eu33212 | [X]Major depression, recurrent without psychotic symptoms | Feb-09 |
| Depression | 1908 | 115306 | 6024 | 0 | 0 | 2257 | O/E - depressed | Feb-09 |
| Depression | 65435 | 25629 | 1 | 0 | 0 | 9k40.00 | Depression - enhanced service completed | Feb-09 |
| Depression | 29520 | 8782 | 87 | 0 | 0 | Eu33100 | [X]Recurrent depressive disorder, current episode moderate | Feb-09 |
| Depression | 44848 | 180597 | 319 | 0 | 0 | 8BK0.00 | Depression management programme | Feb-09 |
| Depression | 51258 | 23010 | 108 | 0 | 0 | 9Ov..00 | Depression monitoring administration | Feb-09 |
| Depression | 16506 | 11388 | 191 | 0 | 0 | E112100 | Single major depressive episode, mild | Feb-09 |
| Depression | 57409 | 172 | 2 | 0 | 0 | E112600 | Single major depressive episode, in full remission | Feb-09 |
| Depression | 29342 | 4747 | 26 | 0 | 0 | E113100 | Recurrent major depressive episodes, mild | Feb-09 |
| Depression | 19054 | 346 | 1 | 0 | 0 | Eu3y111 | [X]Recurrent brief depressive episodes | Feb-09 |
| Depression | 98417 | 704 | 5 | 0 | 0 | Eu32800 | [X]Major depression, severe with psychotic symptoms | Jan-10 |
| Depression | 6854 | 3938 | 45 | 0 | 0 | Eu32y00 | [X]Other depressive episodes | Feb-09 |
| Depression | 24112 | 1031 | 12 | 0 | 0 | Eu32313 | [X]Single episode of psychotic depression | Feb-09 |
| Depression | 9211 | 120851 | 2356 | 0 | 0 | Eu32100 | [X]Moderate depressive episode | Feb-09 |
| Depression | 1533 | 4977 | 392 | 0 | 0 | E290.00 | Brief depressive reaction | Feb-09 |
| Depression | 11329 | 1295 | 20 | 0 | 0 | Eu33211 | [X]Endogenous depression without psychotic symptoms | Feb-09 |
| Depression | 6482 | 73561 | 2324 | 0 | 0 | E113700 | Recurrent depression | Feb-09 |
| Depression | 41989 | 194 | 2 | 0 | 0 | Eu32211 | [X]Single episode agitated depressn w'out psychotic symptoms | Feb-09 |
| Depression | 59386 | 80 | 4 | 0 | 0 | Eu32213 | [X]Single episode vital depression w'out psychotic symptoms | Feb-09 |
| Depression | 22806 | 3286 | 31 | 0 | 0 | Eu32212 | [X]Single episode major depression w'out psychotic symptoms | Feb-09 |
| Depression | 11913 | 57753 | 2068 | 1 | 0 | Eu41200 | [X]Mixed anxiety and depressive disorder | Feb-09 |
| Depression | 36246 | 199 | 1 | 0 | 0 | E290z00 | Brief depressive reaction NOS | Feb-09 |
| Depression | 53148 | 560 | 4 | 0 | 0 | 1BU..00 | Loss of hope for the future | Feb-09 |
| Depression | 59869 | 2553 | 20 | 0 | 0 | 1BP0.00 | Loss of interest in previously enjoyable activity | Feb-09 |
| Depression | 30740 | 27948 | 18 | 0 | 0 | 1BP..00 | Loss of interest | Feb-09 |
| Depression | 25435 | 6808 | 0 | 0 | 0 | 1BQ..00 | Loss of capacity for enjoyment | Feb-09 |
| Alcohol Misuse | 104611 | 773 | 1 | 0 | 0 | J670800 | Alcohol-induced acute pancreatitis | Jul-12 |
| Alcohol Misuse | 10691 | 9573 | 187 | 0 | 0 | J610.00 | Alcoholic fatty liver | Feb-09 |
| Alcohol Misuse | 11106 | 127 | 3 | 0 | 0 | E011100 | Korsakov's alcoholic psychosis with peripheral neuritis | Feb-09 |
| Alcohol Misuse | 11107 | 1275 | 19 | 0 | 0 | C253.00 | Wernicke's encephalopathy | Feb-09 |
| Alcohol Misuse | 11670 | 576 | 9 | 0 | 0 | Eu10611 | [X]Korsakov's psychosis, alcohol induced | Feb-09 |
| Alcohol Misuse | 11740 | 69219 | 475 | 0 | 0 | 9k1..00 | Alcohol misuse - enhanced services administration | Feb-09 |
| Alcohol Misuse | 12353 | 311 | 4 | 0 | 0 | Eu10500 | [X]Mental & behav dis due to use alcohol: psychotic disorder | Feb-09 |
| Alcohol Misuse | 12442 | 22725 | 306 | 0 | 0 | 66e..00 | Alcohol disorder monitoring | Feb-09 |
| Alcohol Misuse | 1399 | 172056 | 10068 | 25 | 0 | E23..12 | Alcohol problem drinking | Feb-09 |
| Alcohol Misuse | 1476 | 2519 | 246 | 0 | 0 | E010.12 | Delirium tremens | Feb-09 |
| Alcohol Misuse | 16225 | 2278 | 90 | 0 | 0 | E010.00 | Alcohol withdrawal delirium | Feb-09 |
| Alcohol Misuse | 16237 | 851 | 41 | 0 | 0 | E01..00 | Alcoholic psychoses | Feb-09 |
| Alcohol Misuse | 16587 | 182 | 20 | 0 | 0 | ZV11311 | [V]Problems related to lifestyle alcohol use | Feb-09 |
| Alcohol Misuse | 17259 | 2255 | 55 | 1 | 0 | Eu10411 | [X]Delirium tremens, alcohol induced | Feb-09 |
| Alcohol Misuse | 17330 | 595 | 32 | 0 | 0 | J613000 | Alcoholic hepatic failure | Feb-09 |
| Alcohol Misuse | 17607 | 876 | 30 | 0 | 0 | Eu10514 | [X]Alcoholic psychosis NOS | Feb-09 |
| Alcohol Misuse | 18156 | 2570 | 23 | 2 | 0 | 13Y8.00 | Alcoholics anonymous | Feb-09 |
| Alcohol Misuse | 18636 | 733 | 22 | 0 | 0 | E011200 | Wernicke-Korsakov syndrome | Feb-09 |
| Alcohol Misuse | 19494 | 24376 | 564 | 0 | 0 | 136S.00 | Hazardous alcohol use | Feb-09 |
| Alcohol Misuse | 20514 | 384 | 2 | 0 | 0 | Eu10300 | [X]Mental and behav dis due to use alcohol: withdrawal state | Feb-09 |
| Alcohol Misuse | 20762 | 120 | 1 | 0 | 0 | E011.00 | Alcohol amnestic syndrome | Feb-09 |
| Alcohol Misuse | 2081 | 67031 | 4459 | 21 | 0 | E23..11 | Alcoholism | Feb-09 |
| Alcohol Misuse | 2082 | 53525 | 1471 | 2 | 0 | E01y000 | Alcohol withdrawal syndrome | Feb-09 |
| Alcohol Misuse | 2083 | 89342 | 2822 | 4 | 0 | 8BA8.00 | Alcohol detoxification | Feb-09 |
| Alcohol Misuse | 2084 | 291183 | 12383 | 16 | 0 | E23..00 | Alcohol dependence syndrome | Feb-09 |
| Alcohol Misuse | 21624 | 163 | 2 | 0 | 0 | E230200 | Episodic acute alcoholic intoxication in alcoholism | Feb-09 |
| Alcohol Misuse | 21650 | 3314 | 199 | 0 | 0 | 8H35.00 | Admitted to alcohol detoxification centre | Feb-09 |
| Alcohol Misuse | 21713 | 405 | 10 | 0 | 0 | J612000 | Alcoholic fibrosis and sclerosis of liver | Feb-09 |
| Alcohol Misuse | 21879 | 622 | 5 | 0 | 0 | Eu10100 | [X]Mental and behav dis due to use of alcohol: harmful use | Feb-09 |
| Alcohol Misuse | 22277 | 1302 | 34 | 0 | 0 | E010.11 | DTs - delirium tremens | Feb-09 |
| Alcohol Misuse | 24064 | 641 | 5 | 0 | 0 | E231100 | Continuous chronic alcoholism | Feb-09 |
| Alcohol Misuse | 24485 | 554 | 1 | 0 | 0 | E231300 | Chronic alcoholism in remission | Feb-09 |
| Alcohol Misuse | 24984 | 1179 | 7 | 0 | 0 | J671000 | Alcohol-induced chronic pancreatitis | Feb-09 |
| Alcohol Misuse | 25110 | 418 | 31 | 0 | 0 | E013.00 | Alcohol withdrawal hallucinosis | Feb-09 |
| Alcohol Misuse | 26106 | 710 | 13 | 0 | 0 | E231200 | Episodic chronic alcoholism | Feb-09 |
| Alcohol Misuse | 26323 | 1001 | 23 | 0 | 0 | Eu10711 | [X]Alcoholic dementia NOS | Feb-09 |
| Alcohol Misuse | 27342 | 1175 | 34 | 0 | 0 | E012.11 | Alcoholic dementia NOS | Feb-09 |
| Alcohol Misuse | 28780 | 4625 | 386 | 0 | 0 | Eu10211 | [X]Alcohol addiction | Feb-09 |
| Alcohol Misuse | 2925 | 1620 | 65 | 0 | 0 | F375.00 | Alcoholic polyneuropathy | Feb-09 |
| Alcohol Misuse | 29691 | 507 | 12 | 0 | 0 | 8G32.00 | Aversion therapy - alcoholism | Feb-09 |
| Alcohol Misuse | 30162 | 161 | 11 | 0 | 0 | Eu10513 | [X]Alcoholic paranoia | Feb-09 |
| Alcohol Misuse | 30404 | 226 | 16 | 0 | 0 | E015.00 | Alcoholic paranoia | Feb-09 |
| Alcohol Misuse | 30460 | 1430 | 130 | 0 | 0 | Z4B1.00 | Alcoholism counselling | Feb-09 |
| Alcohol Misuse | 30604 | 826 | 2 | 0 | 0 | F25B.00 | Alcohol-induced epilepsy | Feb-09 |
| Alcohol Misuse | 30695 | 27827 | 822 | 0 | 0 | 136T.00 | Harmful alcohol use | Feb-09 |
| Alcohol Misuse | 31443 | 5082 | 104 | 0 | 0 | E231.00 | Chronic alcoholism | Feb-09 |
| Alcohol Misuse | 31742 | 206 | 2 | 0 | 0 | F394100 | Alcoholic myopathy | Feb-09 |
| Alcohol Misuse | 3216 | 5423 | 250 | 1 | 0 | J611.00 | Acute alcoholic hepatitis | Feb-09 |
| Alcohol Misuse | 32927 | 11998 | 56 | 0 | 0 | Eu10800 | [X]Alcohol withdrawal-induced seizure | Feb-09 |
| Alcohol Misuse | 32964 | 11094 | 310 | 0 | 0 | 66e0.00 | Alcohol abuse monitoring | Feb-09 |
| Alcohol Misuse | 33635 | 3459 | 129 | 0 | 0 | E231z00 | Chronic alcoholism NOS | Feb-09 |
| Alcohol Misuse | 33670 | 340 | 0 | 0 | 0 | E01y.00 | Other alcoholic psychosis | Feb-09 |
| Alcohol Misuse | 33839 | 126 | 0 | 0 | 0 | F144000 | Cerebellar ataxia due to alcoholism | Feb-09 |
| Alcohol Misuse | 36296 | 474 | 3 | 0 | 0 | E230z00 | Acute alcoholic intoxication in alcoholism NOS | Feb-09 |
| Alcohol Misuse | 36748 | 809 | 12 | 0 | 0 | F11x011 | Alcoholic encephalopathy | Feb-09 |
| Alcohol Misuse | 37605 | 24 | 3 | 0 | 0 | E231.11 | Dipsomania | Feb-09 |
| Alcohol Misuse | 37691 | 422 | 4 | 0 | 0 | Eu10712 | [X]Chronic alcoholic brain syndrome | Feb-09 |
| Alcohol Misuse | 37946 | 102 | 0 | 0 | 0 | E012000 | Chronic alcoholic brain syndrome | Feb-09 |
| Alcohol Misuse | 38061 | 955 | 9 | 0 | 0 | 1B1c.00 | Alcohol induced hallucinations | Feb-09 |
| Alcohol Misuse | 39327 | 2125 | 21 | 0 | 0 | Eu10200 | [X]Mental and behav dis due to use alcohol: dependence syndr | Feb-09 |
| Alcohol Misuse | 39799 | 179 | 1 | 0 | 0 | Eu10600 | [X]Mental and behav dis due to use alcohol: amnesic syndrome | Feb-09 |
| Alcohol Misuse | 40530 | 278 | 3 | 0 | 0 | E230000 | Acute alcoholic intoxication, unspecified, in alcoholism | Feb-09 |
| Alcohol Misuse | 41920 | 9 | 1 | 0 | 0 | E011z00 | Alcohol amnestic syndrome NOS | Feb-09 |
| Alcohol Misuse | 41983 | 348 | 1 | 0 | 0 | Z191.00 | Alcohol detoxification | Feb-09 |
| Alcohol Misuse | 43193 | 330 | 0 | 0 | 0 | E231000 | Unspecified chronic alcoholism | Feb-09 |
| Alcohol Misuse | 4500 | 2495 | 67 | 0 | 0 | E011000 | Korsakov's alcoholic psychosis | Feb-09 |
| Alcohol Misuse | 4501 | 263 | 18 | 0 | 0 | C251.11 | Wernicke's encephalopathy | Feb-09 |
| Alcohol Misuse | 4506 | 12919 | 195 | 10 | 0 | J153.00 | Alcoholic gastritis | Feb-09 |
| Alcohol Misuse | 45169 | 30 | 1 | 0 | 0 | Eu10y00 | [X]Men & behav dis due to use alcohol: oth men & behav dis | Feb-09 |
| Alcohol Misuse | 46677 | 205 | 1 | 0 | 0 | Z191100 | Alcohol withdrawal regime | Feb-09 |
| Alcohol Misuse | 4743 | 18242 | 525 | 3 | 0 | J612.00 | Alcoholic cirrhosis of liver | Feb-09 |
| Alcohol Misuse | 47555 | 151 | 1 | 0 | 0 | F11x000 | Cerebral degeneration due to alcoholism | Feb-09 |
| Alcohol Misuse | 4915 | 1590 | 25 | 0 | 0 | G555.00 | Alcoholic cardiomyopathy | Feb-09 |
| Alcohol Misuse | 54505 | 266 | 3 | 0 | 0 | E012.00 | Other alcoholic dementia | Feb-09 |
| Alcohol Misuse | 5611 | 2502 | 59 | 0 | 0 | Eu10.00 | [X]Mental and behavioural disorders due to use of alcohol | Feb-09 |
| Alcohol Misuse | 56410 | 673 | 192 | 0 | 0 | 7P22100 | Delivery of rehabilitation for alcohol addiction | Feb-09 |
| Alcohol Misuse | 56947 | 71 | 2 | 0 | 0 | E230100 | Continuous acute alcoholic intoxication in alcoholism | Feb-09 |
| Alcohol Misuse | 5740 | 8424 | 460 | 3 | 0 | E230.00 | Acute alcoholic intoxication in alcoholism | Feb-09 |
| Alcohol Misuse | 5758 | 2843 | 68 | 0 | 0 | Eu10212 | [X]Chronic alcoholism | Feb-09 |
| Alcohol Misuse | 57714 | 132 | 0 | 0 | 0 | E230.11 | Alcohol dependence with acute alcoholic intoxication | Feb-09 |
| Alcohol Misuse | 59574 | 180 | 1 | 0 | 0 | E230300 | Acute alcoholic intoxication in remission, in alcoholism | Feb-09 |
| Alcohol Misuse | 61383 | 13 | 0 | 0 | 0 | Z191200 | Planned reduction of alcohol consumption | Feb-09 |
| Alcohol Misuse | 6169 | 12133 | 294 | 0 | 0 | E23z.00 | Alcohol dependence syndrome NOS | Feb-09 |
| Alcohol Misuse | 62000 | 39 | 0 | 0 | 0 | Eu10700 | [X]Men & behav dis due alcoh: resid & late-onset psychot dis | Feb-09 |
| Alcohol Misuse | 63529 | 8949 | 0 | 0 | 0 | 9k12.00 | Alcohol misuse - enhanced service completed | Feb-09 |
| Alcohol Misuse | 64101 | 150 | 3 | 0 | 0 | Eu10400 | [X]Men & behav dis due alcohl: withdrawl state with delirium | Feb-09 |
| Alcohol Misuse | 64389 | 34 | 0 | 0 | 0 | Eu10z00 | [X]Ment & behav dis due use alcohol: unsp ment & behav dis | Feb-09 |
| Alcohol Misuse | 6467 | 671 | 10 | 0 | 0 | Eu10511 | [X]Alcoholic hallucinosis | Feb-09 |
| Alcohol Misuse | 65754 | 24 | 0 | 0 | 0 | C150500 | Alcohol-induced pseudo-Cushing's syndrome | Feb-09 |
| Alcohol Misuse | 65932 | 22 | 0 | 0 | 0 | Eu10512 | [X]Alcoholic jealousy | Feb-09 |
| Alcohol Misuse | 67651 | 170 | 0 | 0 | 0 | E01z.00 | Alcoholic psychosis NOS | Feb-09 |
| Alcohol Misuse | 68111 | 19 | 0 | 0 | 0 | E01yz00 | Other alcoholic psychosis NOS | Feb-09 |
| Alcohol Misuse | 69691 | 2 | 0 | 0 | 0 | Eu10213 | [X]Dipsomania | Feb-09 |
| Alcohol Misuse | 7123 | 2110 | 49 | 0 | 0 | ZV11300 | [V]Personal history of alcoholism | Feb-09 |
| Alcohol Misuse | 7602 | 196 | 3 | 0 | 0 | J617000 | Chronic alcoholic hepatitis | Feb-09 |
| Alcohol Misuse | 7885 | 16601 | 477 | 3 | 0 | J613.00 | Alcoholic liver damage unspecified | Feb-09 |
| Alcohol Misuse | 7943 | 5217 | 89 | 0 | 0 | J617.00 | Alcoholic hepatitis | Feb-09 |
| Alcohol Misuse | 8030 | 27527 | 1072 | 0 | 0 | ZV6D600 | [V]Alcohol abuse counselling and surveillance | Feb-09 |
| Alcohol Misuse | 8363 | 613 | 13 | 0 | 0 | G852300 | Oesophageal varices in alcoholic cirrhosis of the liver | Feb-09 |
| Alcohol Misuse | 8388 | 1851 | 68 | 0 | 0 | ZV57A00 | [V]Alcohol rehabilitation | Feb-09 |
| Alcohol Misuse | 8430 | 10770 | 164 | 1 | 0 | 1462 | H/O: alcoholism | Feb-09 |
| Alcohol Misuse | 94553 | 2373 | 2169 | 0 | 0 | 8HkG.00 | Referral to specialist alcohol treatment service | Feb-09 |
| Alcohol Misuse | 94670 | 9384 | 90 | 0 | 0 | 136W.00 | Alcohol misuse | Feb-09 |
| Alcohol Misuse | 9489 | 9929 | 153 | 0 | 0 | 9NN2.00 | Under care of community alcohol team | Feb-09 |
| Alcohol Misuse | 95181 | 24 | 1 | 0 | 0 | Z191211 | Alcohol reduction programme | Feb-09 |
| Alcohol Misuse | 96053 | 181370 | 37 | 2 | 0 | 9k1A.00 | Brief intervention for excessive alcohol consumptn completed | Feb-09 |
| Alcohol Misuse | 96054 | 7009 | 2 | 0 | 0 | 9k1B.00 | Extended intervention for excessive alcohol consumptn complt | Feb-09 |
| Alcohol Misuse | 96993 | 487 | 67 | 0 | 0 | 8HkJ.00 | Referral to alcohol brief intervention service | Jun-09 |
| Alcohol Misuse | 97309 | 1536 | 1 | 0 | 0 | 8CAv.00 | Advised to contact primary care alcohol worker | Aug-09 |
| Alcohol Misuse | 9849 | 15888 | 19639 | 0 | 0 | 8H7p.00 | Referral to community alcohol team | Feb-09 |
| Dyslipidaemia | 339 | 540024 | 13500 | 694 | 0 | C320.00 | Pure hypercholesterolaemia | Feb-09 |
| Dyslipidaemia | 637 | 448767 | 8244 | 472 | 0 | C324.00 | Hyperlipidaemia NOS | Feb-09 |
| Dyslipidaemia | 2493 | 59226 | 41476 | 263490 | 0 | 44P3.00 | Serum cholesterol raised | Feb-09 |
| Dyslipidaemia | 3386 | 16590 | 409 | 2 | 0 | C320000 | Familial hypercholesterolaemia | Feb-09 |
| Dyslipidaemia | 6243 | 435959 | 276 | 3 | 0 | 8CA4700 | Patient advised re low cholesterol diet | Feb-09 |
| Dyslipidaemia | 7447 | 26294 | 110 | 0 | 0 | C320z00 | Pure hypercholesterolaemia NOS | Feb-09 |
| Dyslipidaemia | 10783 | 28472 | 130 | 4 | 0 | 8BAG.00 | Cholesterol reduction programme | Feb-09 |
| Dyslipidaemia | 10899 | 371 | 1 | 0 | 0 | 8BAG200 | Cholesterol reduction program - declined | Feb-09 |
| Dyslipidaemia | 12569 | 3405 | 97 | 0 | 0 | ZV65317 | [V]Dietary surveillance in hypercholesterolaemia | Feb-09 |
| Dyslipidaemia | 26019 | 853 | 10 | 0 | 0 | C320200 | Hyperlipidaemia, group A | Feb-09 |
| Dyslipidaemia | 33694 | 269 | 1 | 0 | 0 | ZC2CJ00 | Dietary advice for hyperlipidaemia | Feb-09 |
| Dyslipidaemia | 35720 | 316 | 112 | 6609 | 0 | 44P4.00 | Serum cholesterol very high | Feb-09 |
| Dyslipidaemia | 39147 | 2563 | 16 | 0 | 0 | 8BAG000 | Cholesterol reduction programme - invited | Feb-09 |
| Dyslipidaemia | 50923 | 117 | 0 | 0 | 0 | U60C600 | [X]Antihyperlipidaem/antiarterioscl drg caus adv ef ther use | Feb-09 |
| Dyslipidaemia | 51023 | 1771 | 10 | 0 | 0 | 8BAG100 | Cholesterol reduction program - attended | Feb-09 |
| Dyslipidaemia | 53091 | 463 | 1 | 0 | 0 | C320y00 | Other specified pure hypercholesterolaemia | Feb-09 |
| Dyslipidaemia | 66240 | 107 | 0 | 0 | 0 | Cyu8D00 | [X]Other hyperlipidaemia | Feb-09 |
| Dyslipidaemia | 71747 | 40 | 2 | 0 | 0 | 8CR3.00 | Hyperlipidaemia clinical management plan | Feb-09 |
| Dyslipidaemia | 102390 | 552 | 19 | 0 | 0 | C322000 | Familial combined hyperlipidaemia | Jun-11 |
| Dyslipidaemia | 102958 | 333 | 0 | 0 | 0 | C320600 | Polygenic hypercholesterolaemia | Sep-11 |
| Dyslipidaemia | 107252 | 30215 | 322 | 128 | 0 | C329.00 | Hypercholesterolaemia | Nov-13 |
| Dyslipidaemia | 109744 | 68 | 8 | 0 | 0 | 8OAK.00 | Provsn written information about diabetes & high cholesterol | Jun-15 |
| Dyslipidaemia | 1173 | 7756 | 952 | 411 | 0 | C321.00 | Pure hyperglyceridaemia | Feb-09 |
| Dyslipidaemia | 37273 | 453 | 4 | 1 | 0 | C320400 | Fredrickson's hyperlipoproteinaemia, type IIa | Feb-09 |
| Dyslipidaemia | 52992 | 99 | 0 | 0 | 0 | C322.11 | Fredrickson type IIb lipidaemia | Feb-09 |
| Dyslipidaemia | 59564 | 6 | 0 | 0 | 0 | C322.12 | Fredrickson type III lipidaemia | Feb-09 |
| Dyslipidaemia | 104941 | 1 | 0 | 0 | 0 | C323.13 | Fredrickson type V lipaemia | Sep-12 |
| Dyslipidaemia | 69881 | 5 | 0 | 0 | 0 | C323.12 | Fredrickson type I lipaemia | Feb-09 |
| Dyslipidaemia | 55855 | 16 | 1 | 7 | 0 | C320.12 | Fredrickson type IIa lipidaemia | Feb-09 |
| Dyslipidaemia | 54499 | 47 | 0 | 0 | 0 | C321.11 | Fredrickson type IV lipidaemia | Feb-09 |
| Dyslipidaemia | 34825 | 70 | 0 | 0 | 0 | C320100 | Hyperbetalipoproteinaemia | Feb-09 |
| Dyslipidaemia | 34224 | 3560 | 4 | 1 | 0 | C320300 | Low-density-lipoprotein-type (LDL) hyperlipoproteinaemia | Feb-09 |
| Dyslipidaemia | 91603 | 1 | 0 | 0 | 0 | C325.11 | Tangier disease | Feb-09 |

| outcome | ICD-10 code |
| --- | --- |
| Atrial Fibrillation and Flutter | I48 |
| Chronic Kidney Disease | N18.3 |
| Chronic Kidney Disease | N18.4 |
| Chronic Kidney Disease | N18.5 |
| Chronic Kidney Disease | N19 |
| Chronic Kidney Disease | N03 |
| Chronic Kidney Disease | N04 |
| Chronic Kidney Disease | N05 |
| Chronic Kidney Disease | N11 |
| Chronic Kidney Disease | T86.1 |
| Chronic Kidney Disease | Z94.0 |
| Chronic Kidney Disease | Z49 |
| Chronic Kidney Disease | Y84.1 |
| Chronic Kidney Disease | Z99. 2 |
| Erectile Dysfunction: | N48.4 |
| Erectile Dysfunction: | F52.2 |
| Erectile Dysfunction: | N52.9 |
| Family History of Young Myocardial Infarction or Angina | Z82.4 |
| Hypertension | O10 |
| Hypertension | O11 |
| Hypertension | I10 |
| Hypertension | I11 |
| Hypertension | I12 |
| Hypertension | I13 |
| Hypertension | I15 |
| Migraine | G43 |
| Obesity | E65 |
| Obesity | E66 |
| Severe Mental Illness | F20 |
| Severe Mental Illness | F21 |
| Severe Mental Illness | F22 |
| Severe Mental Illness | F23 |
| Severe Mental Illness | F24 |
| Severe Mental Illness | F25 |
| Severe Mental Illness | F28 |
| Severe Mental Illness | F29 |
| Severe Mental Illness | F44 |
| Severe Mental Illness | F52. 1 |
| Severe Mental Illness | F09 |
| Severe Mental Illness | F39 |
| Severe Mental Illness | F06.8 |
| Severe Mental Illness | F31 |
| Severe Mental Illness | F06.3 |
| Severe Mental Illness | F30 |
| Smoking Status | Z72.0 |
| Smoking Status | F17 |
| Smoking Status | Z71.6 |
| Smoking Status | T65.2 |
| Diabetes Mellitus | E10 |
| Diabetes Mellitus | E11 |
| Diabetes Mellitus | E12 |
| Diabetes Mellitus | E13 |
| Diabetes Mellitus | E14 |
| Diabetes Mellitus | H36.0 |
| Diabetes Mellitus | M14.2 |
| Diabetes Mellitus | G59.0 |
| Diabetes Mellitus | H28.0 |
| Diabetes Mellitus | G63.2 |
| Diabetes Mellitus | O24.0 |
| Diabetes Mellitus | O24.1 |
| Diabetes Mellitus | O24. 2 |
| Diabetes Mellitus | O24. 3 |
| Diabetes Mellitus | N08. 3 |
| Depression | F32 |
| Depression | F33 |
| Depression | F34.1 |
| Depression | F38.1 |
| Depression | F53.0 |
| Depression | F92.0 |
| Depression | F41.2 |
| Depression | F38.1 |
| Alcohol Misuse | E24.4 |
| Alcohol Misuse | F10 |
| Alcohol Misuse | G31.2 |
| Alcohol Misuse | G62.1 |
| Alcohol Misuse | G72.1 |
| Alcohol Misuse | I42.6 |
| Alcohol Misuse | K29.2 |
| Alcohol Misuse | K70 |
| Alcohol Misuse | K85.2 |
| Alcohol Misuse | K86.0 |
| Alcohol Misuse | Z50.2 |
| Alcohol Misuse | Z71.4 |
| Dyslipidaemia | E78 |
