## Supplement 2 for "Increased risk of major ischaemic events among autistic people"

**Supplement 2 – Autism Medcode List:**

| exposure | medcode | clinicalevents | referralevents | testevents | immunisationevents | readcode | readterm | databasebuild |
| --- | --- | --- | --- | --- | --- | --- | --- | --- |
| Autism | 1276 | 51334 | 2028 | 1 | 0 | E140.12 | Autism | Feb-09 |
| Autism | 42941 | 7764 | 14 | 0 | 0 | Eu84z11 | [X]Autistic spectrum disorder | Feb-09 |
| Autism | 7302 | 831 | 8 | 0 | 0 | E140.13 | Childhood autism | Feb-09 |
| Autism | 50337 | 43 | 2 | 0 | 0 | Eu84012 | [X]Infantile autism | Feb-09 |
| Autism | 9982 | 27638 | 560 | 0 | 0 | Eu84011 | [X]Autistic disorder | Feb-09 |
| Autism | 22098 | 2773 | 65 | 0 | 0 | E140.00 | Infantile autism | Feb-09 |
| Autism | 3637 | 2051 | 29 | 0 | 0 | Eu84000 | [X]Childhood autism | Feb-09 |
| Autism | 36662 | 158 | 1 | 0 | 0 | E140z00 | Infantile autism NOS | Feb-09 |
| Autism | 34174 | 162 | 3 | 0 | 0 | Eu84112 | [X]Mental retardation with autistic features | Feb-09 |
| Autism | 63251 | 50 | 1 | 0 | 0 | E140000 | Active infantile autism | Feb-09 |
| Autism | 110478 | 3 | 0 | 0 | 0 | Eu84014 | [X]Kanner's syndrome | Jan-16 |
| Autism | 43444 | 19 | 0 | 0 | 0 | E140.11 | Kanner's syndrome | Feb-09 |
| Autism | 46429 | 19 | 0 | 0 | 0 | Eu84313 | [X]Heller's syndrome | Feb-09 |
| Autism | 31599 | 15 | 0 | 0 | 0 | E141.11 | Heller's syndrome | Feb-09 |
| Autism | 56143 | 14 | 0 | 0 | 0 | E141.00 | Disintegrative psychosis | Feb-09 |
| Autism | 68299 | 20 | 0 | 0 | 0 | Eu84300 | [X]Other childhood disintegrative disorder | Feb-09 |
| Autism | 41207 | 1 | 0 | 0 | 0 | E141100 | Residual disintegrative psychoses | Feb-09 |
| Autism | 62222 | 2 | 0 | 0 | 0 | Eu84312 | [X]Disintegrative psychosis | Feb-09 |
| Autism | 61304 | 8 | 0 | 0 | 0 | Eu84013 | [X]Infantile psychosis | Feb-09 |
| Autism | 101999 | 2 | 0 | 0 | 0 | Eu84311 | [X]Dementia infantalis | Apr-11 |
| Autism | 2950 | 29313 | 1476 | 0 | 0 | Eu84500 | [X]Asperger's syndrome | Feb-09 |
| Autism | 69016 | 11 | 0 | 0 | 0 | E140100 | Residual infantile autism | Feb-09 |
| Autism | 44327 | 191 | 0 | 0 | 0 | Eu84z00 | [X]Pervasive developmental disorder, unspecified | Feb-09 |
| Autism | 7226 | 942 | 11 | 0 | 0 | Eu84.00 | [X]Pervasive developmental disorders | Feb-09 |
| Autism | 47948 | 14 | 0 | 0 | 0 | Eu84y00 | [X]Other pervasive developmental disorders | Feb-09 |
| Autism | 24044 | 725 | 8 | 0 | 0 | Eu84100 | [X]Atypical autism | Feb-09 |
